# Genetic influences on the shape of brain ventricular and subcortical structures

**DOI:** 10.1101/2022.09.26.22279691

**Authors:** Bingxin Zhao, Tengfei Li, Xiaochen Yang, Juan Shu, Xifeng Wang, Tianyou Luo, Yue Yang, Zhenyi Wu, Zirui Fan, Zhiwen Jiang, Jie Chen, Yue Shan, Jiarui Tang, Di Xiong, Ziliang Zhu, Mufeng Gao, Wyliena Guan, Chalmer E. Tomlinson, Qunxi Dong, Yun Li, Jason L. Stein, Yalin Wang, Hongtu Zhu

## Abstract

Brain ventricular and subcortical structures are heritable both in size and shape. Genetic influences on brain region size have been studied using conventional volumetric measures, but little is known about the genetic basis of ventricular and subcortical shapes. Here we developed pipelines to extract seven complementary shape measures for lateral ventricles, subcortical structures, and hippocampal subfields. Based on over 45,000 subjects in the UK Biobank and ABCD studies, 60 genetic loci were identified to be associated with brain shape features (*P* < 1.09 × 10^-10^), 19 of which were not detectable by volumetric measures of these brain structures. Ventricular and subcortical shape features were genetically related to cognitive functions, mental health traits, and multiple brain disorders, such as the attention-deficit/hyperactivity disorder. Vertex-based shape analysis was performed to precisely localize the brain regions with these shared genetic influences. Mendelian randomization suggests brain shape causally contributes to neurological and neuropsychiatric disorders, including Alzheimer’s disease and schizophrenia. Our results uncover the genetic architecture of brain shape for ventricular and subcortical structures and prioritize the genetic factors underlying disease-related shape variations.

---

Human brain ventricular and subcortical structures are involved in complex brain activities and have important roles in the regulation of cognitive, emotional, and motor functions^1–7^. Morphometric variations of these brain structures can be quantified in-vivo by structural magnetic resonance imaging (MRI). MRI-based volumetric measures (such as regional brain volumes) can estimate a region’s overall size, providing a conventional measure of the gross variation of the structure. However, such aggregate measures may not be sensitive to within-region local changes and may not fully capture the complexity of structural deformations. Shape analysis has gained increasing attention to overcome these limitations and characterize brain morphometry beyond simple volumetric traits^8–11^. Recent studies have found that shape features can precisely localize shape deformations in brain structures, providing finer-grained information of the location and pattern of morphological variations, which may not be detectable in traditional volume analysis^12, 13^. For example, shape analysis of the ventricular and subcortical structures has provided sensitive biomarkers for healthy aging^14^ and the onset and progression of a wide range of brain diseases, including Alzheimer’s disease (AD)^15, 16^, schizophrenia^17, 18^, epilepsy^19^, major depressive disorder (MDD)^20, 21^, 22q11.2 deletion syndrome^22^, and bipolar disorder^23^.

Both size (volume) and shape of brain ventricular and subcortical structures have been found to be heritable in family studies^24, 25^ and general populations^13, 26–28^. For example, the narrow sense single-nucleotide polymorphism (SNP) heritability estimates for the volume of ventricular and subcortical structures were all higher or close to 40%^26, 27^ in the UK Biobank^29^ (UKB) studies, and the highest SNP heritability estimates for shape features ranged from 32.7% to 53.3% across structures in the Rotterdam Study^28^. Genome-wide association studies (GWAS) have been conducted to uncover the genetic basis of ventricular and subcortical volumes^30–40^, yielding hundreds of associated genetic variants and shared genetic influences with brain disorders and complex traits. However, there is no large-scale GWAS on ventricular and subcortical shape features and their genetic architecture has yet to be determined.

Using raw MRI from over 45,000 subjects in the UKB and Adolescent Brain Cognitive Development^41^ (ABCD) studies, we developed pipelines to extract ventricular and subcortical shape features and characterized their genetic architectures. We identified 60 novel genetic loci that contributing to the shape variations, 19 of which cannot be identified in previous GWAS of volumetric measures of these brain structures using the same datasets. We found ventricular and subcortical shape features had shared genetic influences with many cognitive traits and major brain disorders. We further revealed the localized pattern of genetic effects in vertex-wise analysis and identified causal genetic links between brain shape and disorders using Mendelian randomization analysis. The results of this shape study demonstrated genetic effects on ventricular and subcortical structures at a finer spatial resolution than that of traditional volumetric analysis. Our GWAS results will be available through the Brain Imaging Genetics Knowledge Portal (BIG-KP) https://bigkp.org/.

## RESULTS

### Generating reproducible ventricular and subcortical shape features

We developed pipelines to extract shape features from raw structural MRI images for 8 ventricular and subcortical structures, including the left/right lateral ventricles, nucleus accumbens, amygdala, caudate, hippocampus, pallidum, putamen, and thalamus. Furthermore, we studied 7 subfields of the hippocampus, namely the left/right cornu ammonis 1 (CA1), CA3, fimbria, hippocampus-amygdala-transition-area (HATA), hippocampal tail, presubiculum, and subiculum (Fig.1A). An overview of our workflow can be found in **Fig. S1**. Shape deformations are usually decomposed into two components: one within the surfaces, and the other along the normal axis of the surfaces. For each vertex in the shape image, we calculated 7 complementary shape statistics, including the radial distance from the medial model^14^ (referred to as the radial distance); the (log) determinant and two eigenvalues of the Jacobian matrix from the surface tensor-based morphometry (TBM) model^42^ (referred to as the determinant, eigenvalue1, and eigenvalue2); and three features from the multivariate surface TBM (mTBM) model^10, 11^ (referred to as the mTBM1, mTBM2, and mTBM3) (Figs.1B and **S2**). Briefly, the radial distance describes morphometric changes along the surface normal direction^11^ and is also called the radial thickness^10^. On the other hand, the 6 surface TBM and mTBM model features capture surface deformations perpendicular to the surface normal axis (such as rotation, dilation, and shear within the surfaces). For example, the determinant of Jacobian matrix is analogous to a surface area^43^, measuring local area dilation or contraction. It quantifies the surface dilation ratio between the given template and the study subject by matching a small surface patch around a particular point of the subject surface to the corresponding point on the template. The three advanced surface mTBM features analyze the full surface tensor using log-Euclidean metrics and can capture more complicated surface deformations^10, 11^.

**Fig. 1.**
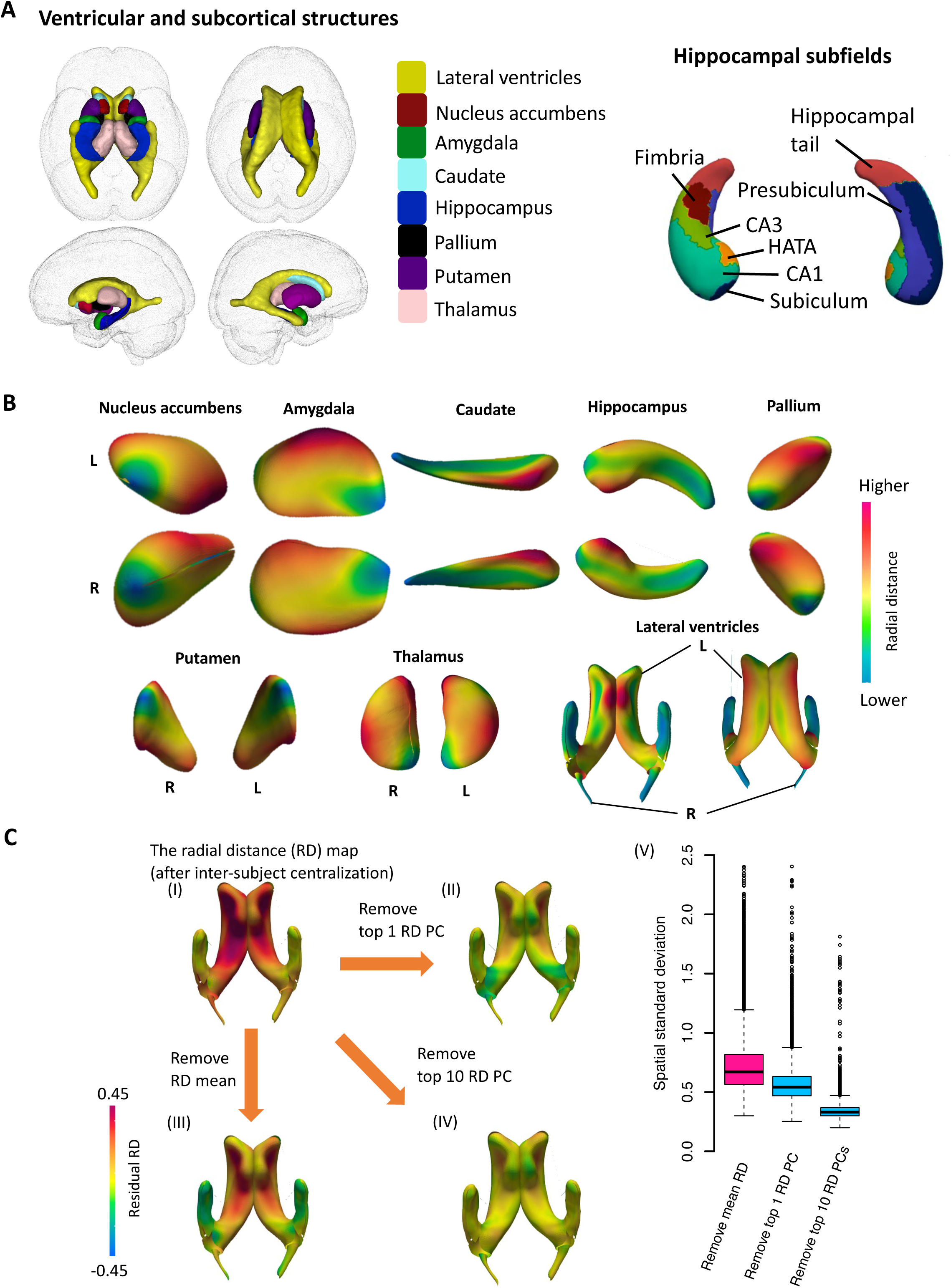
Illustrations of brain structures and their shape features. **(A)** In the left panel, we illustrate the 8 ventricular and subcortical structures, including the lateral ventricles, nucleus accumbens, amygdala, caudate, hippocampus, pallium, putamen, and thalamus. In the right panel, we present the 7 hippocampal subfields, including the cornu ammonis 1 (CA1), CA3, fimbria, hippocampus-amygdala-transition-area (HATA), hippocampal tail, presubiculum, and subiculum. **(B)** We illustrate the spatial pattern of radial distance in the vertex-wise maps of 8 ventricular and subcortical structures. These maps were generated by averaging the data from 500 randomly selected UKB subjects. See Figure S2 for additional maps of other 6 shape statistics (mTBM1, mTBM2, mTBM3, determinant, eigenvalue1, and eigenvalue2). L, left; R, right. **(C)** Comparison between the mean radial distance (RD) and structure-specific RD principal components (PCs) on the lateral ventricles. (I) illustrates an example vertex-wise RD map within the lateral ventricles after inter-subject centralization; (II) shows the residual RD map after removing the within-subject mean RD; In (III) and (IV), instead of removing the within-subject mean as in (II), we removed the top one and 10 RD PCs, respectively. (V) illustrates the standard deviation across the vertices in residual RD map for each subject in the UKB (*n* = 32746). Comparing (II) with (III), the top one PC can capture more spatial variations than the mean RD and thus reduce the standard deviations of residuals in (V). (V) also shows that the standard deviations are further reduced after removing the top 10 RD PCs (in (IV)), suggesting that additional PCs can account for more local spatial variations that are ignored by the mean RD (in (III)) or top one RD PC (in (II)).

After extracting vertex-wise maps, we aggregated them and generated region-specific summary-level features for downstream genetic analyses. For each shape statistics, we have two groups of features. The first group includes 210 structure-averaged shape features in regions or subfields by taking the mean across all the vertices within the structure (7 shape measurements × (8 shape structures + 7 hippocampal subfields) × 2 hemispheres). In the second group, we applied principal component analysis to extract 1,120 region-specific principal components (PCs) by taking the top 10 PCs of the vertex-wise map for each of the 7 statistics in the 8 ventricular and subcortical structures (left/right, 7 × 8 × 2 × 10) (**Methods**). Principal component analysis is a well-established method for dimension reduction with a wide range of neuroimaging applications. In shape analysis, the top-ranked PCs can characterize the strongest variation components of shape statistics within each structure, which can provide more microstructural details about shape deformations omitted by structure-averaged measures, while alleviating multiple testing burdens (Fig.1C). Clinically, variations represented by these PCs may localize shape changes that are more relevant to specific brain-related complex traits or diseases.

We evaluated the intra-subject reproducibility of the above region or subfield-specific shape features using the repeat scans from the UKB repeat imaging visit (average time between visits = 2 years, average *n* = 2,788). Specifically, we quantified individual-level differences between the two visits by calculating the intraclass correlation coefficient (ICC) of each shape feature between two observations from all revisited individuals. The average ICC was 0.443 (standard error = 0.234) across the 1,330 (210 + 1,120) shape features (**Table S1**). There were 457 shape features with ICC > 0.5, including 117 features for lateral ventricles, 246 for subcortical structures, and 94 for hippocampal subfields (**Fig. S3A**, mean ICC = 0.722, standard error = 0.132). The average ICC of the 7 shape statistics ranged from 0.684 to 0.752 (**Fig. S3B**), and the lateral ventricles had the highest mean ICC across all the 8 structures (**Fig. S3C,** mean ICC = 0.816, standard error = 0.129). Our later genetic analyses focused primarily on these 457 reproducible shapes features (ICC > 0.5, 363 region-level and 94 subfield-level traits) (**Table S2**).

### Heritability and associated genetic loci of ventricular and subcortical shape features

We estimated SNP heritability (*h^2^*) for these 457 reproducible shapes features using the UKB individuals of white British ancestry via GCTA^44^ (average *n* = 32,631, phases 1-3 release). Most heritability estimates (456/457) were significant after adjusting for multiple comparisons using the Benjamini-Hochberg procedure to control the false discovery rate (FDR) at 0.05 level (**Fig. S4A** and **Table S3**). The mean heritability ranged from 22.1% to 27.3% (standard error = 1.85%) across the 7 groups of shape statistics, suggesting different shape deformation measures were under comparable genetic controls (**Fig. S4B**). The highest heritability reported in each structure ranged from 51.2% (for lateral ventricles) to 19.3% (for amygdala) (**Fig. S2C**). On average, lateral ventricles had higher heritability than that of subcortical structures (32.2% vs. 21.6%, *P* < 2.2 × 10^-16^). Subfield analysis provided more information for genetic influences on different parts of the hippocampus. For example, we found the CA3 subfield had the highest heritability among the 7 subfields, while the lowest heritability was observed on the HATA subfield (**Fig. S4D**). In addition, the heritability estimates were largely consistent in females and males (**Fig. S5**, correlation = 0.83).

It is known that the volumetric measures of these structures were also heritable^26–28^. To quantify the genetic effects additionally contributing on shape measures, we compared the estimates of genetic variance for 363 region-level shape features before and after adjusting for their corresponding regional volumes as covariates (**Table S4**). For the 129 mean and first PC (PC1) shape features, we found that the average genetic variance reduced from 0.241 to 0.145, indicating that 39.8% (0.096/0.241) genetic variations were shared by regional brain volumes and shape features (Fig. 2A). In this group of shape features, the proportion varied greatly from region to region. For example, the largest proportion was observed in the lateral ventricles (79.2%) and the least was observed in the amygdala (3.4%). As both mean and PC1 features captured the major variations in the brain region^45^, these results quantified the overlapping genetic influences between shape and volumetric measures. On the other hand, for other PCs (other than the PC1s), the genetic variance estimates were much more consistent before and after adjusting for regional volumes (Fig. 2B). It was common for these PCs to capture more local variations that are not captured by the mean or PC1s^45^. Specifically, the average proportion of reduction was 11.9% and the reductions were small for the majority of shape features. There was a substantial reduction of shape features in a few second PCs (PC2) on the lateral ventricles (70.9%), which can be explained by the fact that the ventricles were large and therefore the second PC still mainly reflected global variations. Overall, these results suggest that local PCs of shape features can detect genetic influences which are largely independent of those found in regional volumes or aggregated shape measures.

**Fig. 2.**
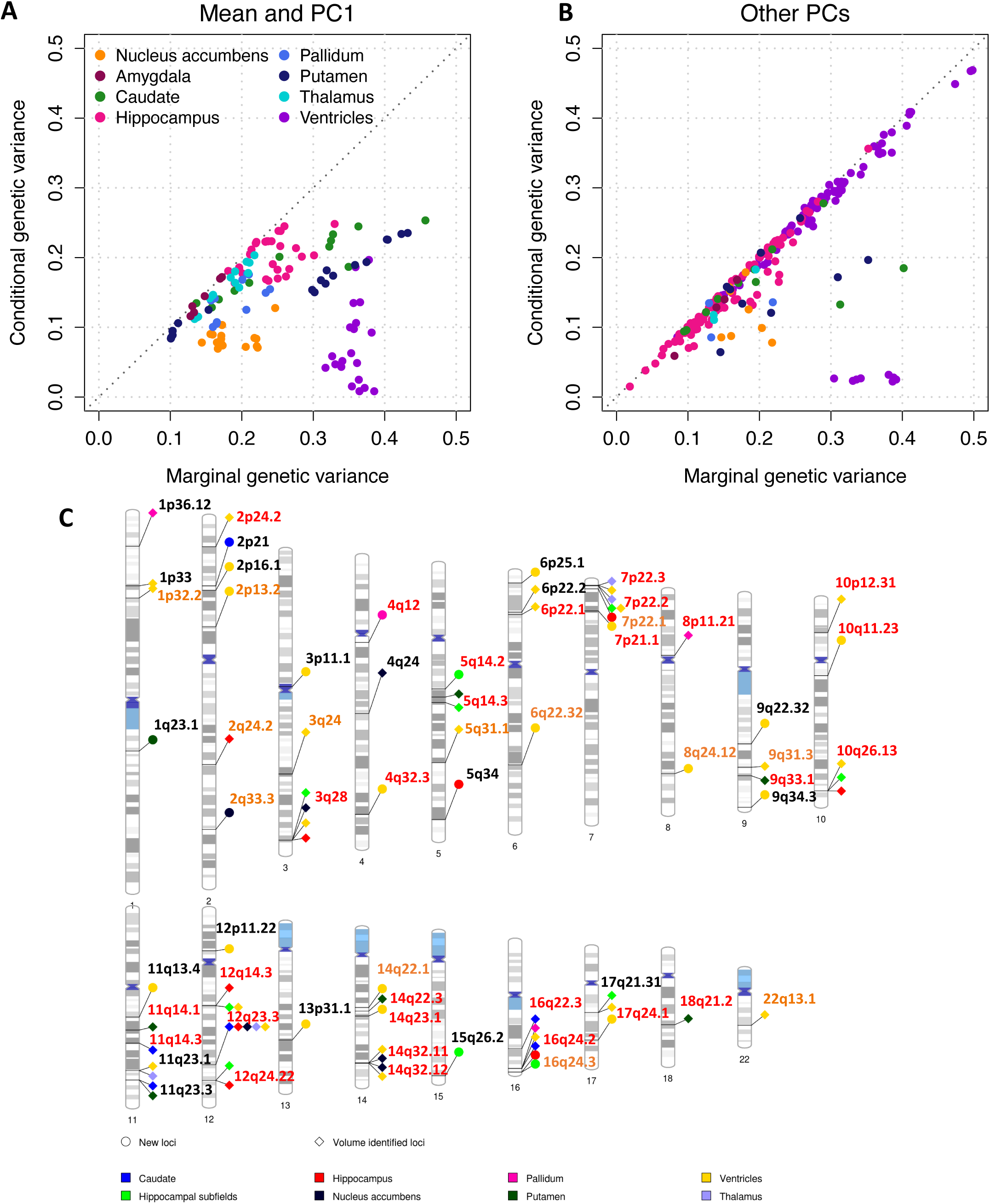
Genetic variance and the associated genomic regions of shape features. **(A-B)** The dots represent the genetic variance estimates of shape features. We compare the original (marginal) genetic variance estimates before adjusting for corresponding volumetric measurements (*x* axis) and the conditional genetic variance estimates after adjusting for volumes (*y* axis). The results for the mean values and top one PCs (PC1s) are displayed in the left panel (**A**), and results for the other PCs are displayed in the right panel (**B**). We show the significant estimates after controlling the false discovery rate of multiple testing at 5% level. Based on these results, we find that volumes can partially capture the genetic influences on PC1s and mean features (in A), while they cannot capture the majority of genetic influences on other PCs (in B). **(C)** Ideogram of 60 genomic regions influencing shape features (*P* < 1.09 × 10^-10^), 19 of which were not identified by the corresponding volumetric measurements. The colors of dots represent the different structures (and hippocampal subfields). Each signal dot indicates that at least one of the shape features of this brain structure is associated with the genomic region. The name of genomic regions replicated in more than one validation datasets or in one validation dataset at the nominal significance level were highlighted in red and brown labels, respectively.

We performed GWAS for the 457 shape features using the UKB individuals of white British ancestry (average *n* = 32,631, **Methods**). The average intercept in linkage disequilibrium score regression (LDSC)^46^ was 1.0072 (range = (0.982, 1.034)), indicating no genomic inflation of summary statistics because of confounding factors. At a stringent significance level 1.09 × 10^-10^ (5 × 10^-8^/457, additionally adjusted for the number of shape features), 622 independent (linkage disequilibrium [LD] *r*^2^ < 0.1) significant shape-variant associations^47^ were identified, which were distributed across 60 genomic regions (cytogenetic bands). There were 38 regions associated with the lateral ventricles, 10 with hippocampal subfields, 9 with hippocampus, 7 with putamen, 6 with nucleus accumbens, 6 with caudate, 4 with pallidum, and 4 with thalamus (Fig. 2C). **Table S5** summarizes the list of index genetic variants and their associated shape features. Among the 60 regions, 19 were not identified by ventricular and subcortical regional volumes (at the 5 × 10^-8^/30 significance level) in the same dataset. The genetic effects were highly consistent between males and females in the sex-specific GWAS (**Fig. S6**, correlation = 0.966), where analysis was conducted for females and males separately.

Using 5 independent European and non-European datasets, we replicated the genomic loci identified in our discovery GWAS. First, we performed GWAS on a UKB European dataset, which includes European individuals in the new UKB phase 4 data (early 2021 release) and individuals of white but non-British ancestry in the UKB phases 1-3 data (UKBE, removed the relatives of the discovery sample, average *n* = 4,596). Of the 622 identified independent (LD *r^2^* < 0.1) shape-variant associations, 410 (65.9%) from 37 (61.7%, 37/60) loci passed the 0.05 nominal significance level in UKBE. Their genetic effects all had concordant directions in the UKBE and original discovery GWAS and were highly similar (correlation = 0.980). Second, we repeated GWAS on a validation dataset with UKB non-European subjects (UKBNE, average *n* = 1,224). In this dataset, 19 loci can be validated, and most of their associations (83/86) had the same directions with those in the discovery GWAS and UKBNE. The validated genetic effects were highly consistent between the white British discovery GWAS and non-European GWAS (correlation = 0.931). These results suggest similar genetic effects on subjects from different ancestries in the same cohort.

Next, we carried out GWAS on 3 ABCD validation datasets: the ABCD European (ABCDE, average *n* = 3,177), ABCD Hispanic (ABCDH, average *n* = 662), and ABCD Black (ABCDB, average *n* = 1,002). In ABCDE, 23 of the 60 genomic loci were significant at nominal significance level and had the same effect direction as in the UKB discovery GWAS. The ABCDH and ABCDB had 13 and 15 validated loci, respectively. Interestingly, we found that the genetic effects of 3q28 locus on lateral ventricles were much larger in the three ABCD datasets than those of the UKB discovery sample, especially for the non-European subjects in ABCDH (mean absolute genetic effects 0.082 vs. 0.3367, *P* = 1.35 × 10^-7^) and ABCDB (mean absolute genetic effects 0.082 vs. 0.401, *P* = 1.67 × 10^-4^). The 3q28 locus was reported to have the strongest associations with lateral ventricular volume^31^ and was widely associated with Alzheimer’s disease risk and biomarkers^48^. Larger 3q28 effects in ABCD may suggest that the genetic effects on lateral ventricles were stronger for younger subjects and/or non-European subjects. Overall, 43 of the 60 loci can be validated in at least one of the 5 datasets, 30 loci can be validated in more than one dataset, and 5 loci can be consistently validated in all datasets, including 3q28, 17q24.1, 14q32.11, 12q14.3, and 10q26.13. The validated loci (such as 3q28) may have higher genetic effect sizes in ABCD than in UKB discovery GWAS. These validation results were summarized in **Figure S7** and **Table S5**.

Finally, we detected gene-level associations using MAGMA^49^ and FUMA^47^. MAGMA reported 127 significant genes with 1,343 associations (*P* < 5.82 × 10^-9^, adjusted for 457 phenotypes), covering all the ventricular and subcortical structures (**Fig. S8** and **Table S6**). Among the 127 significant genes, 59 were identified by regional brain volumes^35^, 53 were observed in DTI parameters^45^, and 12 overlapped with functional MRI (fMRI) traits^50^. Eight genes were associated with all the 4 brain imaging modalities, including *FAM175B*, *FAM53B*, *METTL10*, and *RP11-12J10.3* (*METTL10-FAM53B* readthrough) in the 10q26.13 region, as well as *EPHA3* (3p11.1), *ZIC1* (3q24), *ZIC4* (3q24), and *DAAM1* (14q23.1). The *FAM175B*, *FAM53B,* and *METTL10* genes had important functions in ribosomal translation and cell regeneration^51^, and have been mapped to cocaine dependence^52^ and subjective well-being^53^. The *EPHA3* was involved in axon guidance^54^ and was highly expressed in mesenchymal subtype glioblastoma^55^. The *DAAM1* was an important part of the planar cell polarity signaling in neural development^56^ and was highly expressed in human cerebral cortex^57^. In addition, the *ZIC* genes were important components in patterning the cerebellum^58^. Overall, these 127 MAGMA-significant genes showed gene ontology enrichments^59^ in “cell morphogenesis involved in differentiation (GO:0000904)” and “T cell receptor signaling pathway (GO:0050852)” biological processes at FDR 0.05 level (*P* < 2.76 × 10^-6^). We also used FUMA^47^ to map significant variants (*P* < 1.09 × 10^-10^) to genes through a combination of their base pair location, gene expression, and 3D chromatin (Hi-C) interaction. FUMA reported 383 associated genes, 313 of which were not discovered in MAGMA (**Table S7**). These results demonstrate the polygenic genetic architecture of shape features and prioritize important genes involved in the biological pathways of brain functions and diseases.

### The shared genetic influences with complex brain traits and disorders

We took a further look into the 43 validated regions, providing variant annotations and details of shared genetic influences with other complex traits and diseases. For all the independent (LD *r^2^* < 0.1) significant variants (and variants in LD, *r*^2^ ≥ 0.6) detected in these validated regions, we searched for their GWAS signals reported in the NHGRI-EBI GWAS catalog^60^. Shape deformation of ventricular and subcortical structures has substantial regional and local genetic overlaps with complex traits and clinical endpoints. The full information was presented in **Table S8**, and below we highlighted some regions and their reported variants and genes reported for brain structures/functions, neurological disorders, psychiatric disorders, psychological traits, migraine, cognitive traits, educational attainment, sleep/physical activity, osteoarthrosis/pain, Alzheimer’s Disease biomarkers, diabetes/kidney diseases, blood traits, blood pressure, smoking/drinking, lung/liver, and lipoprotein cholesterol.

Our results were concordant with previous GWAS results for regional brain volumes and cortical thickness in many genomic loci, including 3q24 (index variant rs2279829, the nearest gene *ZIC4*), 8q24.12 (rs10283100, *ENPP2*), 9q31.3 (rs734250, *LPAR1*), 9q33.1 (rs10983205, *ASTN2*), 11q14.3 (rs1531249, *FAT3*), 11q23.1 (rs34077344, *LINC02550*), 11q23.3 (rs10892133, *DSCAML1*), 12q14.3 (rs61921502, *MSRB3*), 12q23.3 (rs12369969, *NUAK1*), 12q24.22 (rs7132910, *HRK*), 14q22.3 (rs945270, *KTN1*), and 16q22.3 (rs7193665, *ZFHX3*) (**Figs. S9-S20**). For example, rs7132910 was identified to be associated with hippocampal volume^61^. It the current study, we found rs7132910-associations with multiple shape features of the hippocampus, particularly the subiculum subfield. Additionally, rs2279829 and rs7132910 were expression quantitative trait loci (eQTLs) of *ZIC4* and *TESC* in human brain tissues^62^, suggesting that these shape-associated variants were known to affect gene expression levels in human brain. Among these regions, we also tagged genetic variants (LD *r*^2^ ≥ 0.6) reported for risk-taking^63^, alcohol consumption, smoking initiation^64^, lung function^65^, chronic obstructive pulmonary disease (COPD)^66^, blood pressure^65^, and Alzheimer’s disease pathologies^67^.

Our shape GWAS results frequently tagged regions reported for white matter microstructure, including 17q24.1 (rs62072157, *GNA13*), 2p13.2 (rs34754475, *DYSF*), 3q28 (3:190672426_CT_C, *GMNC*), 5q14.2 (rs12187334, *ATP6AP1L*), 7p21.1 (rs4329170, *TWISTNB*), 7p22.2 (rs1183079, *GNA12*), 7p22.3 (rs368699386, *AMZ1*), 14q32.12 (rs529889896, *CCDC88C*), 16q24.2 (rs56023709, *C16orf95*), and 16q24.3 (rs8404, *CDK10*) (Figs. 3A and **S21-S30**). For example, rs62072157 was associated with (the ninth PC of) the radial distance of the right lateral ventricle, and it also had significant associations with the fornix (column and body) and anterior corona radiata white matter tracts (*P* < 4 × 10^-9^). Rs62072157 was an eQTL of *GNA13* and *RGS9* in brain tissues^62^. The *GNA13* was a core gene involved in early brain development regulations^68^ and has been implicated in brain diseases such as schizophrenia^69^. In addition, rs1183079 was a brain eQTL of *AMZ1*, which was associated with mTBM2 and radial distance of the right lateral ventricle, as well as multiple white matter tracts, such as the body of corpus callosum, anterior corona radiata, retrolenticular part of internal capsule, posterior corona radiata, and superior corona radiata (*P* < 5.5 × 10^-11^). Furthermore, rs8404 was a brain eQTL of *CDK10, SPATA33, VPS9D1, MC1R,* and *ACSF3*. Rs8404 was associated with multiple shape features of the left hippocampus and affected the integrity of the retrolenticular part of internal capsule and superior longitudinal fasciculus tracts (*P* < 2 × 10^-8^). The *CDK10* was important for neural development^70^. In addition to Alzheimer’s disease biomarkers^48^ and COPD^66^, we found shared genetic influences with type 2 diabetes^71^ and blood traits (such as plasma homocysteine levels^72^ and platelet distribution width^73^) on these white matter-overlapping genomic loci.

**Fig. 3.**
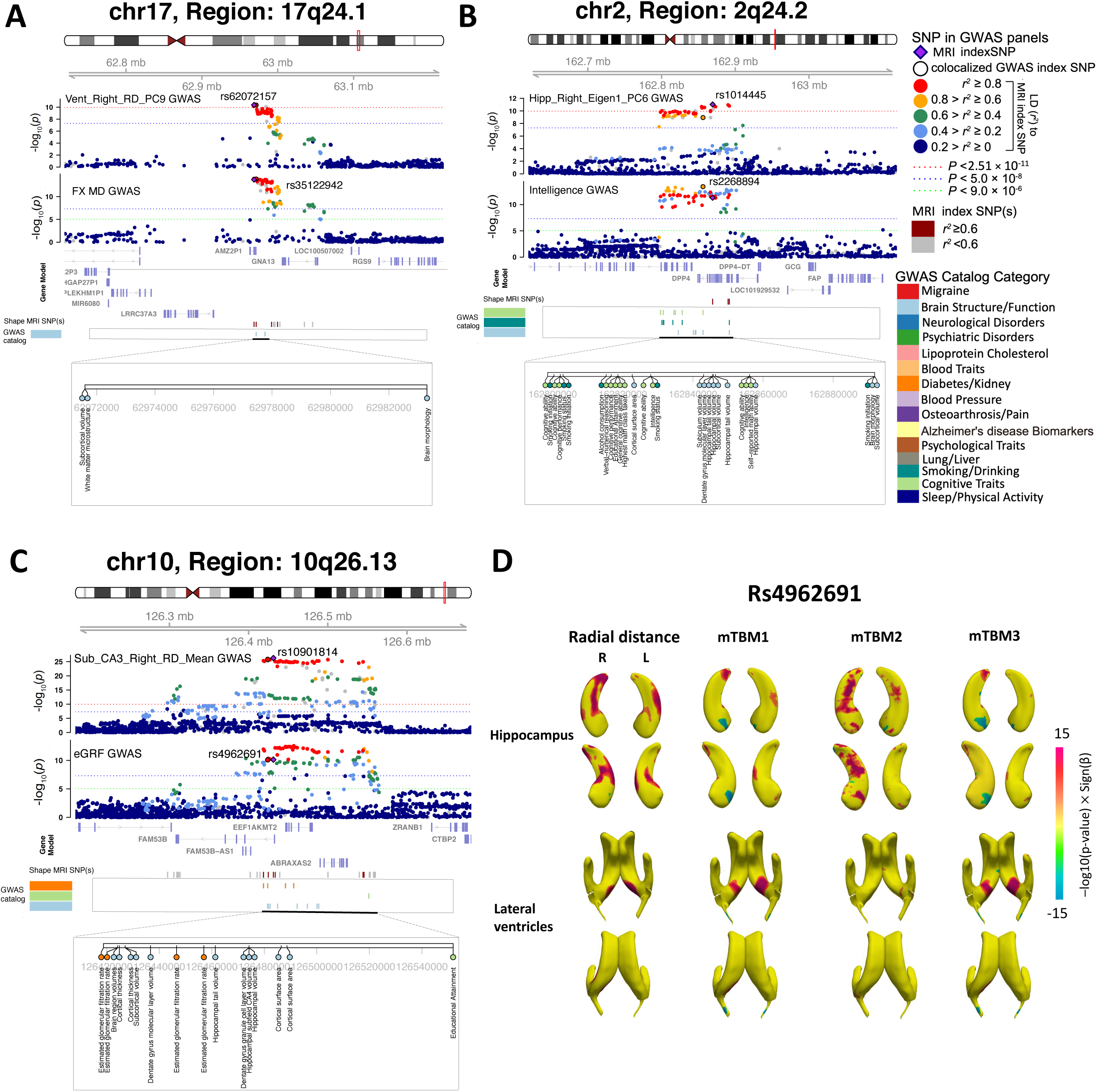
Genetic loci associated with both shape features and other complex traits. **(A)** In the 17q24.1 region, we observed shared genetic influences (LD *r*^2^ ≥ 0.6) between shape features (e.g., Vent_Right_RD_PC9, index variant rs62072157) and brain white matter microstructure (e.g., FX_MD, index variant rs35122942). Vent_Right_RD_PC9, the ninth PC of the radial distance in right lateral ventricle; FX_MD, the mean diffusivity in the fornix tract (column and body of fornix). **(B)** In the 2q24.2 region, we observed shared genetic influences (LD *r*^2^ ≥ 0.6) between shape features (e.g., Hipp_Right_Eigen1_PC6, index variant rs1014445) and cognitive traits (e.g., intelligence, index variant rs2268894). Hipp_Right_Eigen1_PC6, the sixth PC of the eigenvalue1 in right hippocampus. **(C)** In the 10q26.13 region, we observed shared genetic influences (LD *r*^2^ ≥ 0.6) between shape features (e.g., Sub_CA3_Right_RD_Mean, index variant rs10901814) and kidney function biomarker (eGRF, index variant rs4962691). Sub_CA3_Right_RD_Mean, the mean radial distance in the CA3 subfield of right hippocampus; eGRF, estimated glomerular filtration rate. In **(D)**, we illustrate the signed −log10(*P*-value) of associations between the rs4962691 variant and vertex-wise data of the hippocampus and lateral ventricles for 4 shape statistics, including radial distance, mTBM1, mTBM2, and mTBM3. It was observed that the pattern of genetic effects varied by subregion within each structure.

In 6p22.1 and 18q21.2 regions, we observed the shared genetic influences between shape features and multiple psychiatric disorders and psychological traits. For example, we tagged rs7766356 (nearest gene *ZSCAN23*, 6p22.1) and rs11665242 (*DCC*, 18q21.2), which have been implicated with schizophrenia^74, 75^ (**Figs. S31-S32**). Rs7766356 was an eQTL of *ZSCAN23, ZSCAN31, ZKSCAN3,* and *ZSCAN26,* which might represent drug targets for schizophrenia^76^. We also tagged risk variants for bipolar disorder^77^ (e.g., rs144447022) and MDD^78^ (e.g., rs926552) in these two regions and for neuroticism^79^ in 5q14.3 (e.g., rs16902900, *TMEM161B*) (**Fig. S33**). In 2q24.2, 6q22.32, 8p11.21 regions (Fig. 3B and **Figs. S34-S36**), as well as the 6p22.1 and 18q21.2 regions related to brain disorders, we found shared genetic influences with a wide range of cognitive and educational traits, such as intelligence^80^ (e.g., rs2268894, *DPP4*, 2q24.2), educational attainment^81^ (e.g., rs11759026, *CENPW*, 6q22.32; rs2974312, *SMIM19*, 8p11.21), self-reported math ability^81^ (e.g., rs71559051, *H2BC15*, 6p22.1), and verbal-numerical reasoning^82^ (e.g., rs62100026, *DCC*, 18q21.2). Finally, we tagged sleep-related variants in 10p12.31 and 11q14.1 regions, such as insomnia^83^ (e.g., rs12251016, *MLLT10*, 10p12.31; and rs667730, *DLG2*, 11q14.1, **Figs. S37-S38**). These results suggest the shape features could be used as imaging biomarkers to study the etiologic study of brain-related diseases and complex traits.

Our results also help us to better understand the genetic links between brain atrophy and the health of other organs. For example, the index rs10901814 (*FAM53B,* 10q26.13) was associated with the shape features in the hippocampus and lateral ventricles and it was a brain eQTL for *EEF1AKMT2* and *LHPP* genes. In this region, we tagged risk variant (index variant rs4962691 for estimated glomerular filtration rate (eGFR)^84^, which was a clinical biomarker for kidney function and disease (Fig. 3C). Brain and kidney had similar hemodynamic mechanisms and shared physiological links^85^. Cognitive impairment and accompanied brain structural changes (such as hippocampus volume) have been frequently reported in chronic kidney disease^86, 87^. To precisely localize the pattern of genetic effects on brain shapes, we took the eGFR lead index rs4962691 and performed vertex-wise analysis on spatial maps of hippocampus and lateral ventricles. We found that the rs4962691-related shape deformation was mainly localized to specific areas of the hippocampus and lateral ventricles, such as the hippocampal tail and CA1, as well as the atrium and posterior horn of lateral ventricles (Fig. 3D). These genetic overlaps and local structural variations may represent the mediated brain changes related to the cognitive impairment in chronic kidney disease.

### Genetic correlations with complex traits and clinical outcomes

We explored genetic correlations (GC) between shape features and a wide range of other complex traits. First, we used LDSC^88^ to examine pairwise genetic correlation between the 457 shape features and 211 brain structural traits, including 101 regional brain volumes^35^ and 110 diffusion tensor imaging (DTI) parameters^45^. Among the 96,427 (457 × 211) tests, 16.22% were significant at the FDR 5% level (**Fig. S39** and **Table S9**). Both regional brain volumes and DTI parameters had significant genetic correlations with ventricular and subcortical shape features. For DTI parameters, the strongest associations were observed on the lateral ventricles, and the top 5 associated white matter tracts included the fornix, body of corpus callosum, superior corona radiata, posterior corona radiata, and posterior limb of internal capsule (*P* < 1.49 × 10^-55^). Meanwhile, either the fornix or body of corpus callosum tracts consistently had the strongest associations with the subcortical structures, such as the hippocampus (*P* = 9.67 × 10^-25^), nucleus accumbens (*P* = 2.18 × 10^-23^), amygdala (*P* = 4.51 × 10^-8^), caudate (*P* = 1.32 × 10^-19^), pallidum (*P* = 9.36 × 10^-16^), putamen (*P* = 2.73 × 10^-15^), and thalamus (*P* = 8.95 × 10^-38^). Our subfield analysis further revealed that the fornix and body of corpus callosum associations were mainly localized in the presubiculum subfield of hippocampus (*P* < 3.68 × 10^-14^). The fornix located below the corpus callosum and connected the hippocampus to subcortical structures^89^. Our results suggested that the fornix integrity and shape deformations had strong genetic overlaps. For regional brain volumes, we found associations for volumes of both cortical and subcortical structures. As expected, the ventricular and subcortical structures had strong genetic correlations with subcortical volumes. In addition, genetic correlations were also widely observed between cortical structures and ventricular and subcortical shapes. Top-ranked cortical volumes included the left/right insula (with putamen, *P* < 7.61 × 10^-18^), left/right isthmus cingulate (with ventricle and hippocampus presubiculum, *P* < 9.77 × 10^-10^), left/right lingual (with ventricle, *P* < 2.70 × 10^-14^), left/right pericalcarine (with ventricle, *P* < 3.41 × 10^-12^), and left/right cuneus (with ventricle, *P* < 2.37 × 10^-10^). Overall, these results suggest that ventricular and subcortical shape features are genetically related to white matter integrity and structural variations of cortical regions.

Next, we examined genetic correlations between the 457 shape features and 48 complex traits and diseases. At the FDR 5% level (457 × 48 tests), we found the shape features were associated with brain disorders (such as attention-deficit/hyperactivity disorder (ADHD), schizophrenia, and anorexia nervosa), cognitive traits (such as cognitive function, intelligence, and reaction time), sleep traits (such as snoring, insomnia, extreme chronotype), neuroticism, risk-taking, metabolic traits, and cardiovascular diseases (such as hypertension and coronary artery disease (CAD)) (**Fig. S40** and **Table S10**). For example, ADHD was positively correlated with shape features of the left hippocampus and lateral ventricles (|GC| > 0.184, *P* < 5.69 × 10^-4^, Figs. 4A-4B). In ADHD, there have been reports of abnormalities in hippocampal structures, possibly as a result of the brain’s efforts to compensate for disruptions of time perception and a tendency to avoid waiting^90^. The Rotterdam Study reported that ADHD-related genetic variants were associated with structural brain changes in the lateral ventricles^91^. In addition, schizophrenia was genetically associated with shape features of the thalamus and hippocampus (|GC| > 0.133, *P* < 6.14 × 10^-4^). Both the thalamus and hippocampus had crucial roles in functional and structural pathways related to schizophrenia^92^ and smaller volumes in the two structures were frequently reported in schizophrenia patients^93, 94^. An earlier study using subcortical brain volumes was not able to detect genetic overlap between schizophrenia risk and subcortical structures^95^. Furthermore, most significant genetic correlations with cognitive function, intelligence, and education were with the lateral ventricles (|GC| > 0.113, *P* < 5.42 × 10^-4^), while the reaction time additionally had genetic correlations with the thalamus (|GC| > 0.115, *P* < 7.90 × 10^-4^, Fig. 4C). Ventricular enlargement was strongly correlated with cognitive performance decline^96^. The thalamus passed information between the brain and body and anticipatory thalamic activity can predict reaction time^97^. We also observed specific genetic correlations for other traits, such as between insomnia and the caudate (|GC| > 0.141, *P* < 7.85 × 10^-4^), risk tolerance and the hippocampus (|GC| > 0.283, *P* < 8.15 × 10^-4^), and automobile speeding and the lateral ventricles (|GC| > 0.148, *P* < 3.52 × 10^-4^). Furthermore, we found that CAD was genetically correlated with the hippocampus and lateral ventricles (|GC| > 0.260, *P* < 6.40 × 10^-4^, Fig. 4D). CAD had long-term negative impact on brain health and reduced neural connectivity changes in the hippocampus were observed in CAD and may contribute to cognitive impairment^98^. A significant genetic correlation between AD and the hippocampus was not observed.

**Fig. 4.**
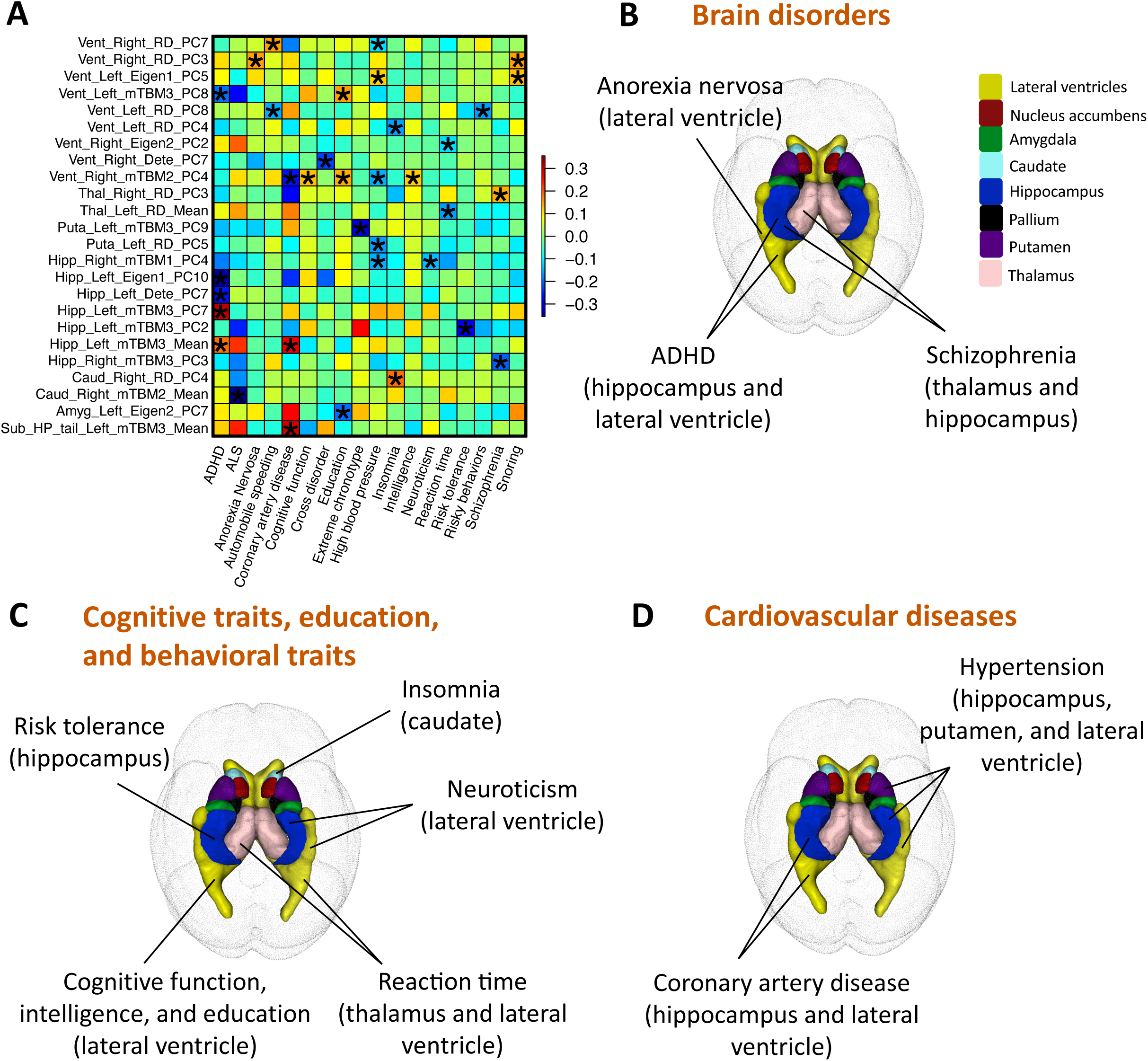
Selected genetic correlations with complex traits and diseases. **(A)** We illustrate pairwise genetic correlations between shape features (*x* axis) and other complex traits and diseases (*y* axis) estimated by LDSC. The asterisks highlight significant pairs after controlling the FDR at 5% level. The colors represent the genetic correlation estimates. See Table S1 for descriptions of the shape features. **(B)** We illustrate brain structures whose shape features are genetically related to brain disorders, such as schizophrenia, attention-deficit/hyperactivity disorder (ADHD), and anorexia nervosa. **(C)** We illustrate brain structures whose shape features are genetically related to cognitive traits, education, and behavioral traits. **(D)** We illustrate brain structures whose shape features are genetically related to cardiovascular diseases, such as coronary artery disease and hypertension.

### Phenome-wide association study using shape polygenic risk scores

We tested associations between the 457 shape features and more complex traits and clinical outcomes using polygenic risk scores (PRS) of shape features in the UKB non-imaging cohort (**Methods**). A total of 276 complex traits and clinical outcomes were selected from a variety of categories (**Table S11**). Briefly, we constructed PRS using PRS-CS^99^ for shape features on unrelated UKB subjects without brain MRIs (*n* = 379,860, also removing relatives of imaging subjects). We then focused on the UKB white British subjects and randomly selected 70% individuals (average *n* = 202,405) as discovery sample to test pairwise associations (457 × 276 tests) and validated these results in a hold-out dataset consisting of the rest of 30% white British subjects (average *n* = 86,736), UKB white non-British subjects (average *n* = 20,746), and UKB non-white subjects (average *n* = 21,587). Detailed information on the adjusted covariates can be found in the **Methods** section. We prioritized significant associations in the validation sample (at the nominal significance level) with concordant regression coefficients when they passed the Bonferroni significance level in the discovery sample. Among the 457 × 276 tests, 2,907 were significant at Bonferroni significance level in the discovery sample, among which 94.60% (2,750) were validated (**Fig. S41** and **Table S12**). We highlighted below the association patterns with clinical outcomes, mental health, cognitive function, physical activity, lifestyle, and biomarkers.

We observed significant associations between shape PRS and multiple diseases, including diabetes, hyperthyroidism, hypothyroidism, multiple sclerosis, psoriasis, and vascular heart problems. For example, diabetes was significantly associated with PRS for multiple shape features of the left hippocampus and its subfields (|*β* | > 0.0109, *P* < 2.47 × 10^-7^). These findings were consistent with previous studies showing patients with long diabetes had higher risk of hippocampal atrophy, and loss of hippocampal neuroplasticity and neurogenesis^100, 101^. In addition, both hyperthyroidism and hypothyroidism were mostly associated with shape PRS of the lateral ventricles and hippocampus (and hippocampal subfields) (|*β* | > 0.0107, *P* < 3.87 × 10^-7^). Adults with hypothyroidism were reported to have decreased hippocampal volume^102^ and people with hyperthyroidism had smaller grey matter volume in bilateral hippocampus^103^. Hyperthyroidism and hypothyroidism had also been linked to changes of brain ventricle size^104^. Multiple sclerosis was mostly associated with shape features of the putamen and hippocampus (|*β* | > 0.0108, *P* < 3.42 × 10^-7^). Decreased putamen and hippocampal volumes have been reported in multiple sclerosis patients^105, 106^. We also found associations between vascular heart problems and shape PRS of the hippocampus (|*β* | > 0.0105, *P* < 3.71 × 10^-7^), consistent with previous studies showing decreased hippocampal volume among patients with vascular heart problems^107^.

There were significant associations between shape PRS and multiple mental health traits related to anxiety, depression, and neuroticism (|*β* | > 0.0106, *P* < 1.09 × 10^-11^). Most of these mental health traits were associated with shape features of the hippocampus and its subfields. These observed associations were consistent with recent findings on reduced hippocampal volume in subiculum^108^ and fimbria^109^ among patients with MDD. Significant associations were also found between mental health and shape PRS of other brain structures, such as between nervous feelings and the nucleus accumbens, neuroticism and the putamen, as well as tiredness/lethargy and the caudate.

The blood biochemistry biomarkers were widely associated with the shape PRS of all structures, with the majority of associations involving the hippocampus, hippocampal subfields, and lateral ventricles. Examples of associated biomarkers included apolipoprotein A, aspartate aminotransferase, glycated haemoglobin (HbA1c), high-density lipoprotein (HDL) cholesterol, insulin-like growth factor 1 (IGF-1), total protein, and urate (|*β* | > 0.009, *P* < 4.28 × 10^-42^). It is known that urate caused hippocampal infection, which in turn induced cognitive dysfunction^110^. HbA1c measured the blood sugar level and was used in diagnosis of diabetes. Our results were consistent with a recent study that higher level of HbA1c was associated with smaller hippocampal volume^111^. In summary, shape PRS uncovered the links between shape features and a wide range of complex traits and diseases. As the shape PRS were genetically predicted traits, these observed associations also indicate the widespread underlying shared genetic influences.

### Causal relationships with clinical endpoints detected by Mendelian randomization

To explore the causes and consequences of shape deformations, Mendelian randomization (MR) was used to identify potential causal relationships between the 457 shape features and 288 clinical endpoints collected by FinnGen^112^ and the Psychiatric Genomics Consortium^113^ (**Table S13**). We tested 14 different MR methods^114–117^, and the detailed implementation information can be found in the **Methods** section.

At the Bonferroni significance level (*P* < 3.92 × 10^-8^), we found strong evidence of genetic causal effects from shape features to brain disorders, including AD, schizophrenia, and cross disorders (five major psychiatric disorders^118^) (Fig. 5A and **Table S14**). For example, multiple ventricular shape statistics had significant genetic causal effects on Alzheimer’s disease (|*β* | > 0.474, *P* < 2.73 × 10^-8^), most of which were from the left lateral ventricle. AD can affect gray and white matter structures surrounding the ventricles, and ventricular enlargement and expansion have been frequently identified in AD^9,119–121^. Our findings provide further evidence for the causal genetic pathway underlying brain structural variations and Alzheimer’s disease. There was a consistent sign of causal genetic effects across different MR methods. In addition, shape features of the putamen and hippocampus were causally linked to schizophrenia (|*β*| > 0.339, *P* < 1.97 × 10^-8^). In schizophrenia, neuropsychological impairments have been associated with hippocampal structures^94^. It is well known that the putamen is associated with both increased dopamine synthesis capacity and frontostriatal dysconnectivity in schizophrenia, as well as with antipsychotic treatment effects^122^. Additionally, we did not detect causal genetic links to schizophrenia by analyzing the volumes of putamen and hippocampus structures (|*β*| < 0.293, *P* > 0.0011). Furthermore, there were significant genetic causal relationships between shape features and other diseases of the nervous system, such as carpal tunnel syndrome (|*β* | > 0.249, *P* < 1.26 × 10^-8^) and migraine (|*β* | > 0.256, *P* < 7.61 × 10^-14^). All of the above results passed the MR-Egger intercept test, indicating the absence of horizontal pleiotropy.

**Fig. 5.**
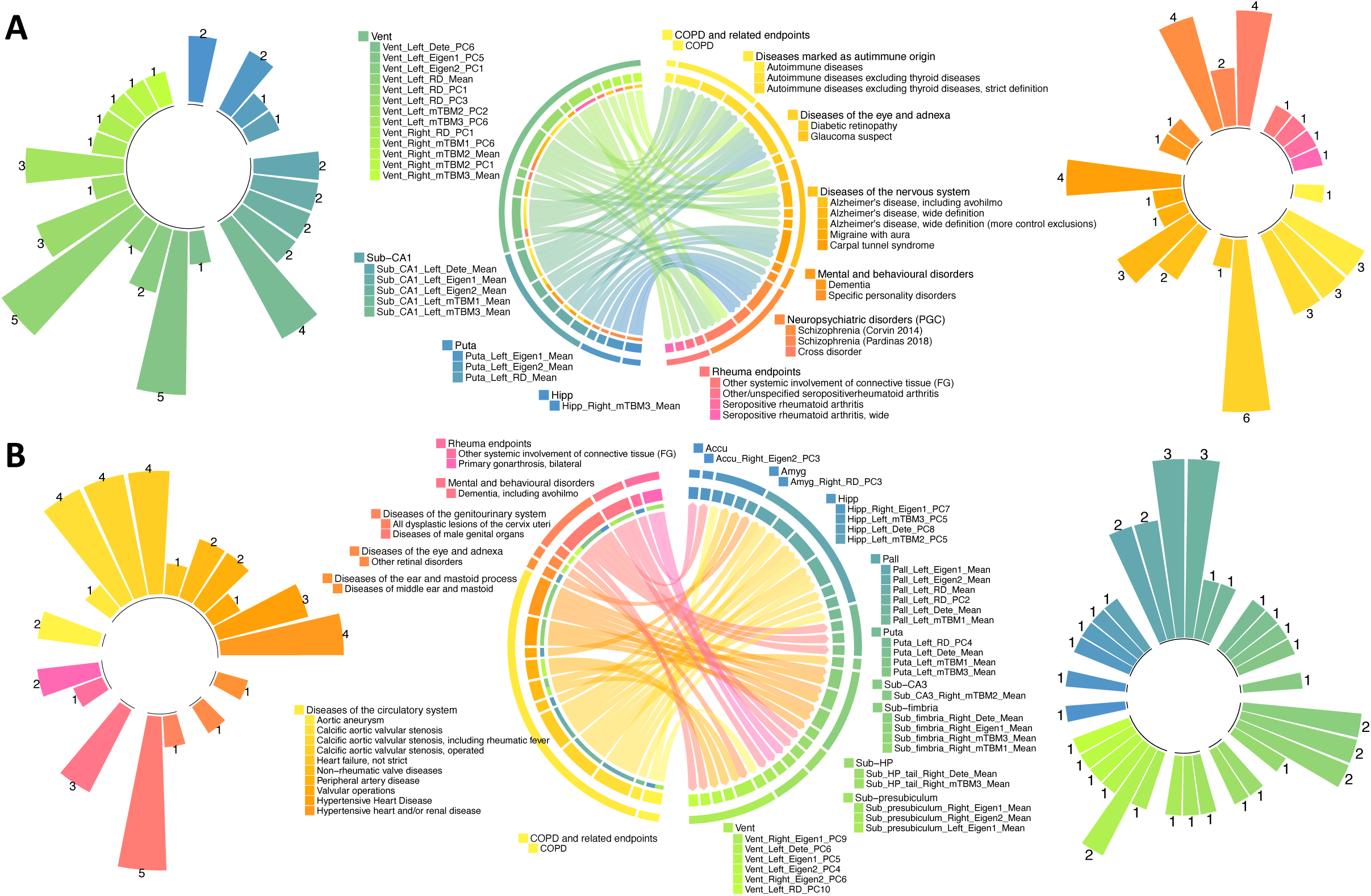
Mendelian randomization analysis with clinical outcomes. **(A)** Significant causal genetic links from brain shape features to clinical endpoints (*P* < 3.92 × 10^-8^). **(B)** Significant causal genetic links from clinical endpoints to brain shape features (*P* < 1.12×10^-8^). In both A and B, the chord plot in the middle display each causal pair. The first level (out) circle indicates each disease or brain structure. The second level circle indicates the specific diseases with each disease category, or shape features within each brain structure. The third level circle on the shape feature side indicates which diseases the shape feature links to. In addition, the circle plots on the left- and right-hand sides display the number of significant pairs for each exposure and outcome variable, respectively. FG, FinnGen; PGC, Psychiatric Genomics Consortium; COPD, chronic obstructive pulmonary disease; Sub, subfield of hippocampus; HP, hippocampal tail; and CA, cornu ammonis. See Table S1 for descriptions of the shape features.

Additionally, we identified causal relationships where sleep traits were the exposure and brain structural traits were the outcome at Bonferroni significance level (*P* < 1.12×10^-8^). A large proportion of the significant findings (26/42) were related to the diseases of the circulatory system, such as aortic aneurysm, calcific aortic valvular stenosis, heart failure, and hypertensive heart disease (Fig. 5B and **Table S14**). For example, calcific aortic valvular stenosis had causal genetic links to the shape of the pallidum (|*β* | > 0.059, *P* < 1.02 × 10^-8^), and hypertensive heart disease may lead to the changes of the fimbria subfield (|*β* | > 0.075, *P* < 4.28 × 10^-9^). Multiple brain disorders, such as stroke^123^ and dementia^124^, can be caused by diseases of the circulatory system. We also found that COPD was causally related to the shape of hippocampus and lateral ventricles (|*β* | > 0.064, *P* < 9.87 × 10^-9^). Cognitive impairments and related alterations of hippocampus had been found in COPD patients^125^. Overall, our MR results suggest that genetic causal pathways may exist between brain shapes and brain disorders, such as Alzheimer’s disease. These findings also reveal possible genetic mechanisms of non-brain diseases (e.g., heart diseases) that underlie brain health.

## DISCUSSION

Brain volumes are commonly used to analyze brain ventricular and subcortical structures. Increasing evidence, however, indicates that volumetric measurements can only partially capture structural complexity. As a result, shape analysis has been performed to uncover deformations that are not visible in volumetric analysis. Using over 45,000 MRI scans in the UKB and ABCD studies, our study uncovered the genetic architecture of shape features within the ventricular and subcortical regions. We identified genetic connections between shape features and a wide range of complex traits and clinical outcomes. Leveraging shape features identified new loci that could not be identified by brain volumes and provided fine details for localizing the genetic effect patterns within brain structures. The results of our study improved the spatial resolution for identifying genetically important brain areas that influenced clinical outcomes. For example, rs4962691, a risk variant for eGFR, had genetic effects in specific parts of the hippocampus and lateral ventricles. These shape features may be used as endophenotypes of the cognitive impairment in kidney disease. In summary, as one of the first large-scale studies to examine the genetic architecture of ventricular and subcortical shape features, our results provide specific shape biomarkers that can be used in clinical research questions.

The present study has a few limitations. First, the current analysis mainly used data from the European UKB subjects, which may limit the generalizability of our research findings. In the validation analysis, we have observed that European-significant genetic variants tended to have larger effects in non-European cohorts of the ABCD study. The inclusion of more global samples and identification of cross-population components of genetic effects on brain shape will be of great interest in future studies. Second, we used PCA to extract low-rank features from vertex-wise maps, which can capture local shape deformations while mitigating multiple testing burden for genome-wise testing. PCA is a powerful statistical tool to extract linear structures in the data. Nevertheless, PCA may not be the most efficient method for reducing dimensions because complicated shape variations can be non-linear. It might be possible to generate more powerful shape features using variational autoencoder^126^ and transfer learning^127^. Lastly, we studied hippocampal subfields, which may reflect specific biological processes and cognitive functions^128, 129^. Other subcortical structures (such as amygdala^130^) and lateral ventricles^12^ can also be divided into different portions or subregions with distinct functions. The development and application of automated segmentation methods in large-scale MRI datasets will enable the discovery of genetic influences at the subfield level for more brain structures.

## Data Availability

The individual-level data used in the present study can be obtained from the UK Biobank (https://www.ukbiobank.ac.uk/) and ABCD (https://abcdstudy.org/) studies. Our GWAS summary statistics will be shared on Zenodo and at the BIG-KP https://bigkp.org/. Our GWAS results will also be available via the interactive web browser at http://165.227.78.169:443/.

## METHODS

Methods are available in the ***Methods*** section.

Note: One supplementary information pdf file, one supplementary figure pdf file, and one supplementary table zip file are available.

## ACKNOWLEDGEMENTS

This research was partially supported by U.S. NIH grants MH086633 (HT.Z.), MH116527 (TF.L.), and U01HG011720 (Y.L.). We thank the individuals represented in the UK Biobank and ABCD studies for their participation and the research teams for their work in collecting, processing and disseminating these datasets for analysis. We thank Doug Crabill for helpful conversations on computing. We would like to thank the University of North Carolina at Chapel Hill and Purdue University and their research computing group for providing computational resources and support that have contributed to these research results. We gratefully acknowledge all the studies and databases that made GWAS summary data available. This research has been conducted using the UK Biobank resource (application number 22783), subject to a data transfer agreement. Part of the data used in the preparation of this article were obtained from the Adolescent Brain Cognitive Development (ABCD) Study (https://abcdstudy.org), held in the NIMH Data Archive (NDA). This is a multisite, longitudinal study designed to recruit more than 10,000 children age 9-10 and follow them over 10 years into early adulthood. The ABCD Study is supported by the National Institutes of Health and additional federal partners under award numbers U01DA041022, U01DA041028, U01DA041048, U01DA041089, U01DA041106, U01DA041117, U01DA041120, U01DA041134, U01DA041148, U01DA041156, U01DA041174, U24DA041123, U24DA041147, U01DA041093, and U01DA041025. A full list of supporters is available at https://abcdstudy.org/federal-partners.html. A listing of participating sites and a complete listing of the study investigators can be found at https://abcdstudy.org/scientists/workgroups/. ABCD consortium investigators designed and implemented the study and/or provided data but did not necessarily participate in analysis or writing of this report. This manuscript reflects the views of the authors and may not reflect the opinions or views of the NIH or ABCD consortium investigators. Assistance for this project was provided by the UNC Intellectual and Developmental Disabilities Research Center (NICHD; P50 HD103573).

## AUTHOR CONTRIBUTIONS

B.Z. and H.Z. designed the study. B.Z., TF.L., X.Y., J.S., X.W., TY. L, Y.Y., Z.W., Z.F., and Z. J. analyzed the data. TF. L., X.W., TY. L, Y.Y., J.C., Y.S., J.T., D.X, Z.Z., M.G., W.G., and C.T. processed the MRI data. Y.W. and Q.D. helped on the shape feature pipelines. Y.L. and J.L.S. provided feedback on study design and results interpretations. B.Z. wrote the manuscript with feedback from all authors.

**CORRESPINDENCE AND REQUESTS FOR MATERIALS** should be addressed to H.Z.

## COMPETETING FINANCIAL INTERESTS

The authors declare no competing financial interests.

## METHODS

### Shape features and imaging datasets

The raw structural MRI data from the UKB and ABCD studies were used in this study. The UKB study’s ethics approval was obtained from the North West Multicentre Research Ethics Committee (approval number: 11/NW/0382). The procedures of the ABCD study were approved by the institutional review boards at ABCD collection sites (approval numbers: 201708123 and 160091). The image collection and processing procedures can be found in Alfaro-Almagro, et al. ^131^ for the UKB study and Casey, et al. ^132^ for the ABCD study.

The shape feature generation pipeline was detailed in the **Supplementary Note** and an overview of the procedures and examples were virtualized in **Figures S1 and S42**. Briefly, we focused on 8 ventricular and subcortical structures, including the left/right lateral ventricles, nucleus accumbens, amygdala, caudate, hippocampus, pallidum, putamen, and thalamus. We constructed the vertex-wise map for 7 different shape statistics, including radial distance, mTBM1, mTBM2, mTBM3, determinant, eigenvalue1, and eigenvalue2. For each shape statistics, we aggregated these vertex-wise data by 1) taking the mean across all vertices for each structure; and 2) generating the top-ranked PCs for each structure (left and right hemispheres separately). Intuitively, the purpose of these PCs was to capture global and local variations within the vertex-wise representation of brain structures. Typically, the first one or two PCs represented the global patterns, which were similar to the mean values. Other PCs, however, captured local variations that mean or top-ranked PCs missed. See Figure 1C for an illustration. Additionally, we segmented the hippocampus and calculated the mean of shape statistics for each of the following subfields: the left/right CA1, CA3, fimbria, HATA, hippocampal tail, presubiculum, and subiculum. In total, we had 1,330 (210 mean values and 1,120 PCs) shape features. Based on the UKB revisit data, we calculated the reproducibility (ICC) and selected those with ICC > 0.5, resulting in a final set of 457 shape features (363 at the structure-level and 94 at the subfield-level) for genetic analysis. See **Table S1** for their names and descriptions.

We analyzed the above 457 reproducible traits in the following datasets: 1) the white British discovery dataset, where the data were from white British subjects in UKB phases 1 to 3 imaging data (average *n* = 32,631, released up through 2020); 2) the UKB European validation dataset, which included White individuals in the newly released UKB phase 4 data and the UKB non-British white individuals in phases 1 to 3 data (UKBE, removed the relatives of the discovery sample, average *n* = 4,596); 3) the UKB non-European validation dataset that consisting of non-White subjects in the UKB phases 1 to 4 data (UKBNE, average *n* = 1,224); 4) the UKB first revisit dataset (average *n* = 2,788); and 5) the ABCD dataset (average *n* = 8,496). The average age (at imaging) of all UKB subjects was 64.2 (standard error = 7.73), 51.6% were females; the average age for all ABCD students was 9.93 (standard error = 0.62), 48.2% were females. Self-reported ethnicity (Data-Field 21000) was used to assign ancestry in UKB, whose accuracy was verified in Bycroft, et al.^133^. We assigned ancestry to the ABCD participants based on self-reported ethnic background combined with SNPweights^134^ inferences, see Zhao, et al. ^45^ for more information.

### Heritability and GWAS analysis

We downloaded UKB imputed genetics data^133^ (Data-Category 263) and locally imputed the ABCD genetic data using the Michigan Imputation Server (https://imputationserver.sph.umich.edu/) with the 1000 Genomes Phase 3 (Version 5) reference panel^45^. In both UKB and ABCD, the following quality controls were performed on imaging subjects with genetics data: 1) removing subjects with > 10% missing genotypes; 2) removing genetic variants with minor allele frequency (MAF) < 0.01; 3) removing genetic variants with missing genotype rate > 10%; 4) removing variants that failed the Hardy-Weinberg test at 1 × 10^-7^ significance level; and 5) removing genetic variants with imputation INFO score < 0.8. We used GCTA^44^ to estimate SNP-based heritability with all autosomal SNPs in the white British discovery dataset (average *n* = 32,631). The adjusted covariates include age (at imaging), age-squared, sex, age-sex interaction, age-squared-sex interaction, imaging site, the top 40 genetic PCs^133^, volumetric scaling, head motion, head motion-squared, brain position, and brain position-squared^37, 40^. For the 363 structure-level shape features, the genetic variance estimates were also extracted from the GCTA and were compared before and after additionally adjusting for the regional volumetric measurements. Specifically, for each of the 7 subcortical structures, we additionally controlled the corresponding FIRST subcortical volumes (Category 1102). For the lateral ventricles, we adjusted for the regional volumes estimated from ANTs^135^. The genome-wide association analysis was conducted with a linear mixed effect model using fastGWA^136^, adjusted for the same covariates as the GCTA. We also conducted GWAS separately in validation datasets and adjusted for only the top 10 genetic PCs rather than the top 40. In the ABCD dataset, we performed validation GWAS separately for African American, European, and Hispanic subjects, removing one subject randomly from each twin pair^45^. In all analyses, we removed values greater than five times the median absolute deviation from the median for each continuous phenotype or covariate variable.

We used FUMA^47^ (version v1.3.8) to characterize genomic loci with European LD files from the 1000 Genomes. To define the LD boundaries, FUMA used independent significant variants, which were genetic variants whose *P*-value smaller than the predefined threshold (here was 1.09 × 10^-10^, 5 × 10^-8^/457) and were independent of other significant variants (LD *r*^2^ < 0.6). FUMA then constructed LD blocks for these independent significant variants by tagging all variants in LD (*r*^2^ ≥ 0.6) with at least one independent significant variant with a MAF ≥ 0.0005. There may have been variants from the 1000 Genomes reference panel that were not included in the GWAS. Moreover, within these significant variants, we defined independent lead variants as those that were independent from each other (LD *r*^2^ < 0.1). In the case of close LD blocks (<250 kb based on the closest boundary variants of LD blocks), they were merged into one genomic locus. Independent significant variants and all the variants in LD with them (*r*^2^ ≥ 0.6) were looked up on the NHGRI-EBI GWAS catalog (version e104_2021-09-15) to search for associations (*P* < 9 × 10^-6^) reported for any traits. For selected colocalized index variants, we also performed association analysis in vertex-wise data to illustrate local association patterns. The significance threshold was set to be 0.05/number of vertices in each structure. We adjusted for the same set of covariates as in the above genome-wise analysis.

Gene-based testing was performed using UKB white British discovery GWAS summary statistics for 18,796 protein-coding genes via MAGMA^49^ (version 1.08). We used default MAGMA settings with zero window size around each gene. We also conducted functional annotation and mapping analysis in FUMA, where genetic variants were annotated with their genomic functionality and then were mapped to 35,808 candidate genes using positional, eQTL, and 3D chromatin interaction information. Brain-related tissues/cells were selected in all options and the default values were used for all other parameters in FUMA. LDSC^88^ (version 1.0.1) was used to infer genetic correlations. LD scores were computed using 1000 Genomes European data provided by LDSC. The major histocompatibility complex (MHC) region was removed from the HapMap3 variants.

### Polygenic risk scores on UKB non-imaging subjects

As a first step, we constructed a PRS based on PRS-CS^99^ for each shape feature. We input GWAS summary statistics from the UKB white British discovery dataset (average *n* = 32,631), and randomly selected 1,500 subjects from the UKB European validation dataset as validation. We used all default parameters in the PRS-CS software (https://github.com/getian107/PRScs) and generated the PRS for all non-imaging individuals in the UKB study (removing relatives of the UKB imaging individuals). The second step was to explore the associations with 276 phenotypes across various trait domains using these non-imaging UKB individuals, including 24 mental health traits (Category 100060), 5 cognitive traits (Category 100026), 12 physical activity traits (Category 100054), 6 electronic device use traits (Category 100053), 8 sun exposure traits (Category 100055), 3 sexual factor traits (Category 100056), 3 social support traits (Category 100061), 12 family history of diseases (Category 100034), 21 diet traits (Category 100052), 9 alcohol drinking traits (Category 100051), 6 smoking traits (Category 100058), 34 blood biochemistry biomarkers (Category 17518), 3 blood pressure traits (Category 100011), 3 spirometry traits (Category 100020), 32 early life factors (Categories 135, 100033, 100034, and 100072), 9 greenspace and coastal proximity (Category 151), 2 hand grip strength (Category 100019), 13 residential air pollution traits (Category 114), 5 residential noise pollution traits (Category 115), 2 body composition traits by impedance (Category 100009), 4 health and medical history traits (Category 100036), 3 female specific factors (Category 100069), 1 education trait (Category 100063), and 57 curated disease phenotypes based on Dey, et al. ^137^ (**Table S11**).

We used a discovery-validation design and repeated our analysis in two independent samples: 1) the discovery sample, which consisted of 70% randomly selected independent UKB non-imaging subjects of white British ancestry (average *n* = 202,405) and 2) the validation sample, including the left 30% independent UKB white British non-imaging subjects (average *n* = 86,736), white non-British non-imaging subjects (average *n* = 20,746), and non-white non-imaging subjects (average *n* = 21,587). The adjusted covariates included age, age-squared, sex, age-sex interaction, age-squared-sex interaction, as well as 40 genetic PCs. We reported *P* values from the two-sided t test and prioritized on the results that were 1) significant after Bonferroni correction in the discovery dataset, 2) significant at nominal significance level (0.05) in the validation dataset; and 3) the regression coefficients had matched directions in the discovery and validation datasets.

### MR analysis with clinical endpoints

We examined the genetic causal relationships between the 457 shape features and 288 clinical endpoints, where 275 of them were from FinnGen (https://www.finngen.fi/en/access_results) and 13 were from the PGC (https://pgc.unc.edu/). For FinnGen, we selected 275 clinical traits from the latest release (R7) and with more than 5,000 cases. To reduce the potential influence of sample overlap, we avoid PGC studies that have been using data solely from the UKB study. More detailed information can be found in **Table S13**.

Before running MR methods, we performed standard preprocessing steps on GWAS data. The genetic variants were first selected with significance threshold 5 × 10^-8^ in the exposure GWAS. To ensure the genetic variants used in the MR were independent, LD clumping was implemented using *r*^2^ = 0.01, window size = 10,000, and the 1000 Genomes European ancestry data being the reference panel. The harmonization procedure in the TwoSampleMR package (https://mrcieu.github.io/TwoSampleMR/) helped us infer the correct allele alignment, therefore the selected variants on the exposure and the reported effect of the same variant on the outcome corresponded to the same allele.

We tested 14 MR methods^114–117^, including the IVW, IVW multiple random effect model, IVW fixed effect model, MR-Egger, Simple Median, Weighted Median, Penalized Weighted Median, Simple Mode, Simple Mode (NOME), Weighed Mode, Weighted Mode (NOME), DIVW, GRAPPLE, and MR-RAPS. To ensure the reliability of our results, we compared estimates from different methods. After running MR analysis on all pairs between brain shape features and clinical endpoints, we used two steps to select the significant causal results. We first removed all the estimated causal associations with less than 6 variants used in the MR analysis. Then for the remaining estimates, we performed Bonferroni adjustments for multiple testing. Besides comparing estimates across different MR methods, we also tested potential violations in MR analysis to make sure our results were reliable. For example, a significant intercept of MR Egger regression indicated the presence of horizontal pleiotropy. All reported results have passed these tests.

### Code availability

We made use of publicly available software and tools. The pipelines used in shape feature extractions can be found at https://www.nitrc.org/frs/?group_id=1461. The codes used in other parts of the paper are available upon reasonable request.

### Data availability

The individual-level data used in the present study can be applied from the UKB (https://www.ukbiobank.ac.uk/) and ABCD (https://abcdstudy.org/) studies. Our GWAS summary statistics will be shared on Zenodo and at the BIG-KP https://bigkp.org/. Our GWAS results will also be available via the interactive web browser at http://165.227.78.169:443/.

## Notes

### Competing Interest Statement

The authors have declared no competing interest.

### Author Declarations

The individual-level data used in the present study have been openly available from the UK Biobank (https://www.ukbiobank.ac.uk/) and ABCD (https://abcdstudy.org/) studies.

