## Supplementary material for "Genetic influences on the shape of brain ventricular and subcortical structures": supp_figures

**This PDF file includes:**

Supplementary Figures S1 to S42

**Other Supplementary Materials for this manuscript include the following:**

Supplementary Note (available in a PDF file)

Supplementary Tables S1 to S14 (.xlsx) (available in a zip file)

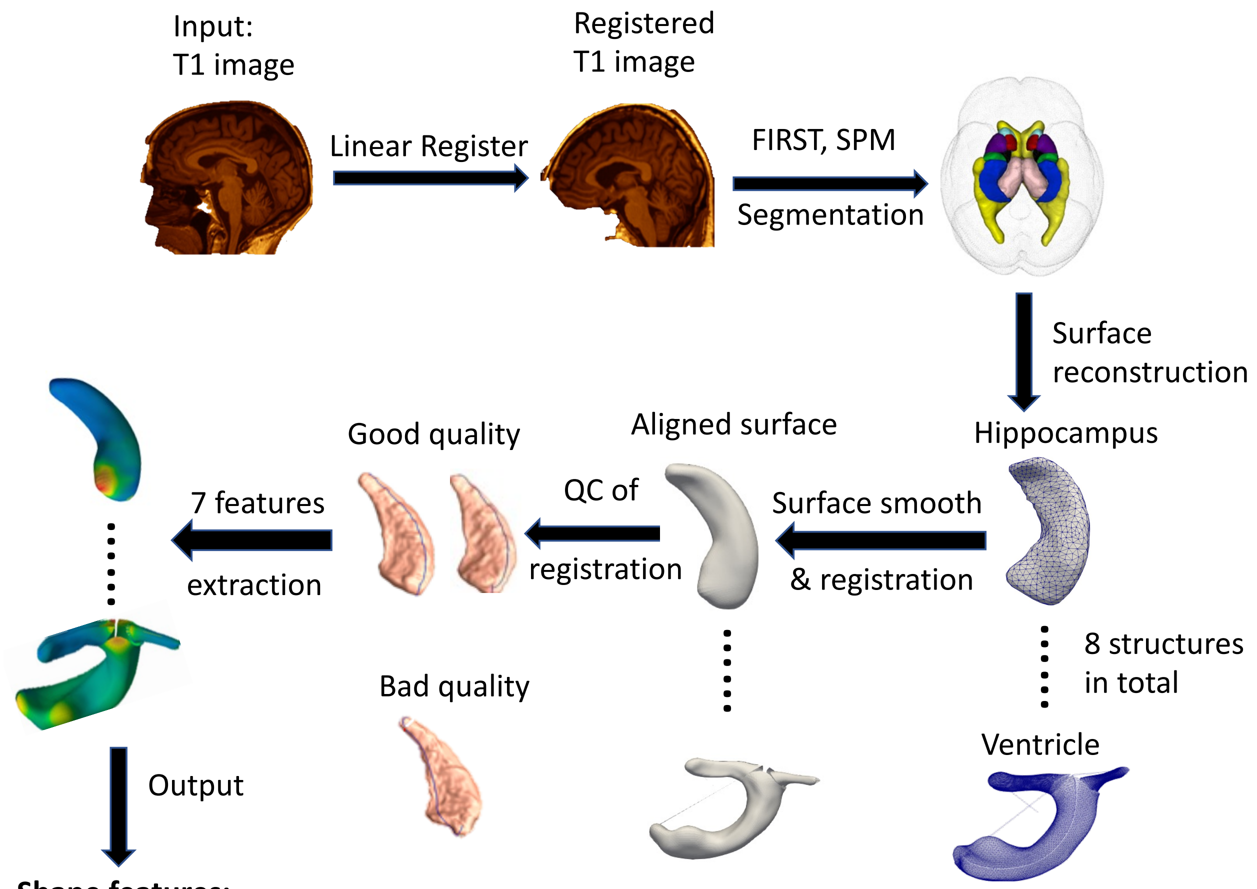


**Shape features:**

1) Top 10 principal components and regional average across each of the 8 subcortical structure
2) Subfield average according to divisions of 7 hippocampal subfields

**Fig. S1 Overview of the workflow to extract shape features.**

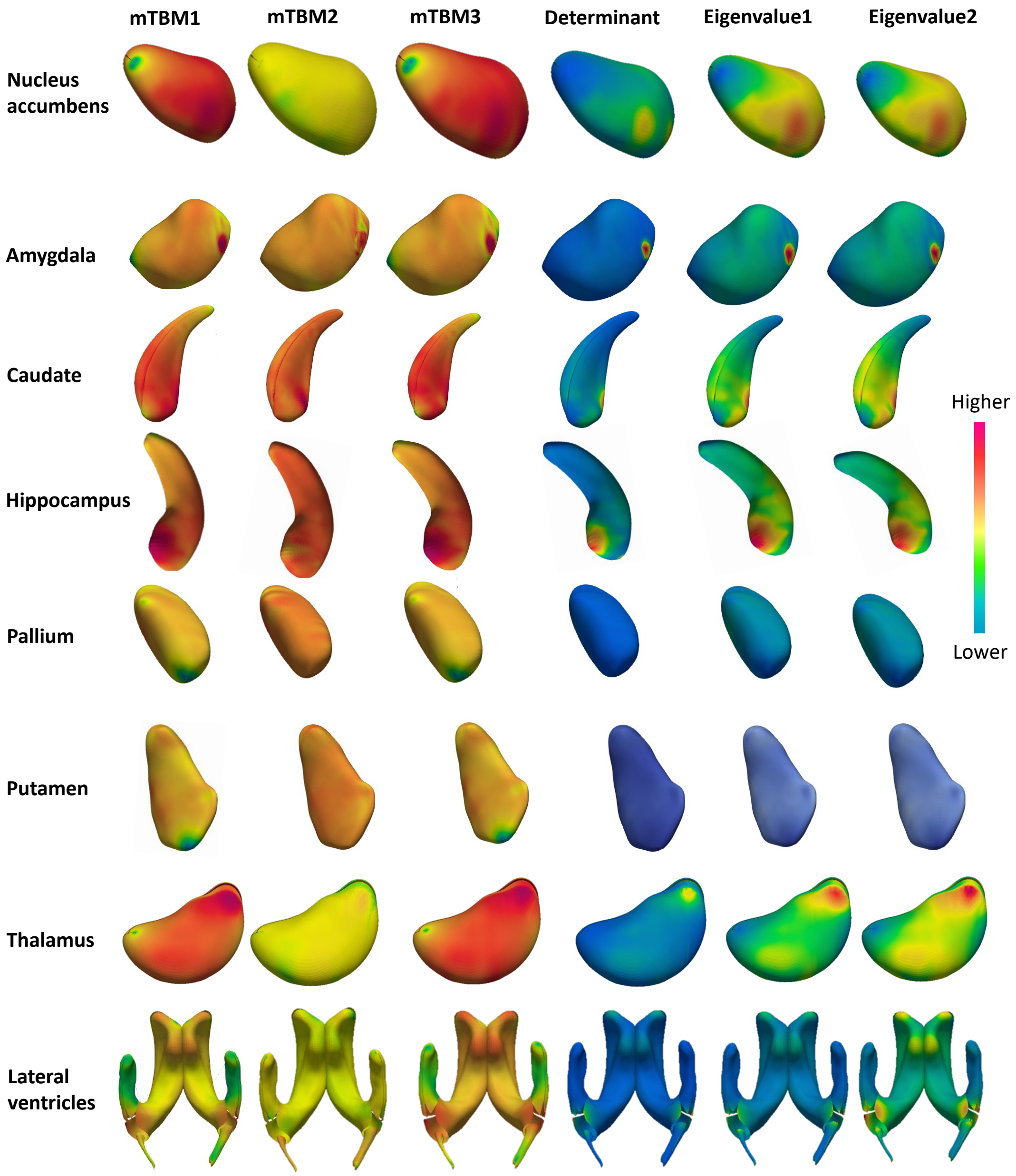


**Fig. S2 Illustration of the structures and shape features.** We illustrate the spatial pattern of 6 shape statistics (mTBM1, mTBM2, mTBM3, determinant, eigenvalue1, and eigenvalue2) in the vertex-wise maps of 8 ventricular and subcortical structures. These maps were generated by averaging the data from 500 randomly selected UKB subjects.

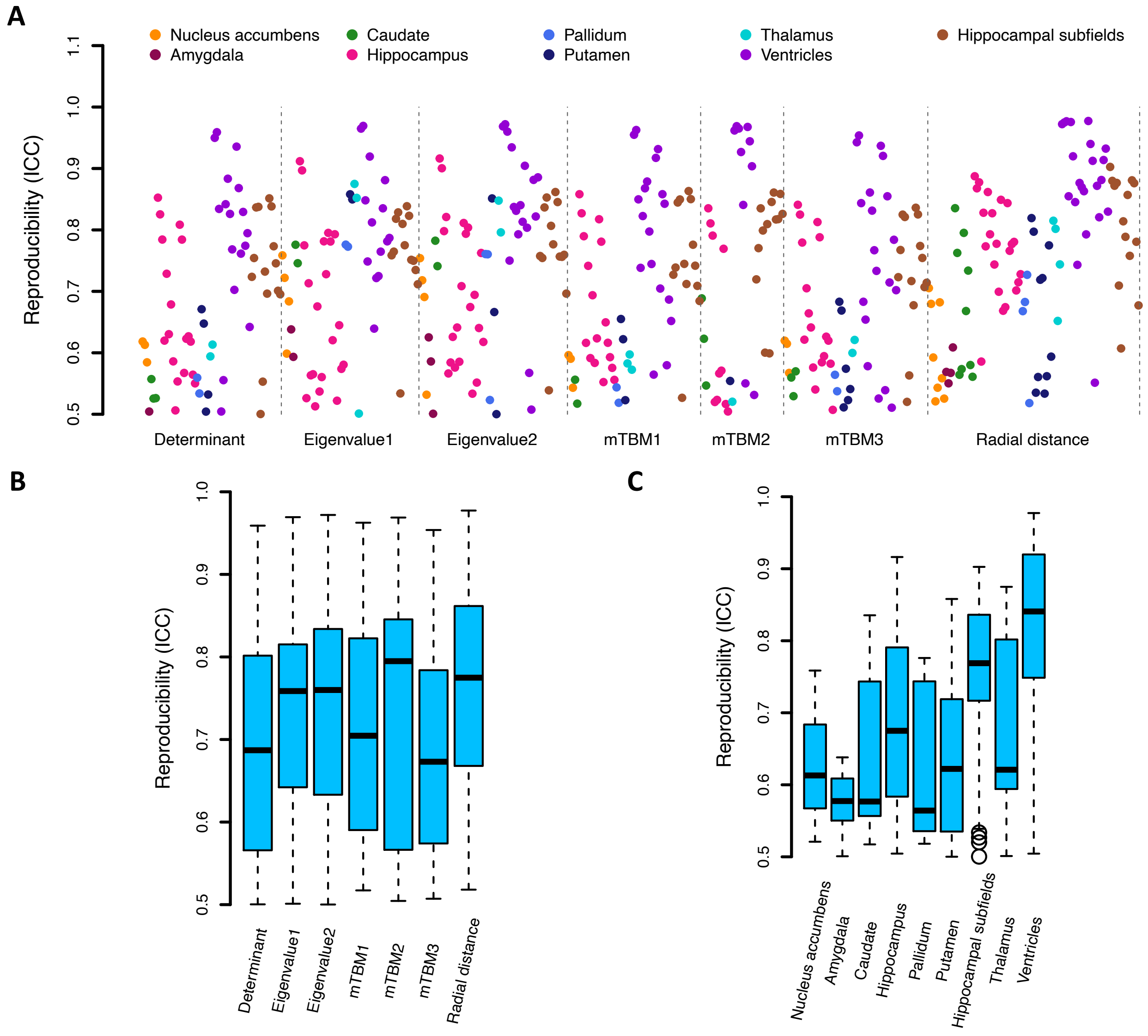


**Fig. S3 Intra-subject reproducibility of shape features.** We calculated the intraclass correlation coefficient (ICC) using subjects with repeated visits (average *n* = 2,788).

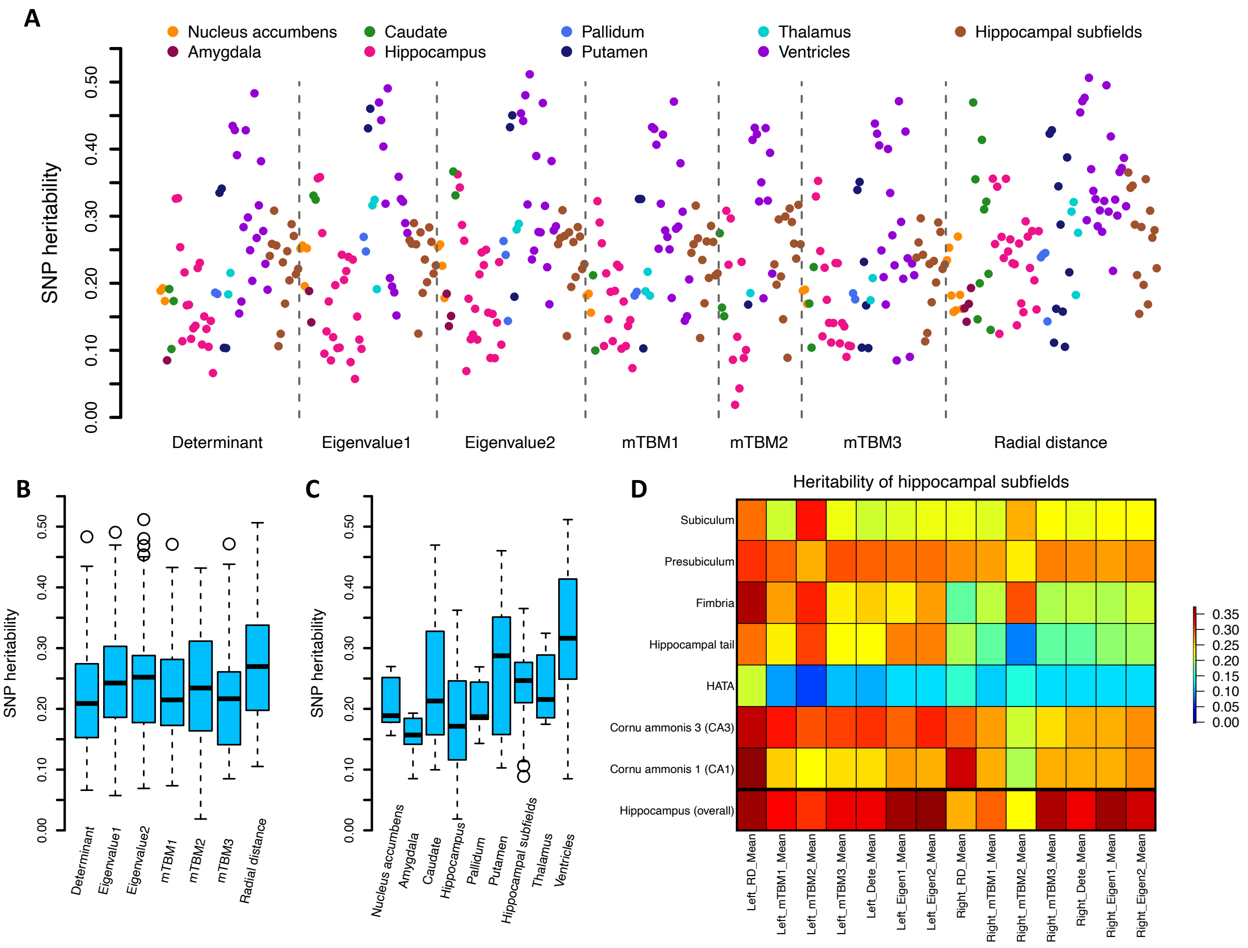


**Fig. S4 Summary of SNP heritability estimates.**

Heritability was estimated using UKB individuals of British ancestry UKB individuals of British ancestry (average sample size $n$ = 32,631). **A:** Heritability estimates of different structures and shape statistics. **B:** Comparison of the heritability estimates among different shape statistics. **C:** Comparison of the heritability estimates among different structures. **D:** Heritability estimates for shape features of subfields of the hippocampus.

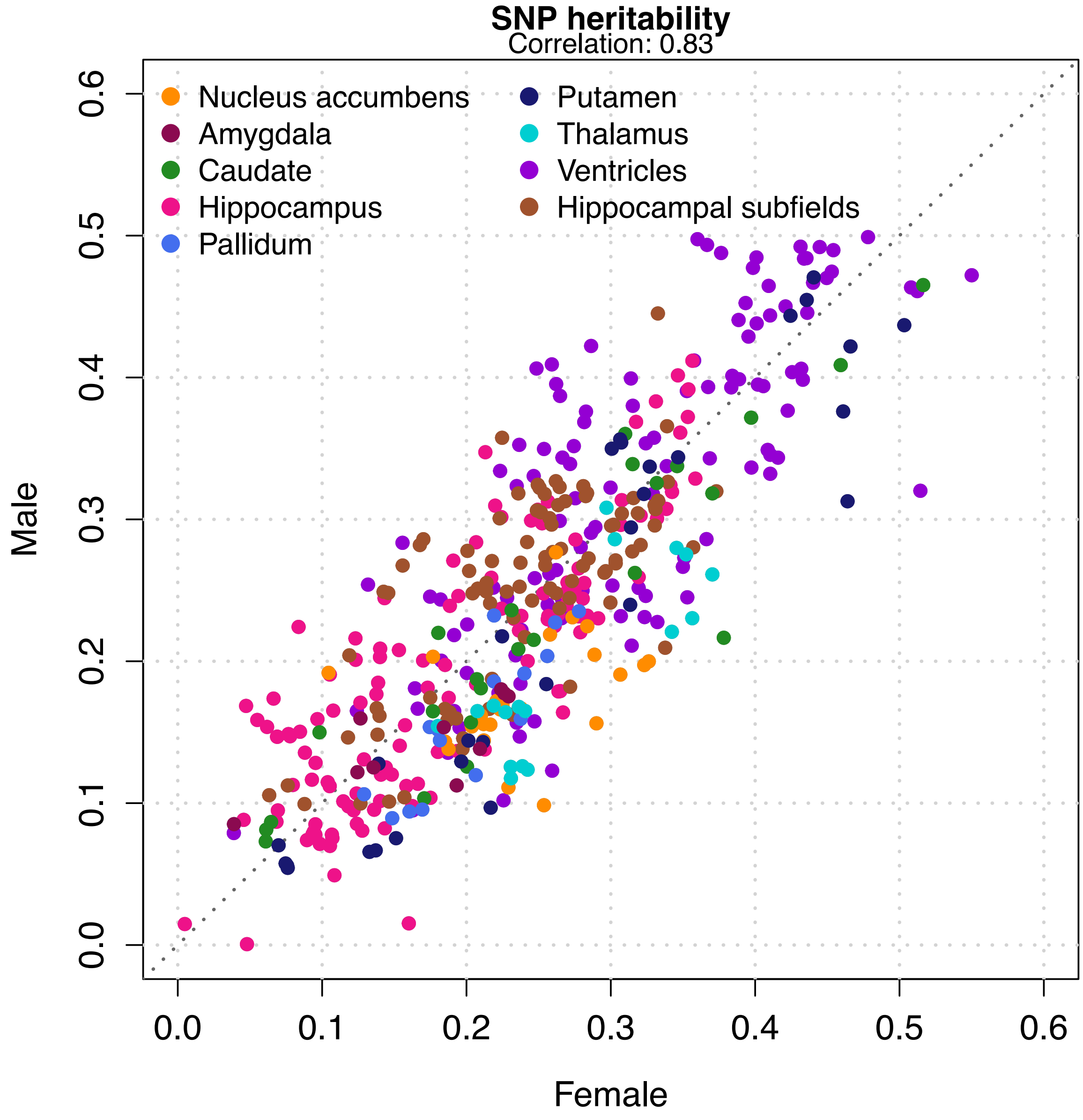


**Fig. S5 Heritability estimates in the sex-specific analysis.**

Heritability was separately estimated in male and female subjects.

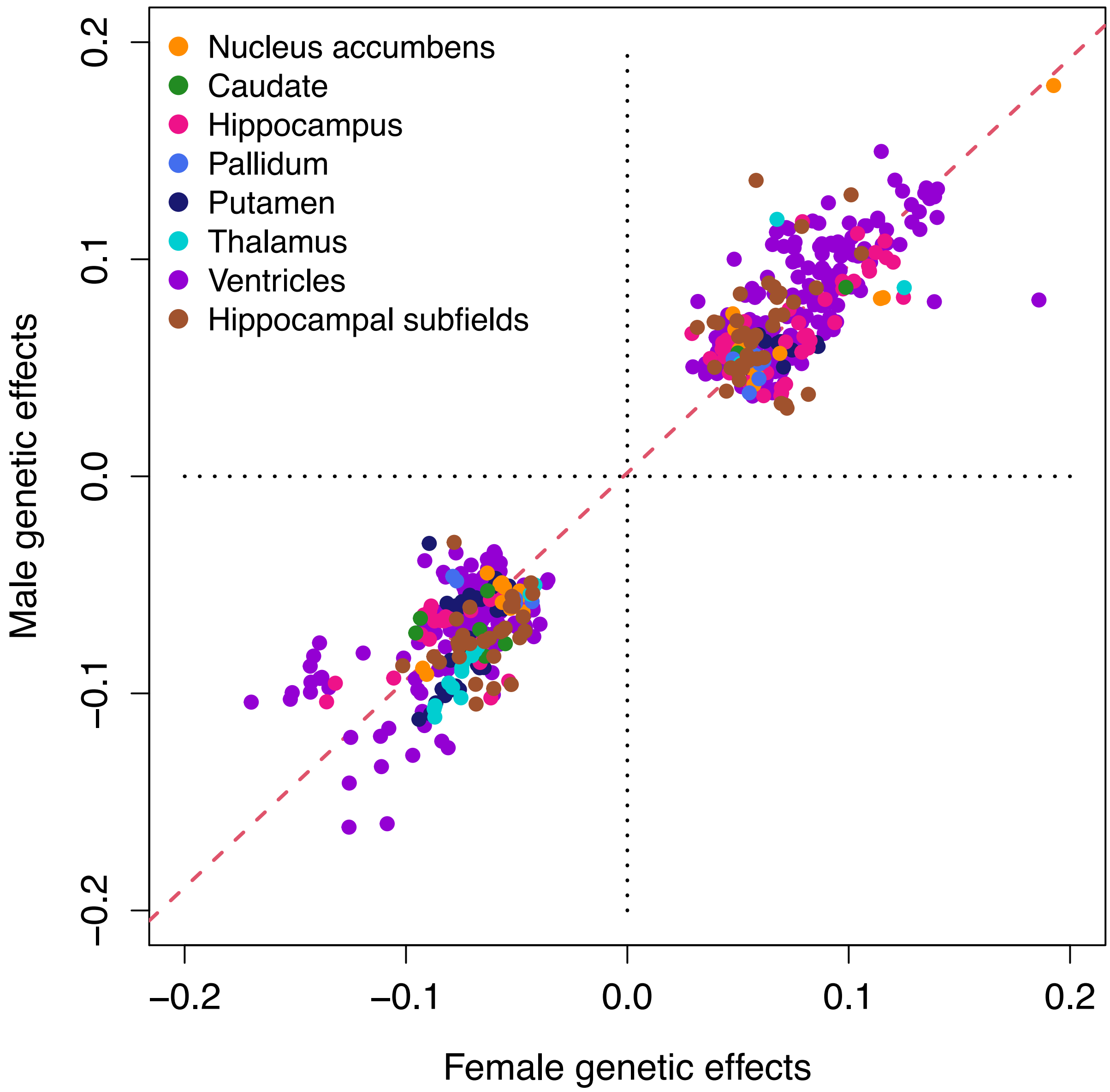


**Fig. S6 Genetic effects in the sex-specific GWAS.**

GWAS was separately conducted for male and female subjects.

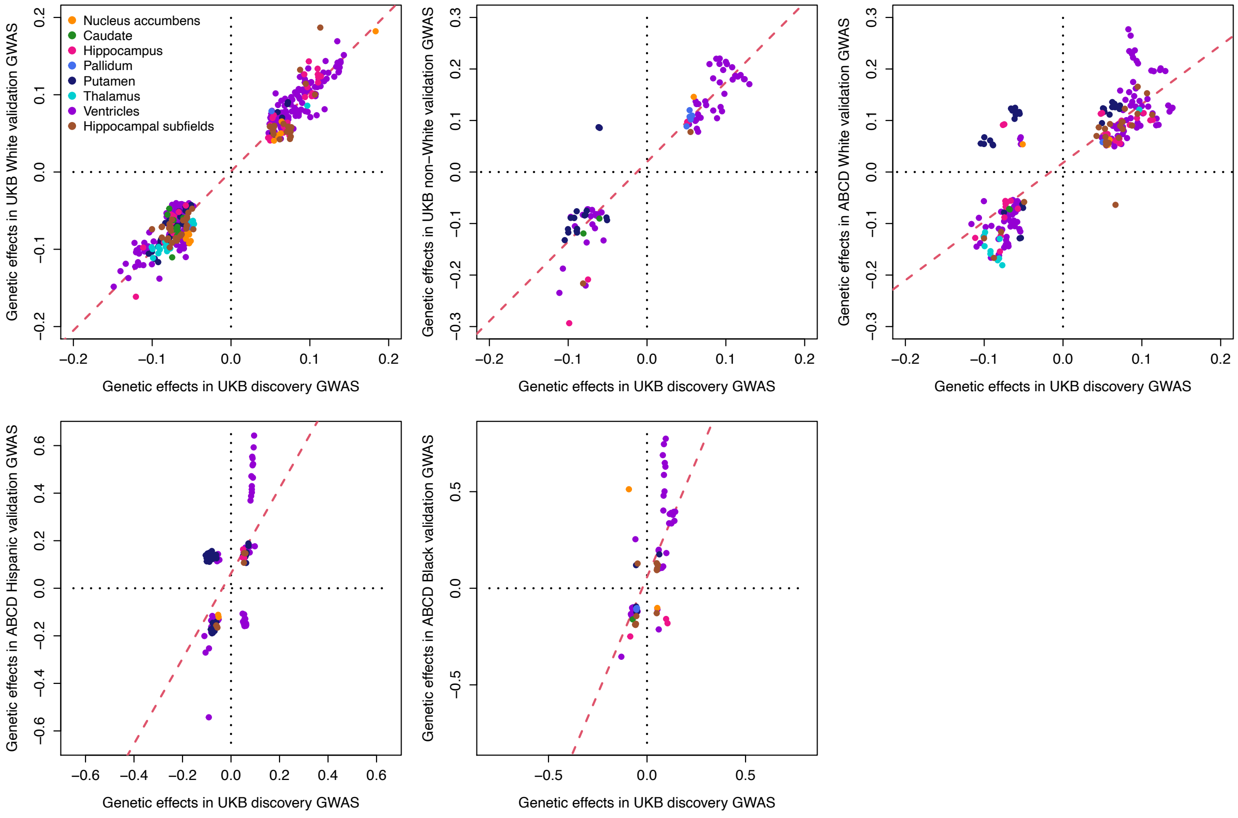


**Fig. S7 Genetic effects in discovery and validation GWAS.**

We show the genetic effects (the beta estimates) of associations that passed the nominal significance level (0.05) in 5 different validation GWAS, including the UKB white validation GWAS, UKB non-white validation GWAS, ABCD white validation GWAS, ABCD Hispanic validation GWAS, and ABCD black validation GWAS.

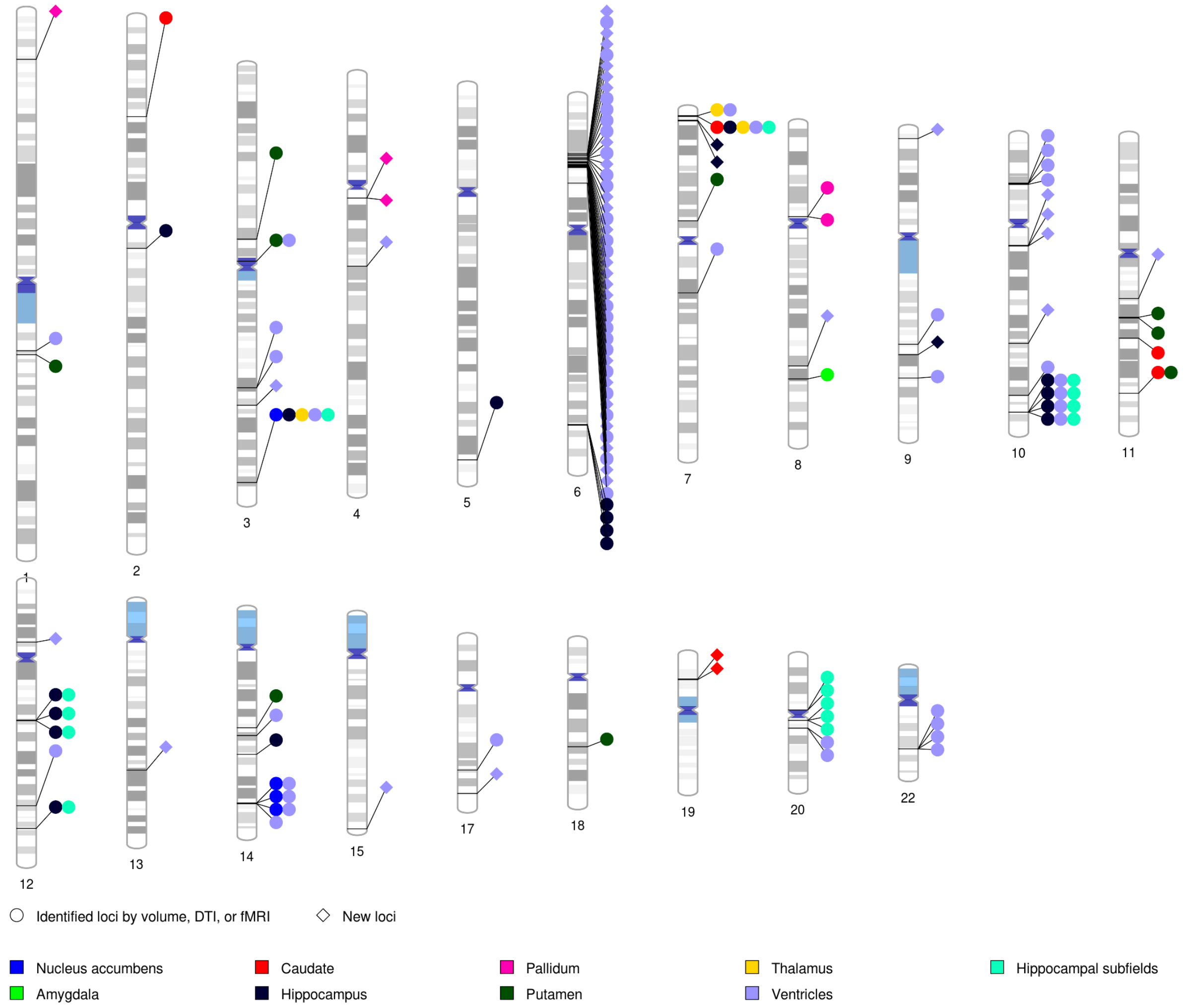


**Fig. S8 MAGMA-significant genes associated with shape features.**

There were 127 MAGMA-significant genes associated with shape features from ventricular and subcortical structures. Each signal dot indicates that at least one of the shape traits of this structure was associated with this gene. Newly identified genes and genes have been previously identified by other imaging modalities (regional brain volumes, DTI parameters, and fMRI traits) are presented by different shapes.

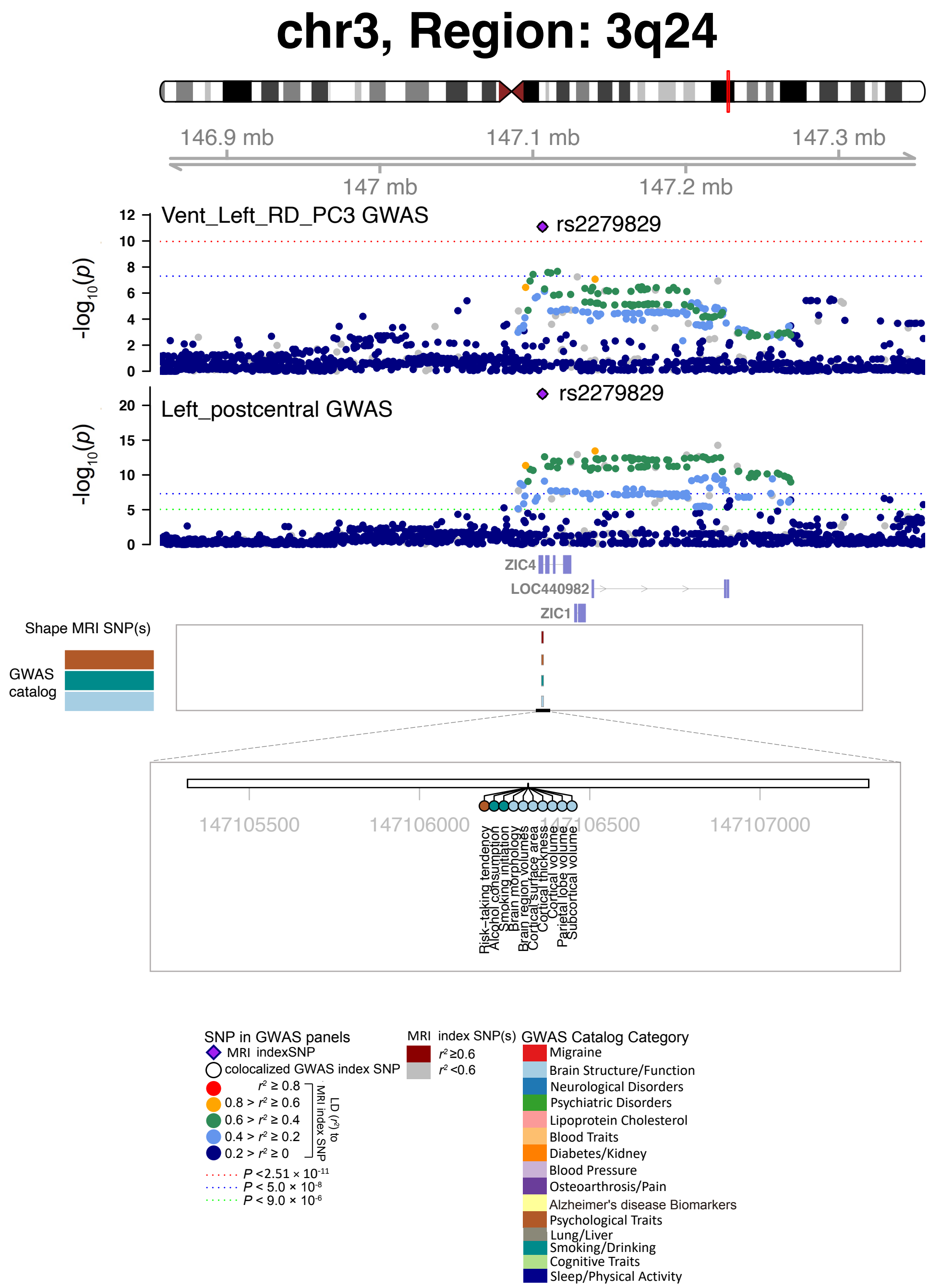


**Fig. S9 Selected genetic locus that were associated with both shape features and other complex traits.**

In 3q24, we observed shared genetic influences (LD $r^2 \geq 0.6$) between shape features (e.g., Vent_Left_RD_PC3, index variant rs2279829) and regional brain volumes (e.g., Left_postcentral, index variant rs2279829). Vent_Left_RD_PC3, third PC of the radial distance in left lateral ventricle; Left_postcentral, regional brain volume in the left postcentral region.

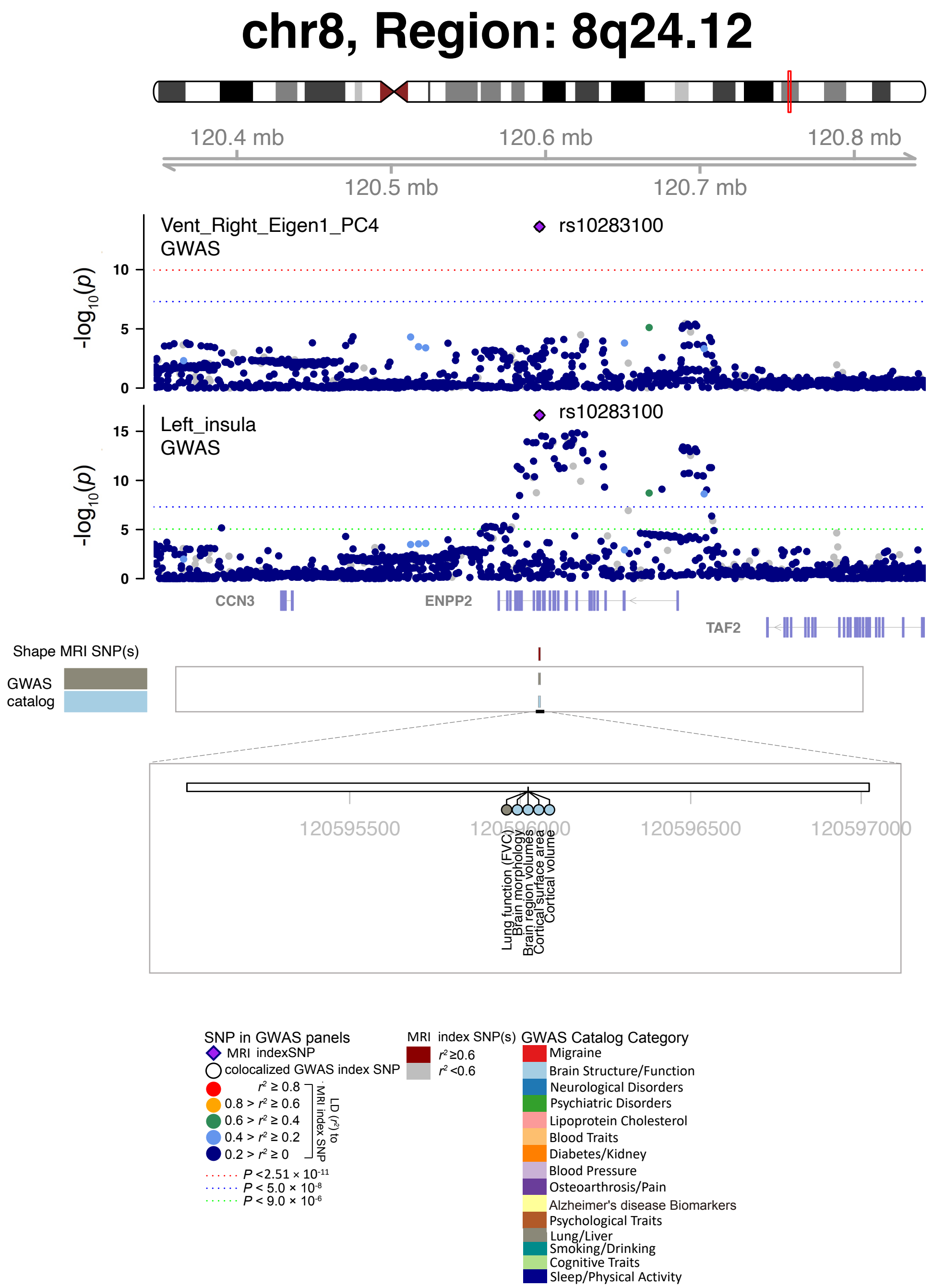


**Fig. S10 Selected genetic locus that were associated with both shape features and other complex traits.**

In 8q24.12, we observed shared genetic influences (LD $r^2 \geq 0.6$) between shape features (e.g., Vent_Right_Eigen1_PC4, index variant rs10283100) and regional brain volumes (e.g., Left_insula, index variant rs10283100). Vent_Right_Eigen1_PC4, the fourth PC of the largest eigenvalue in the right ventricles; Left_insula, regional brain volume in the left insula region.

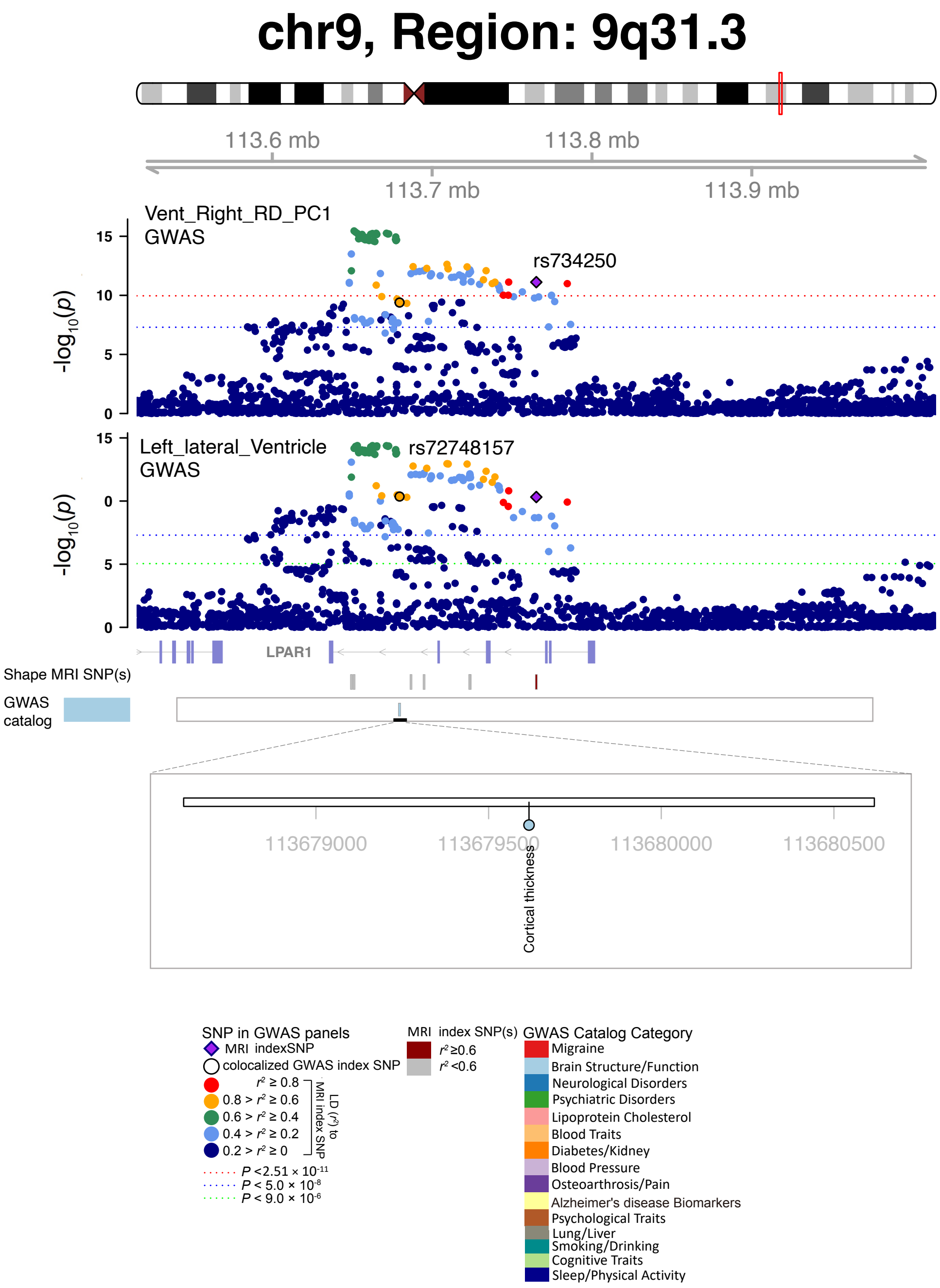


**Fig. S11 Selected genetic locus that were associated with both shape features and other complex traits.**

In 9q.31.3, we observed shared genetic influences (LD $r^2 \geq 0.6$) between shape features (e.g., Vent_Right_RD_PC1, index variant rs734250) and regional brain volumes (e.g., Left_lateral_Ventricle, index variant rs72748157). Vent_Right_RD_PC1, the first PC of radial distance in the right ventricles; Left_lateral_Ventricle, regional brain volume in the left lateral ventricle region.

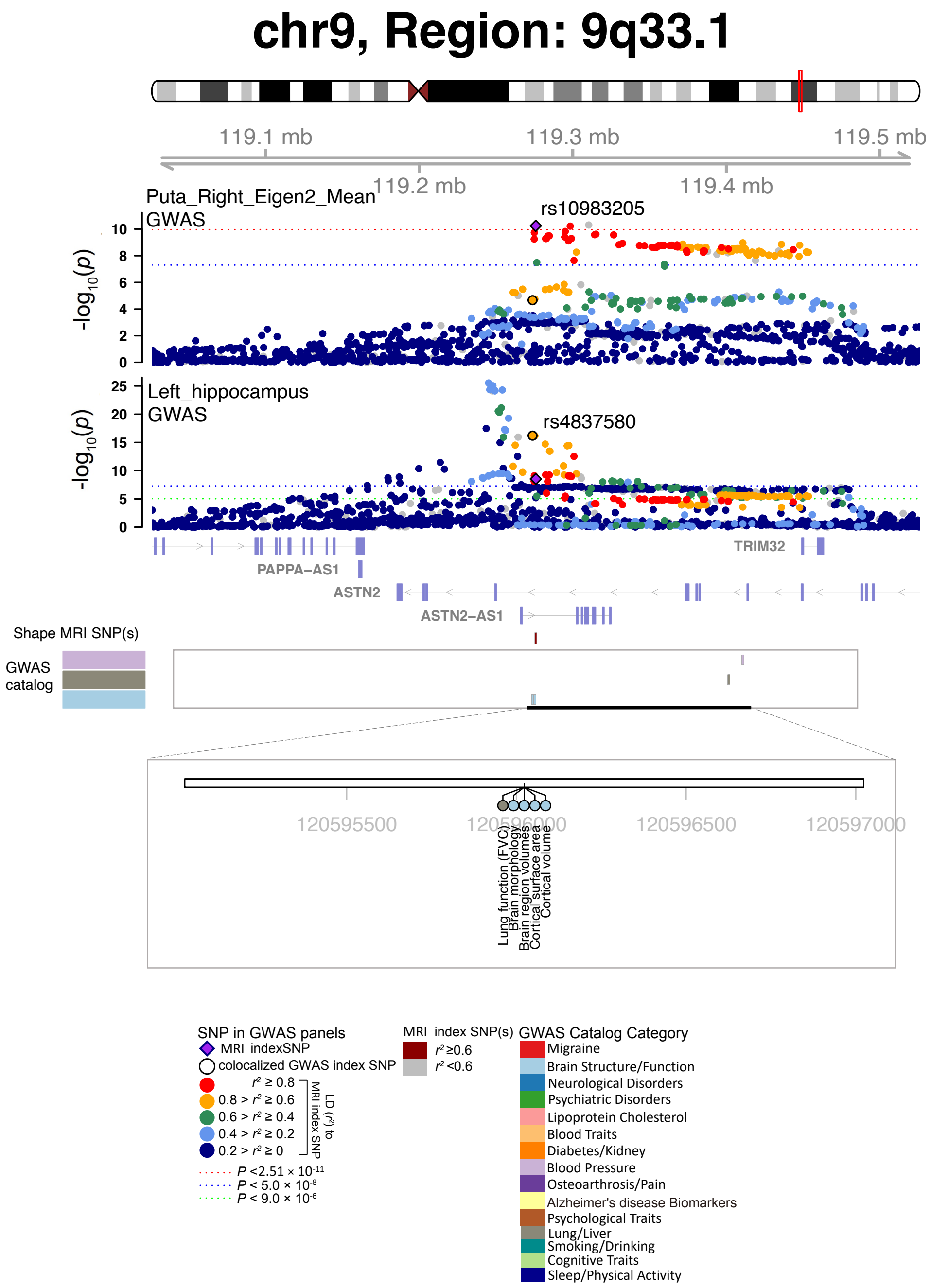


**Fig. S12 Selected genetic locus that were associated with both shape features and other complex traits.**

In 9q.33.1, we observed shared genetic influences (LD $r^2 \geq 0.6$) between shape features (e.g., Puta_Right_Eigen2_Mean, index variant rs10983205) and regional brain volumes (e.g., Left_hippocampus, index variant rs4827580). Puta_Right_Eigen2_Mean, mean of the second largest eigenvalue of the right putamen; Left_hippocampus, regional brain volume in the left hippocampus region.

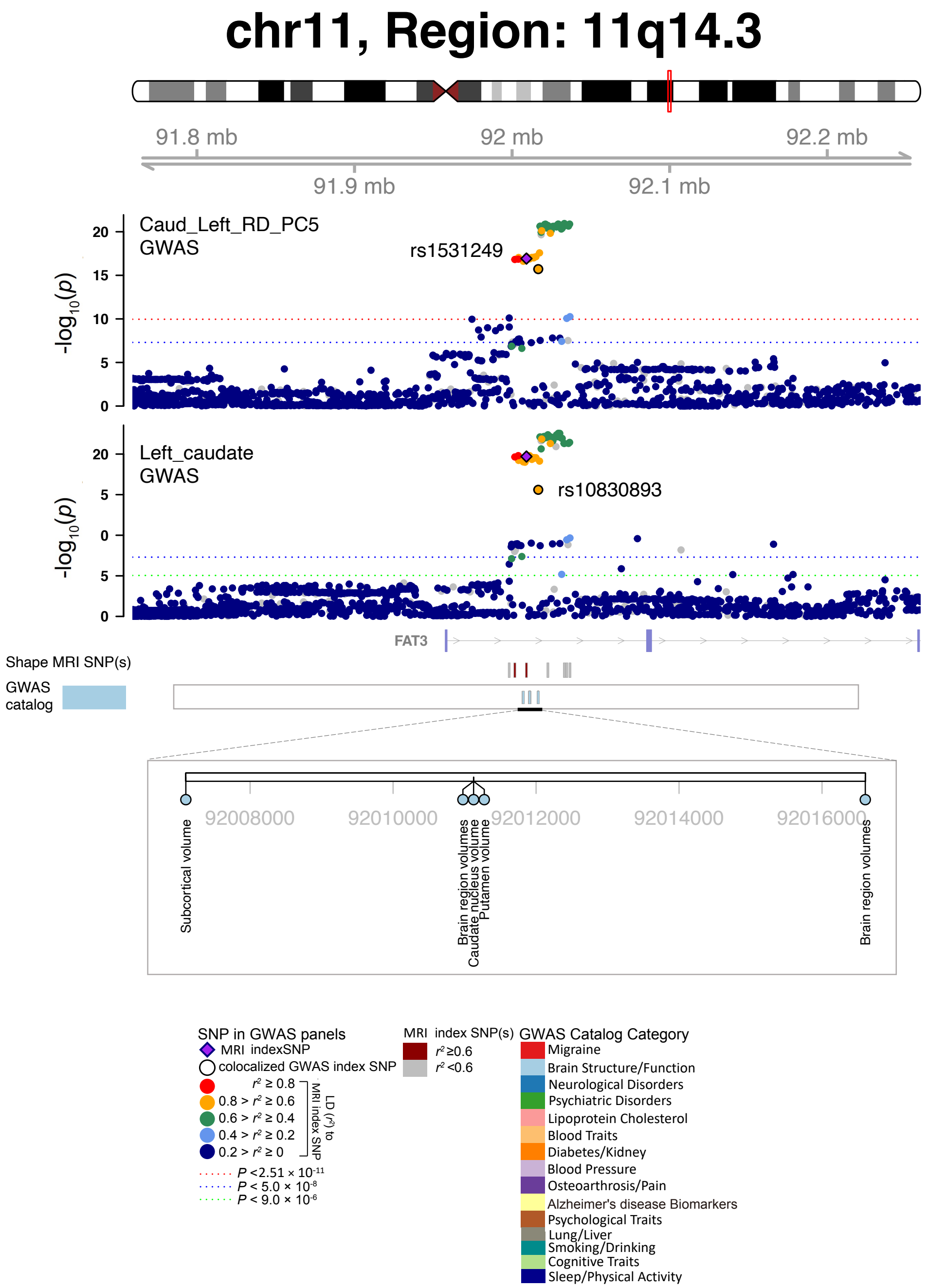


**Fig. S13 Selected genetic locus that were associated with both shape features and other complex traits.**

In 11q.14.3, we observed shared genetic influences (LD $r^2 \geq 0.6$) between shape features (e.g., Caud_Left_RD_PC5, index variant rs1531249) and regional brain volumes (e.g., Left_caudate, index variant rs10830893). Caud_Left_RD_PC5, the fifth PC of radial distance in the left caudate; Left_caudate, regional brain volume in the left caudate region.

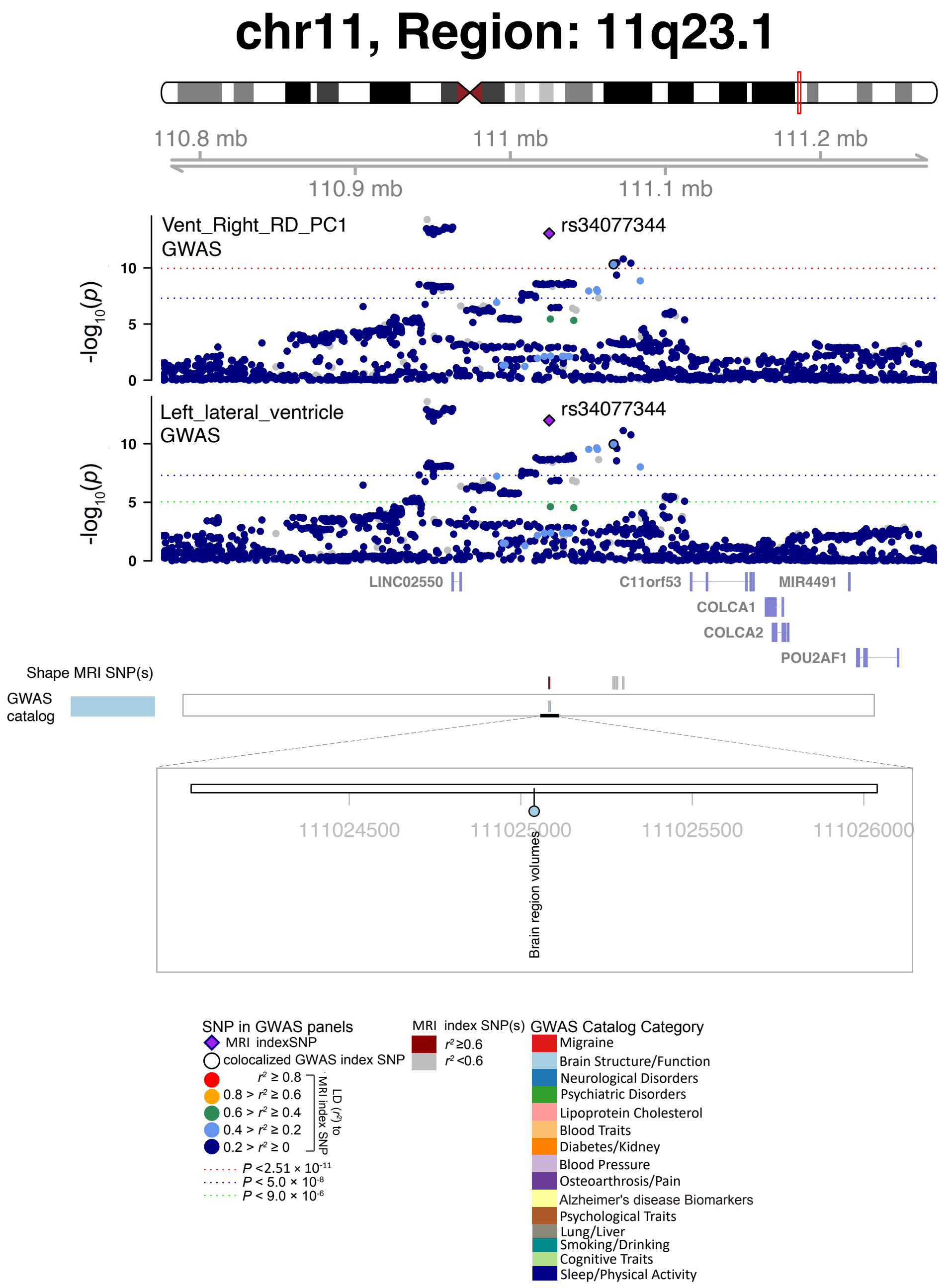


**Fig. S14 Selected genetic locus that were associated with both shape features and other complex traits.**

In 11q.23.1, we observed shared genetic influences (LD $r^2 \geq 0.6$) between shape features (e.g., Vent_Right_RD_PC1, index variant rs34077344) and regional brain volumes (e.g., Left_lateral_ventricle, index variant rs34077344). Vent_Right_RD_PC1, the first PC of radial distance in the right ventricles; Left_lateral_ventricle, regional brain volume in the left lateral ventricle region.

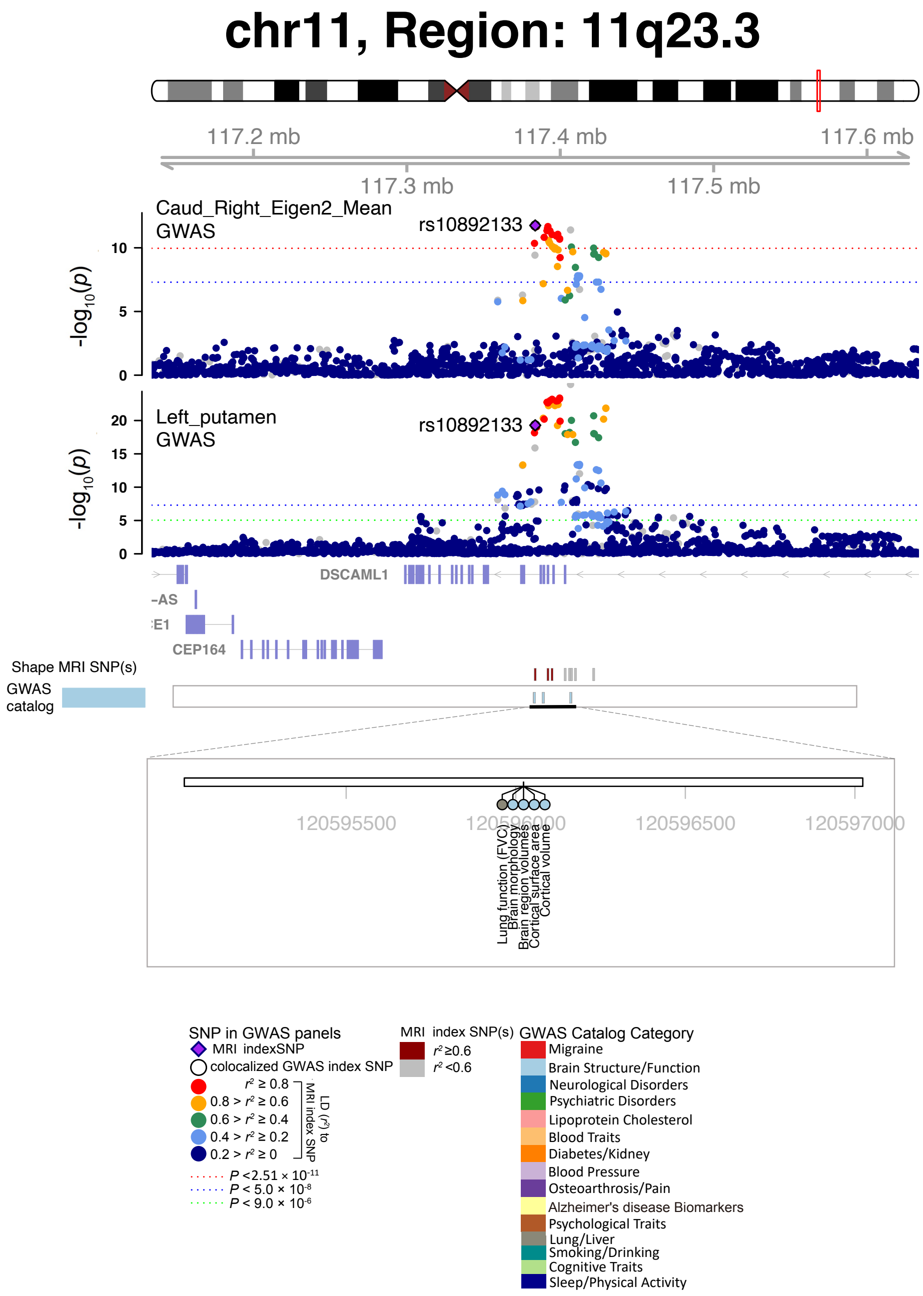


**Fig. S15 Selected genetic locus that were associated with both shape features and other complex traits.**

In 11q.23.3, we observed shared genetic influences (LD $r^2 \geq 0.6$) between shape features (e.g., Caud_Right_Eigen2_Mean, index variant rs10892133) and regional brain volumes (e.g., Left_putamen, index variant rs10892133). Caud_Right_Eigen2_Mean, mean of the second largest eigenvalue in the right caudate; Left_putamen, regional brain volume in the left putamen region.

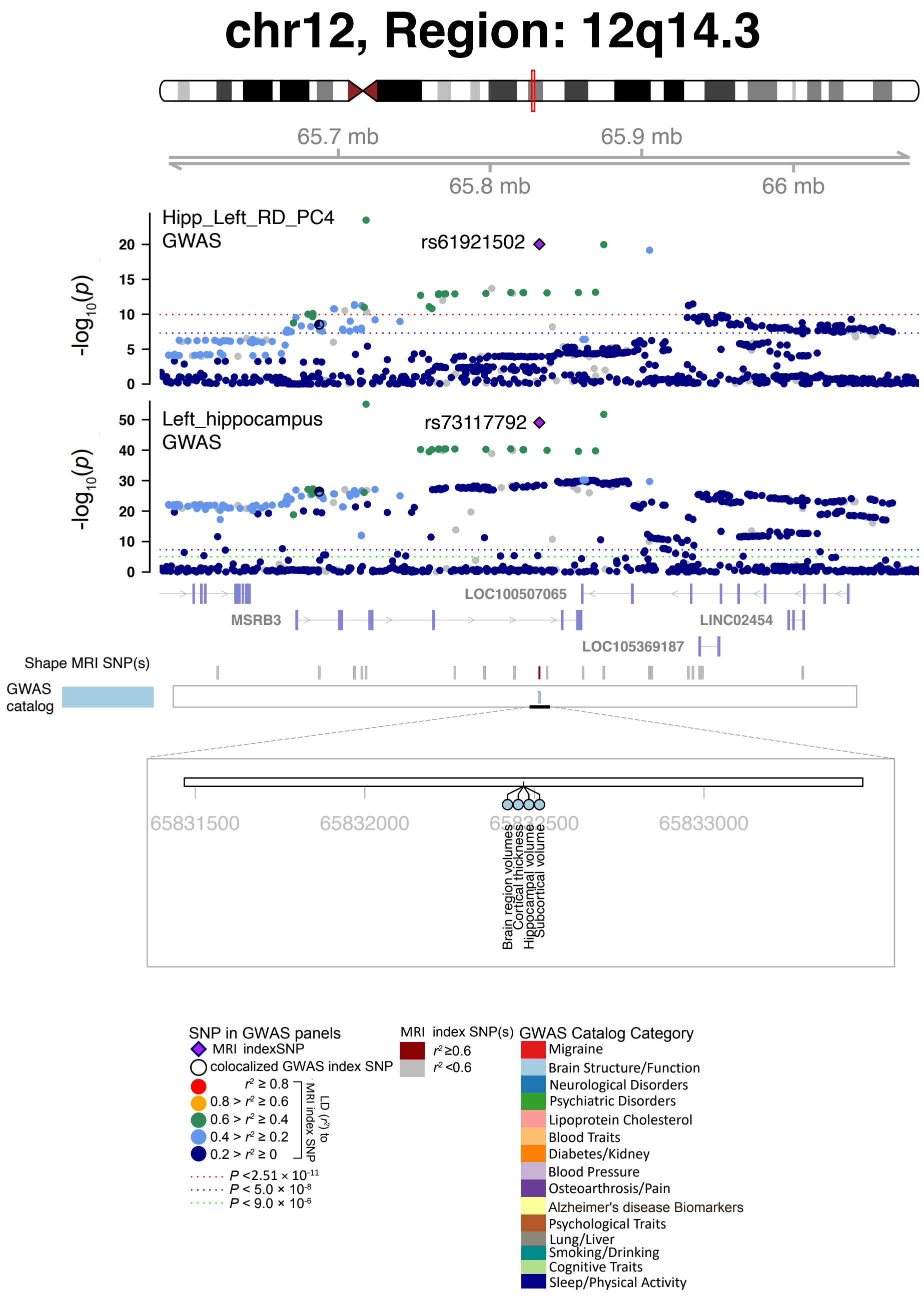


**Fig. S16 Selected genetic locus that were associated with both shape features and other complex traits.**

In 12q.14.3, we observed shared genetic influences (LD $r^2 \geq 0.6$) between shape features (e.g., Hipp_Left_RD_PC4, index variant rs61921502) and regional brain volumes (e.g., Left_hippocampus, index variant rs73117792). Hipp_Left_RD_PC4, the fourth PC of radial distance in the left hippocampus; Left_hippocampus, regional brain volume in the left hippocampus region.

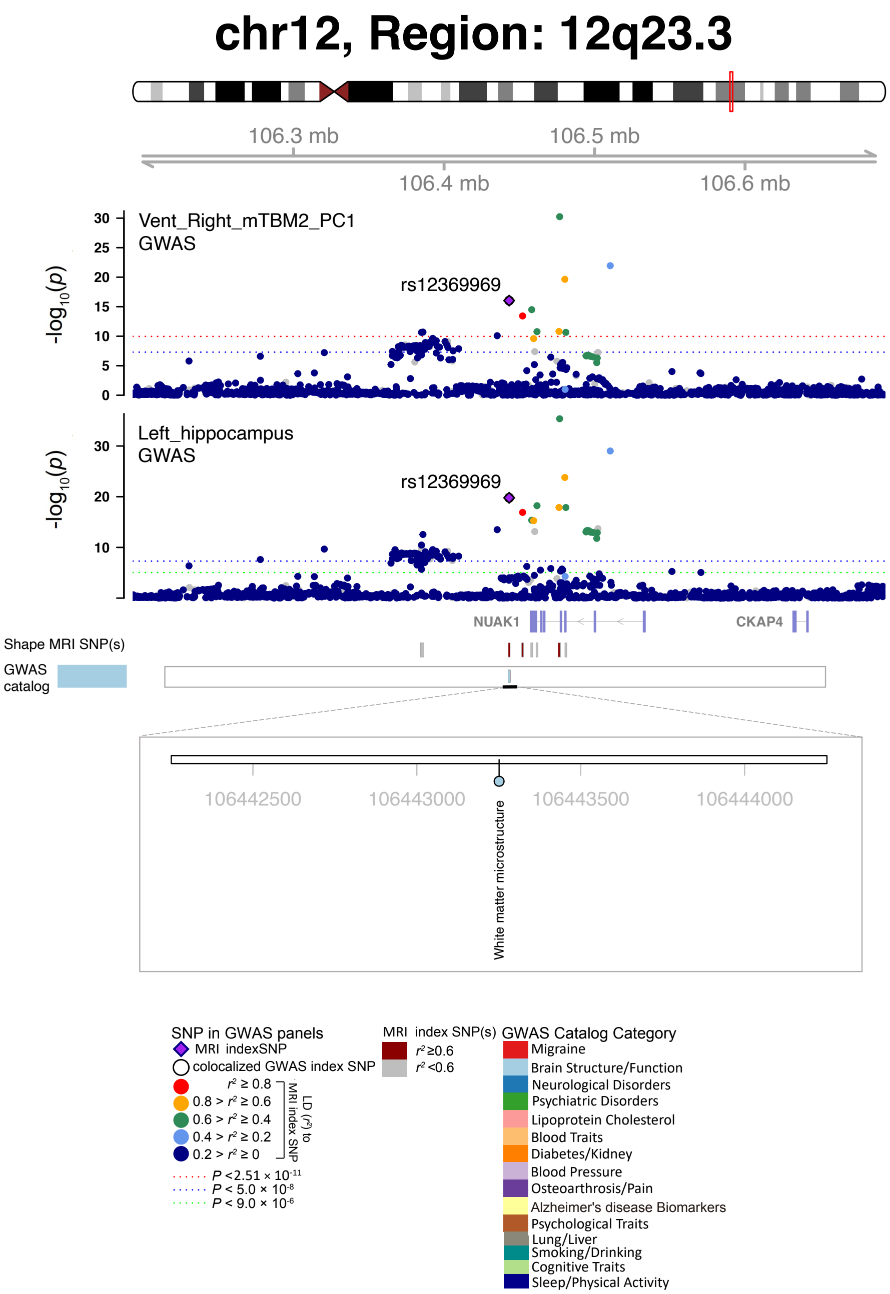


**Fig. S17 Selected genetic locus that were associated with both shape features and other complex traits.**

In 12q.23.3, we observed shared genetic influences (LD $r^2 \geq 0.6$) between shape features (e.g., Vent_Right_mTBM2_PC1, index variant rs12369969) and regional brain volumes (e.g., Left_hippocampus, index variant rs12369969). Vent_Right_mTBM2_PC1, the first PC of mTBM2 in the right ventricles; Left_hippocampus, regional brain volume in the left hippocampus region.

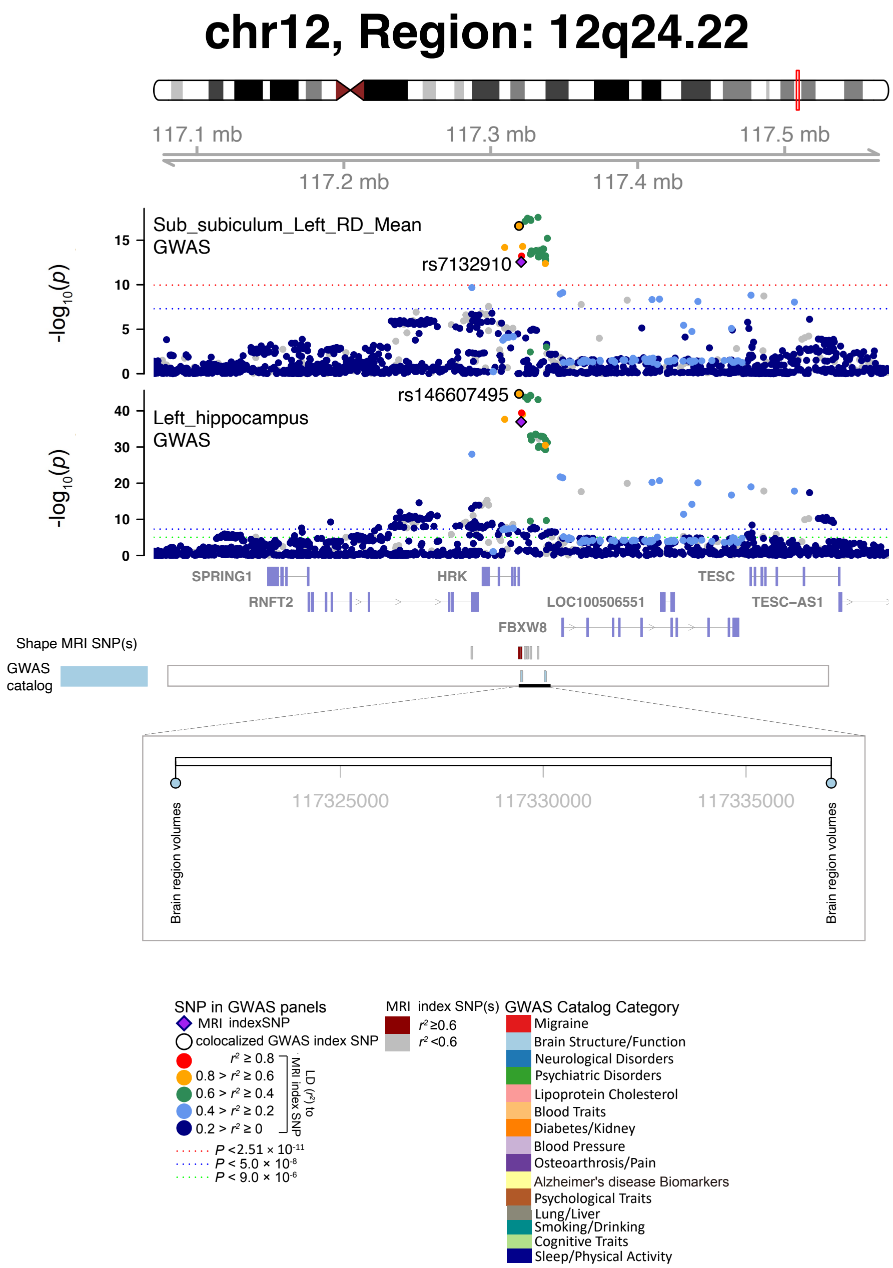


**Fig. S18 Selected genetic locus that were associated with both shape features and other complex traits.**

In 12q.24.22, we observed shared genetic influences (LD $r^2 \geq 0.6$) between shape features (e.g., Sub_subiculum_Left_RD_Mean, index variant rs7132910) and regional brain volumes (e.g., Left_hippocampus, index variant rs146607495). Sub_subiculum_Left_RD_Mean, mean of radial distance in the left hippocampal subfield subiculum; Left_hippocampus, regional brain volume in the left hippocampus region.

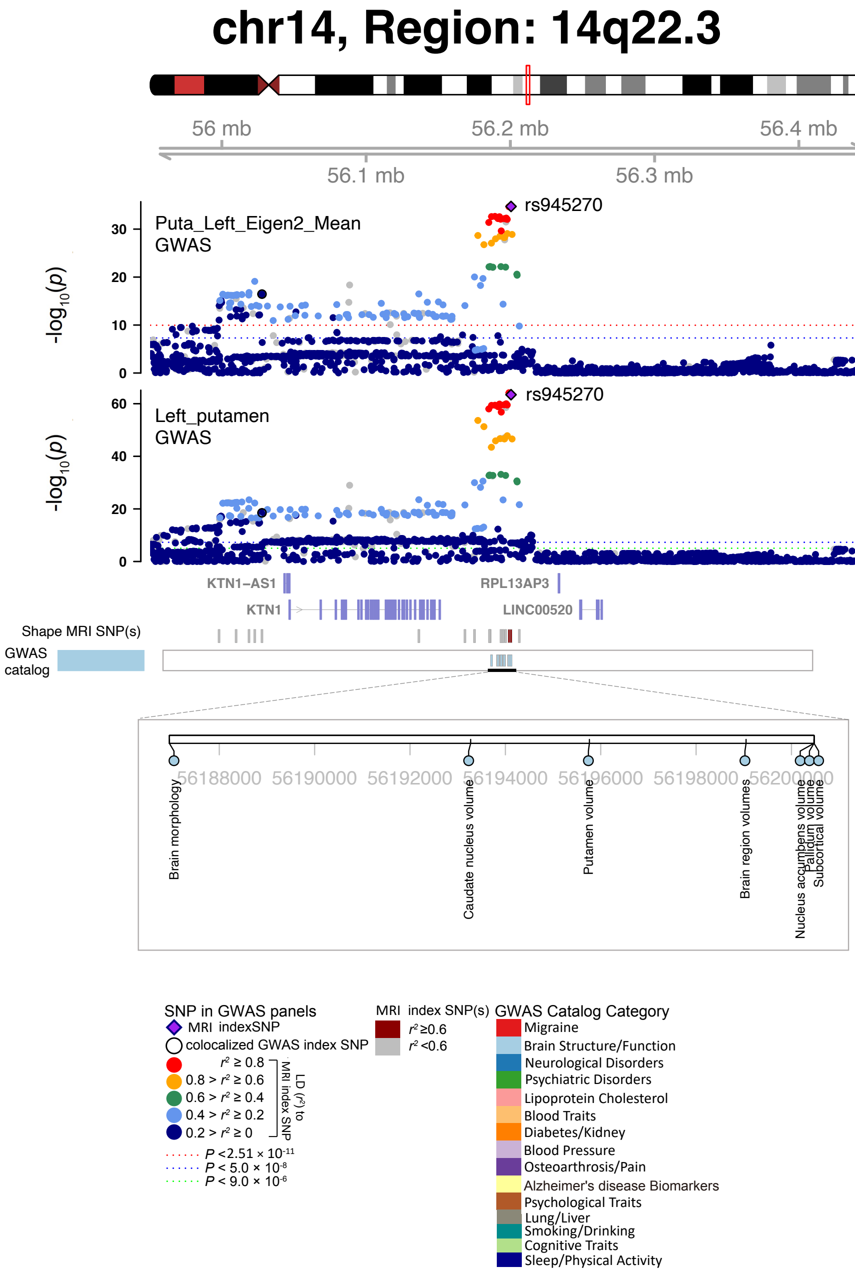


**Fig. S19 Selected genetic locus that were associated with both shape features and other complex traits.**

In 14q.22.3, we observed shared genetic influences (LD $r^2 \geq 0.6$) between shape features (e.g., Puta_Left_Eigen2_Mean, index variant rs945270) and regional brain volumes (e.g., Left_putamen, index variant rs945270). Puta_Left_Eigen2_Mean, mean of the second largest eigenvalue of the left putamen; Left_putamen, regional brain volume in the left putamen region.

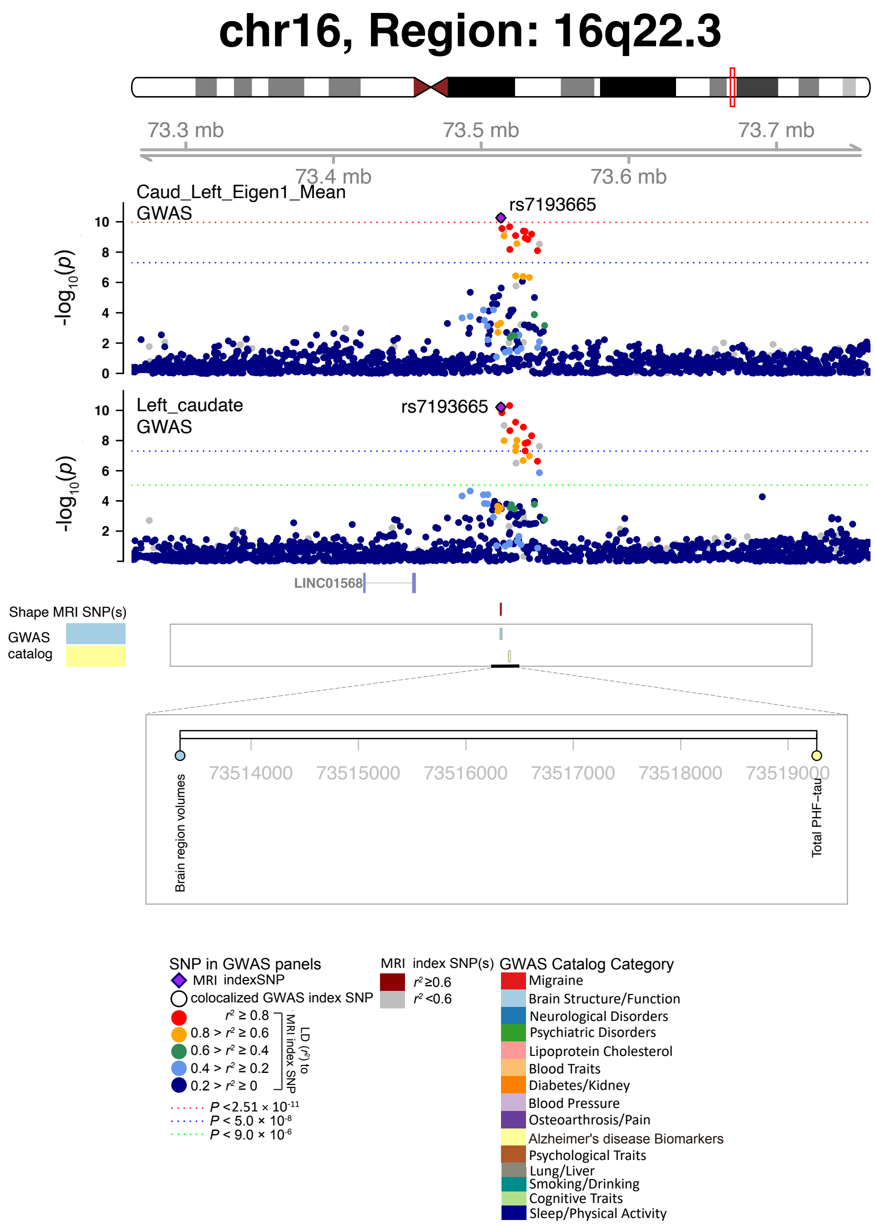


**Fig. S20 Selected genetic locus that were associated with both shape features and other complex traits.**

In 16q.22.3, we observed shared genetic influences (LD $r^2 \geq 0.6$) between shape features (e.g., Caud_Left_Eigen1_Mean, index variant rs7193665) and regional brain volumes (e.g., Left_caudate, index variant rs7193665). Caud_Left_Eigen1_Mean, mean of the largest eigenvalue in the left caudate; Left_caudate, regional brain volume in the left caudate region.

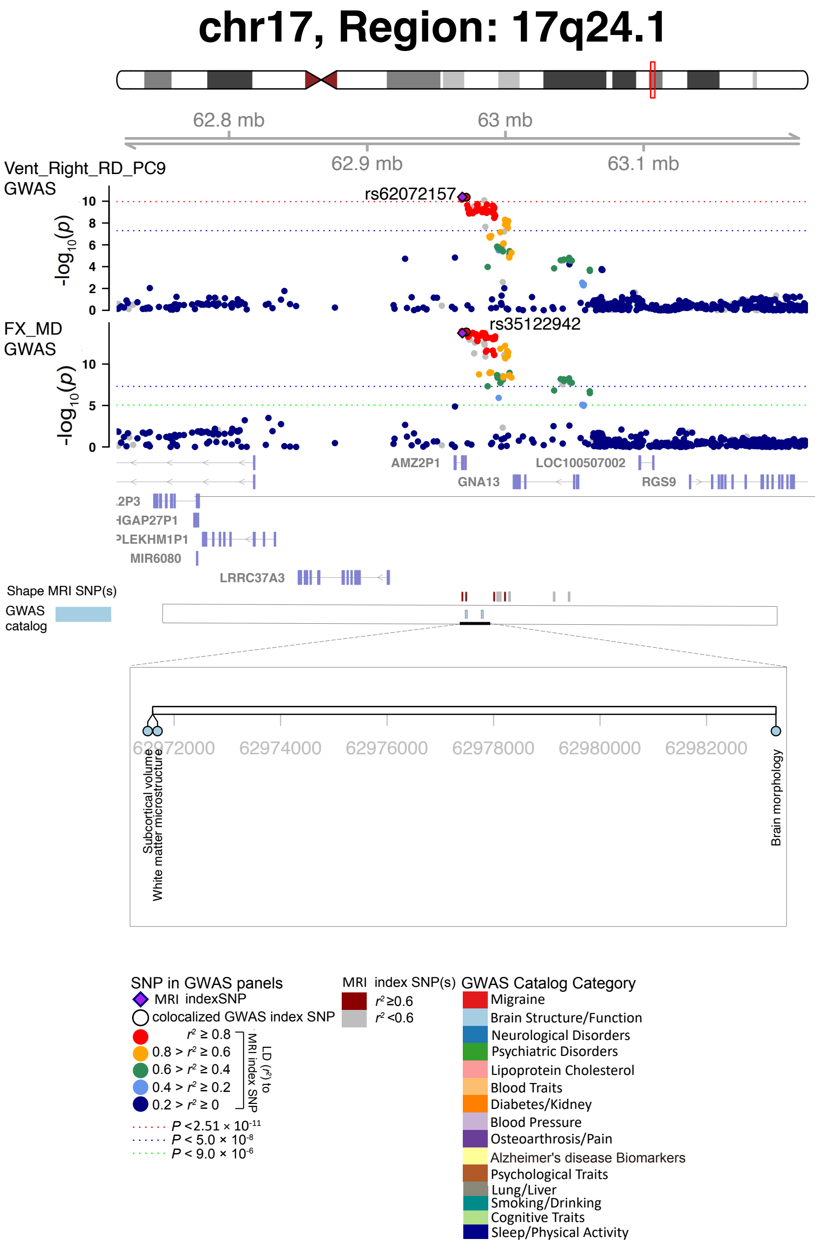


**Fig. S21 Selected genetic locus that were associated with both shape features and other complex traits.**

In 17q.24.1, we observed shared genetic influences (LD $r^2 \geq 0.6$) between shape features (e.g., Vent_Right_RD_PC9, index variant rs62072157) and DTI parameters (e.g., FX_MD, index variant rs35122942). Vent_Right_RD_PC9, the ninth PC of radial distance in the right ventricles; FX_MD, mean diffusivity (MD) of fornix (column and body of fornix).

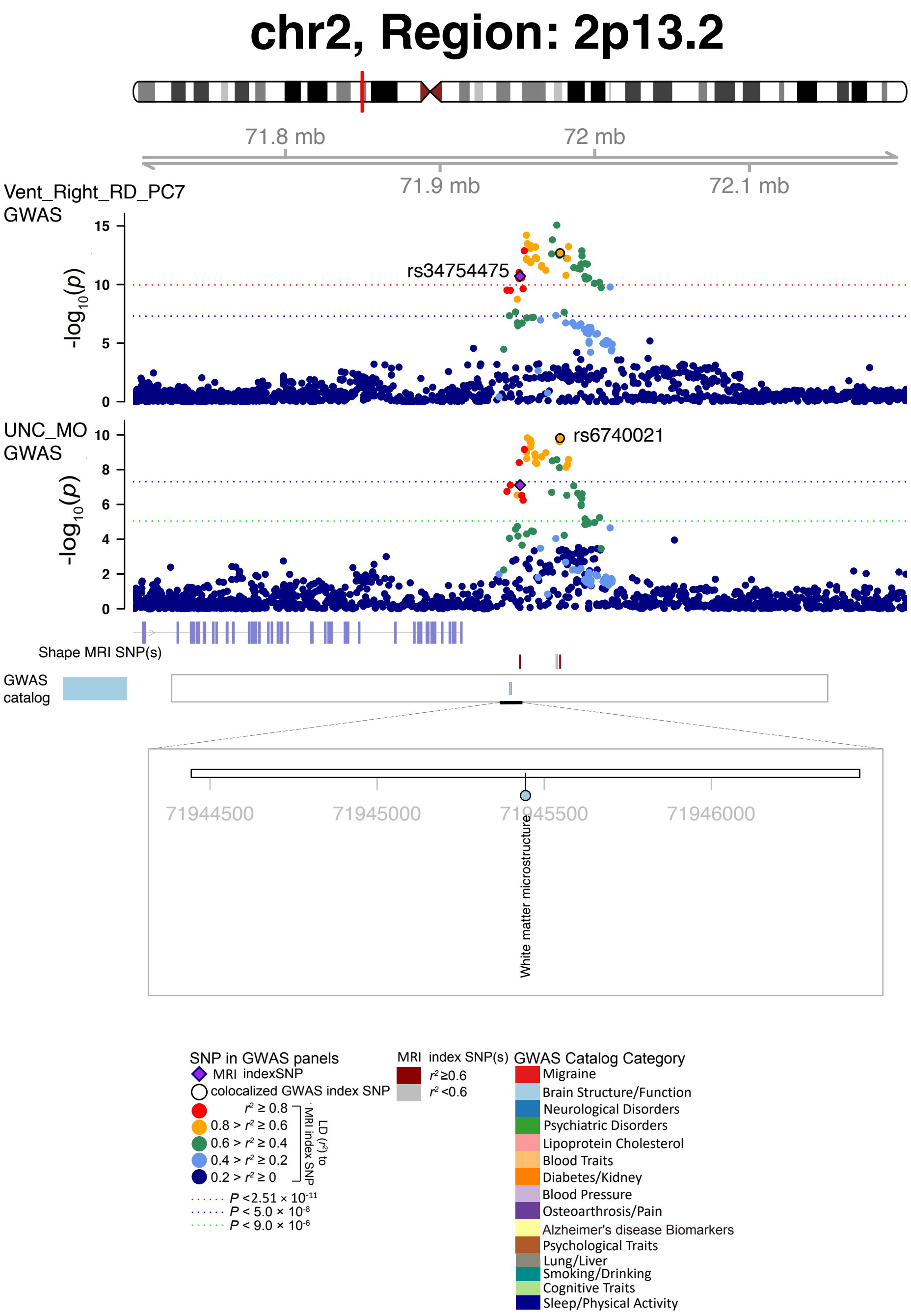


**Fig. S22 Selected genetic locus that were associated with both shape features and other complex traits.**

In 2p.13.2, we observed shared genetic influences (LD $r^2 \geq 0.6$) between shape features (e.g., Vent_Right_RD_PC7, index variant rs34754475) and DTI parameters (e.g., UNC_MO, index variant rs6740021). Vent_Right_RD_PC7, the seventh PC of radial distance in the right ventricles; UNC_MO, mean mode of anisotropy (MO) of uncinate fasciculus.

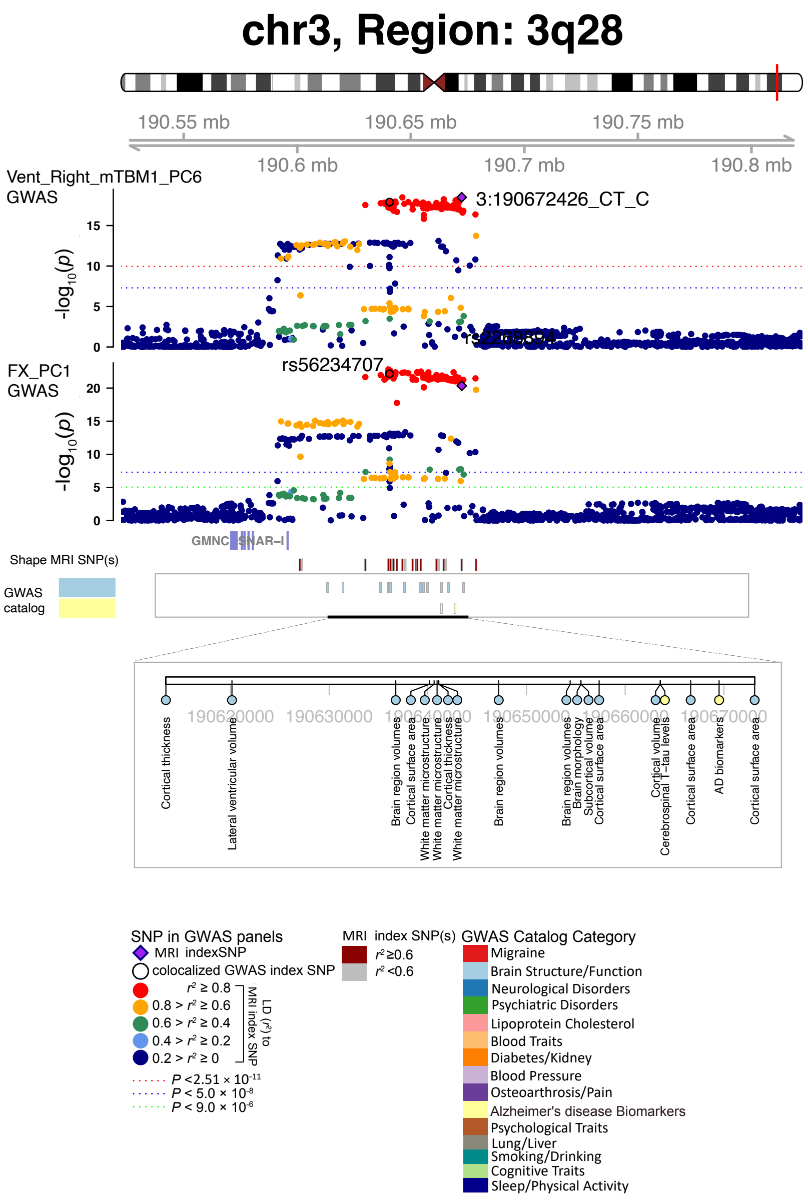


**Fig. S23 Selected genetic locus that were associated with both shape features and other complex traits.**

In 3q28, we observed shared genetic influences (LD $r^2 \geq 0.6$) between shape features (e.g., Vent_Right_mTBM1_PC6, index variant 3:190672426_CT_C) and DTI parameters (e.g., FX_PC1, index variant rs56234707). Vent_Right_mTBM1_PC6, the sixth PC of mTBM1 in the right ventricles; FX_PC1, the first fractional anisotropy PC in fornix (column and body of fornix).

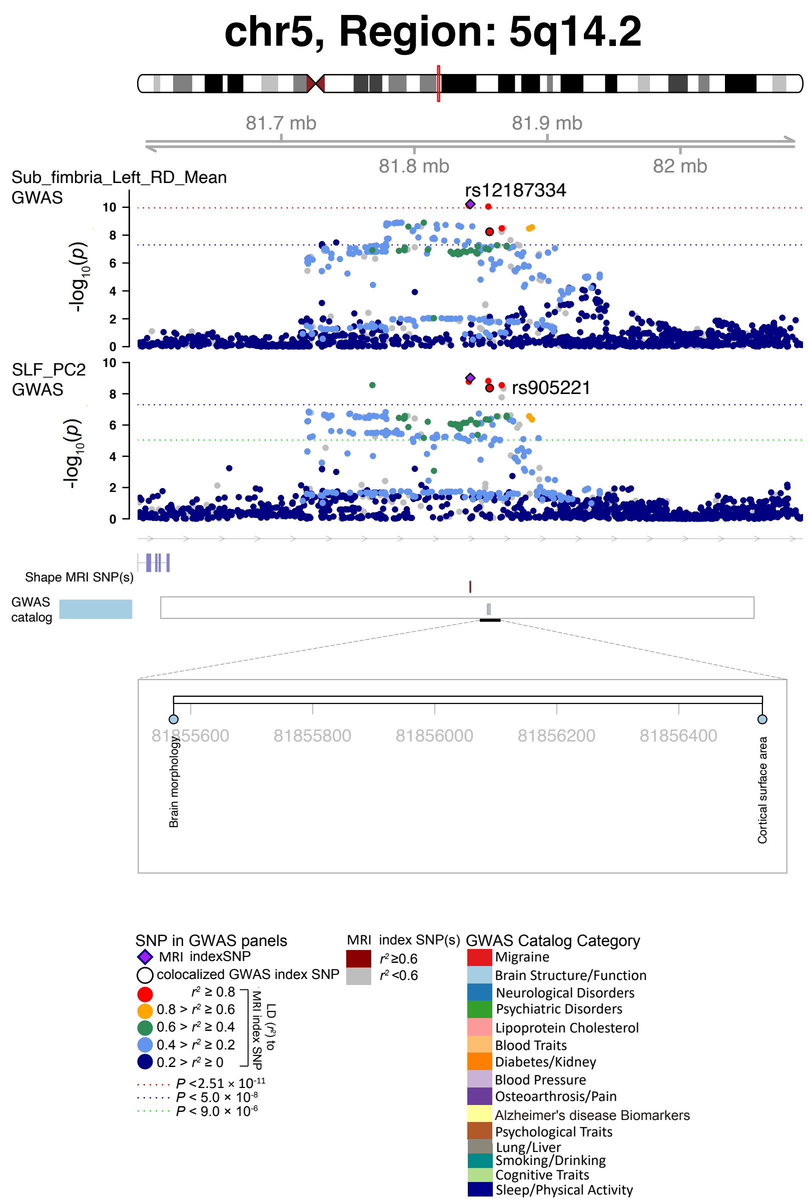


**Fig. S24 Selected genetic locus that were associated with both shape features and other complex traits.**

In 5q14.2, we observed shared genetic influences (LD $r^2 \geq 0.6$) between shape features (e.g., Sub_fimbria_Left_RD_Mean, index variant rs2279829) and DTI parameters (e.g., SLF_PC2, index variant rs2279829). Sub_fimbria_Left_RD_Mean, mean of radial distance in the left hippocampal subfield fimbria; SLF_PC2, the second fractional anisotropy PC in superior longitudinal fasciculus.

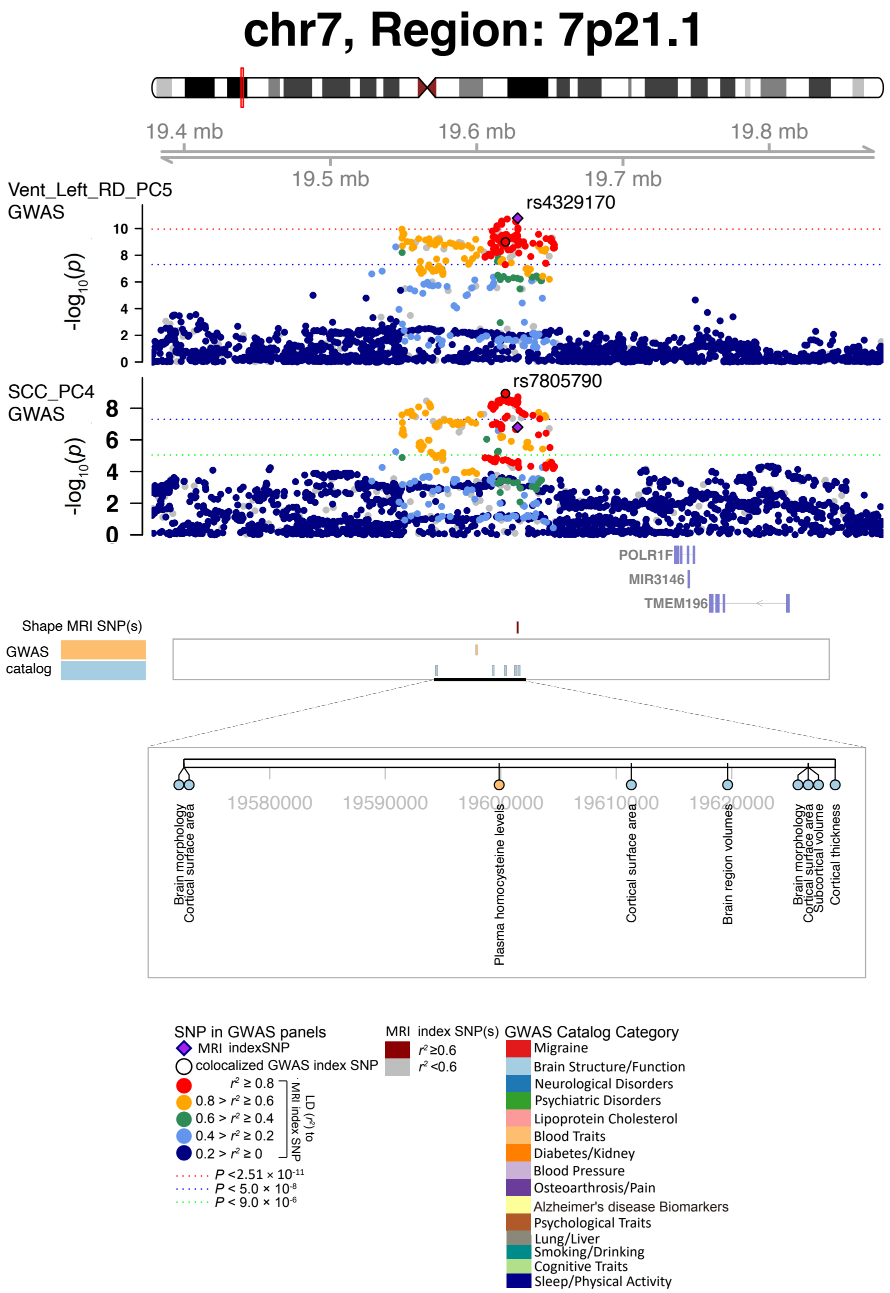


**Fig. S25 Selected genetic locus that were associated with both shape features and other complex traits.**

In 7p21.1, we observed shared genetic influences (LD $r^2 \geq 0.6$) between shape features (e.g., Vent_Left_RD_PC5, index variant rs4329170) and DTI parameters (e.g., SCC_PC4, index variant rs7805790). Vent_Left_RD_PC5, the fifth PC of radial distance in the left ventricles; SCC_PC4, the fourth fractional anisotropy PC in splenium of corpus callosum.

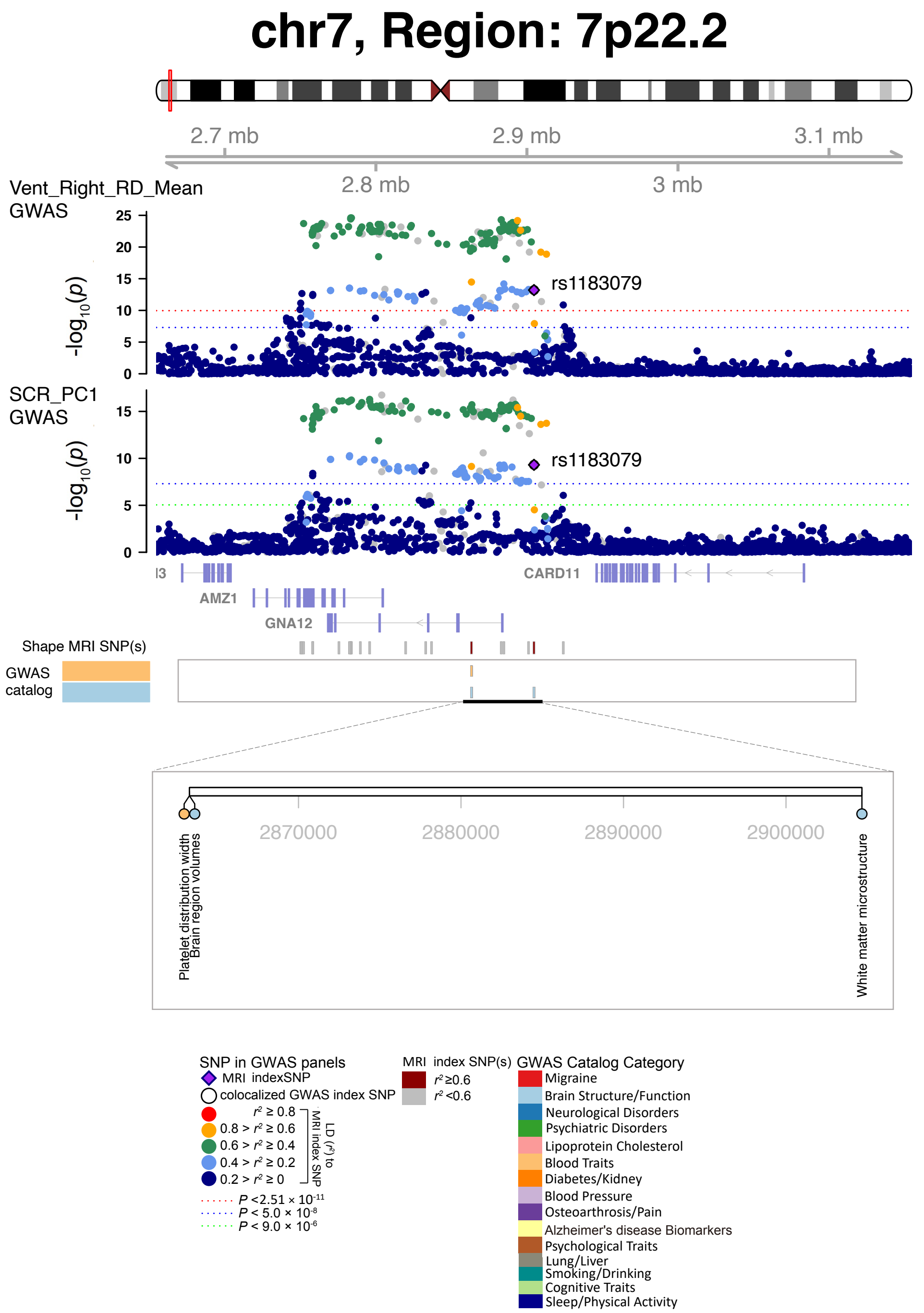


**Fig. S26 Selected genetic locus that were associated with both shape features and other complex traits.**

In 7p22.2, we observed shared genetic influences (LD $r^2 \geq 0.6$) between shape features (e.g., Vent_Right_RD_Mean, index variant rs1183079) and DTI parameters (e.g., SCR_PC1, index variant rs1183079). Vent_Right_RD_Mean, mean of radial distance in the right ventricles; SCR_PC1, the first fractional anisotropy PC in superior corona radiata.

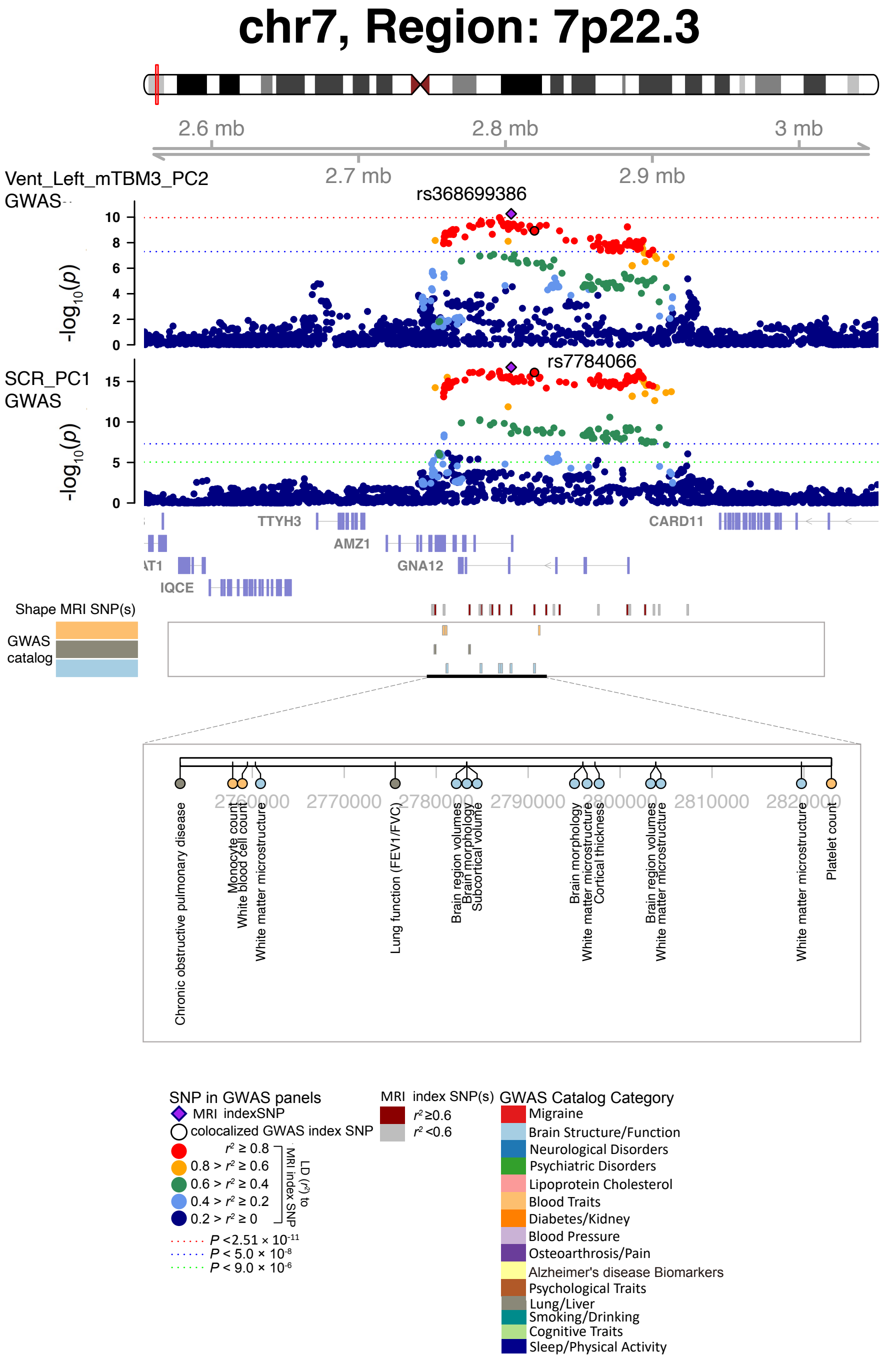


**Fig. S27 Selected genetic locus that were associated with both shape features and other complex traits.**

In 7p22.3, we observed shared genetic influences (LD $r^2 \geq 0.6$) between shape features (e.g., Vent_Left_mTBM3_PC2, index variant rs368699386) and DTI parameters (e.g., SCR_PC1, index variant rs7784066). Vent_Left_mTBM3_PC2, the second PC of mTBM3 in the left ventricles; SCR_PC1, the first fractional anisotropy PC in superior corona radiata.

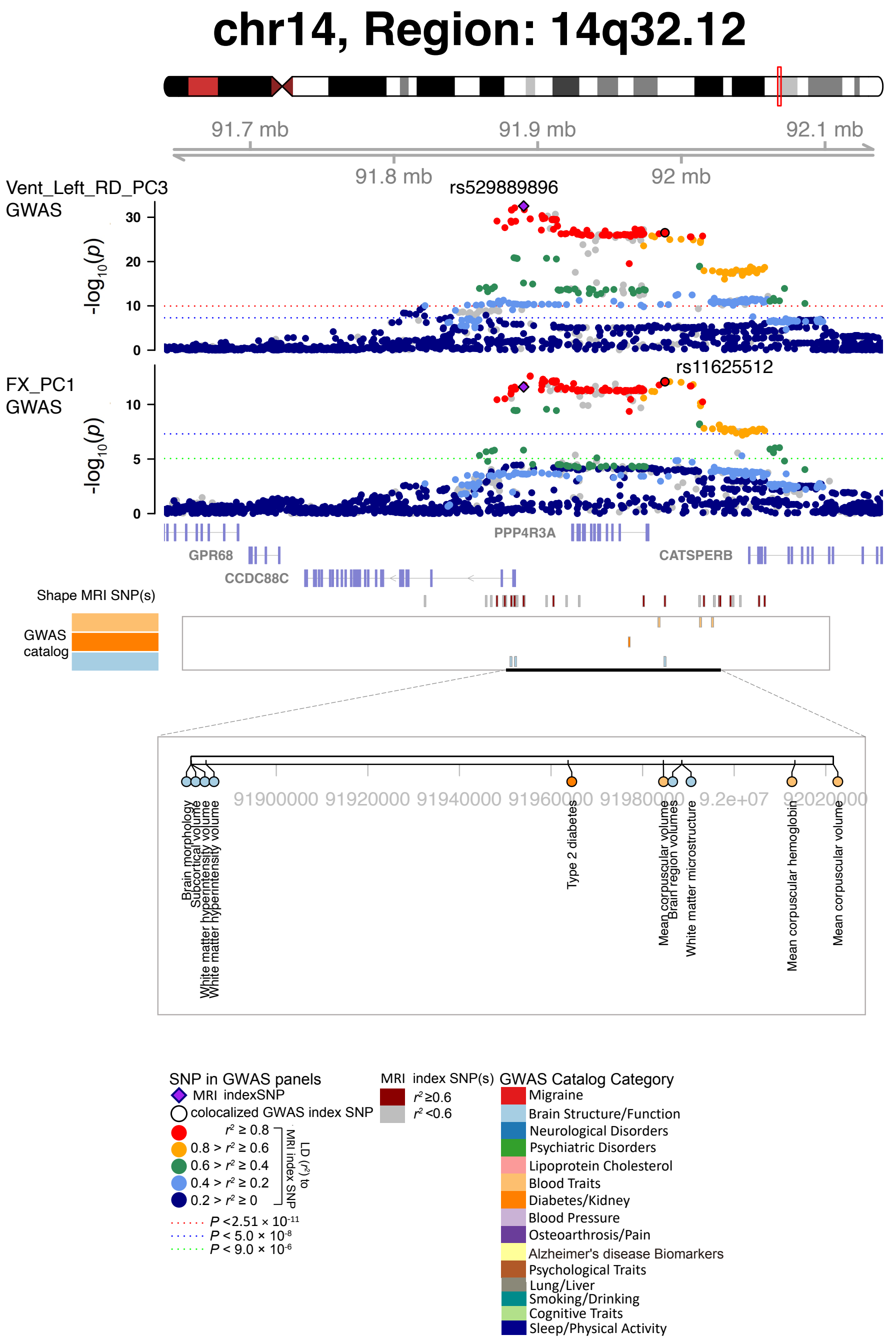


**Fig. S28 Selected genetic locus that were associated with both shape features and other complex traits.**

In 14q32.12, we observed shared genetic influences (LD $r^2 \geq 0.6$) between shape features (e.g., Vent_Left_RD_PC3, index variant rs529889896) and DTI parameters (e.g., FX_PC1, index variant rs11625512). Vent_Left_RD_PC3, the third PC of radial distance in the left ventricles; FX_PC1, the first fractional anisotropy PC in fornix (column and body of fornix).

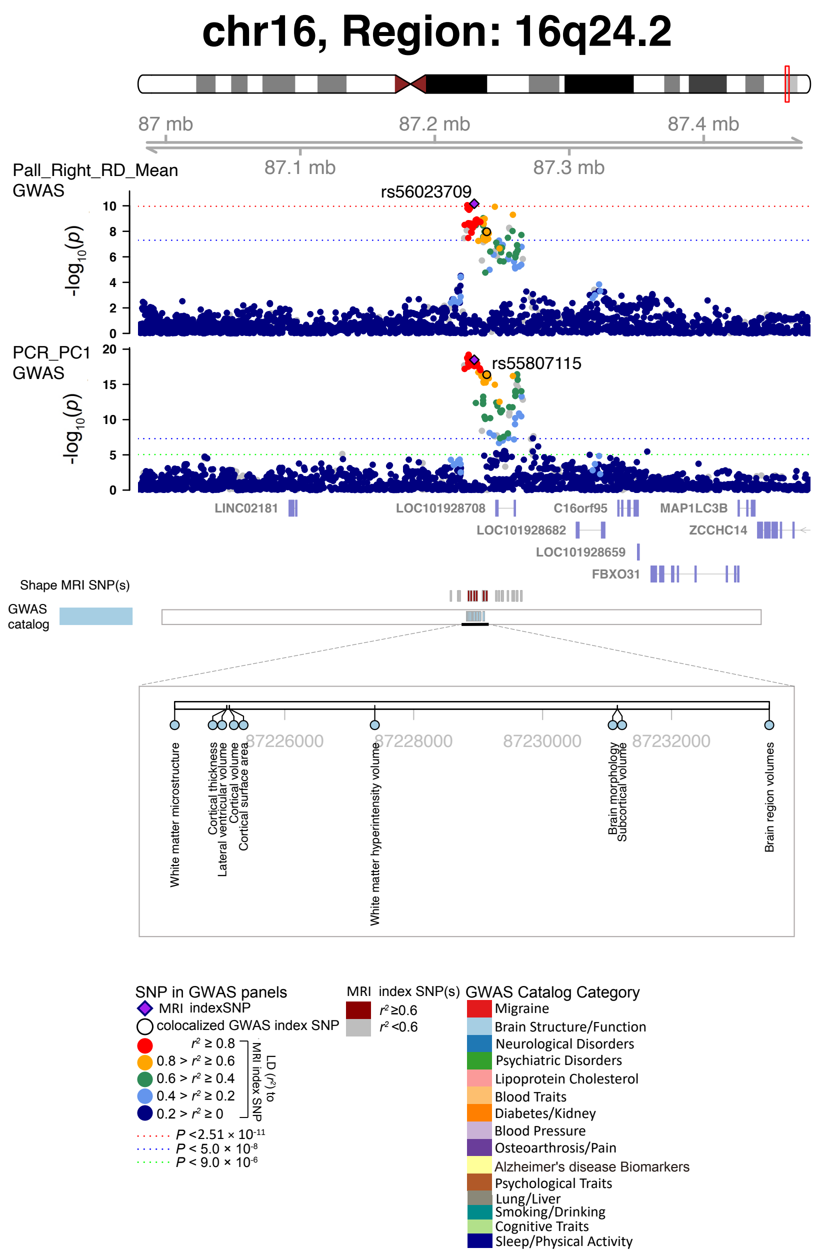


**Fig. S29 Selected genetic locus that were associated with both shape features and other complex traits.**

In 16q24.2, we observed shared genetic influences (LD $r^2 \geq 0.6$) between shape features (e.g., Pall_Right_RD_Mean, index variant rs56023709) and DTI parameters (e.g., PCR_PC1, index variant rs55807115). Pall_Right_RD_Mean, mean of radial distance in the right pallidum; PCR_PC1, the first fractional anisotropy PC in posterior corona radiata.

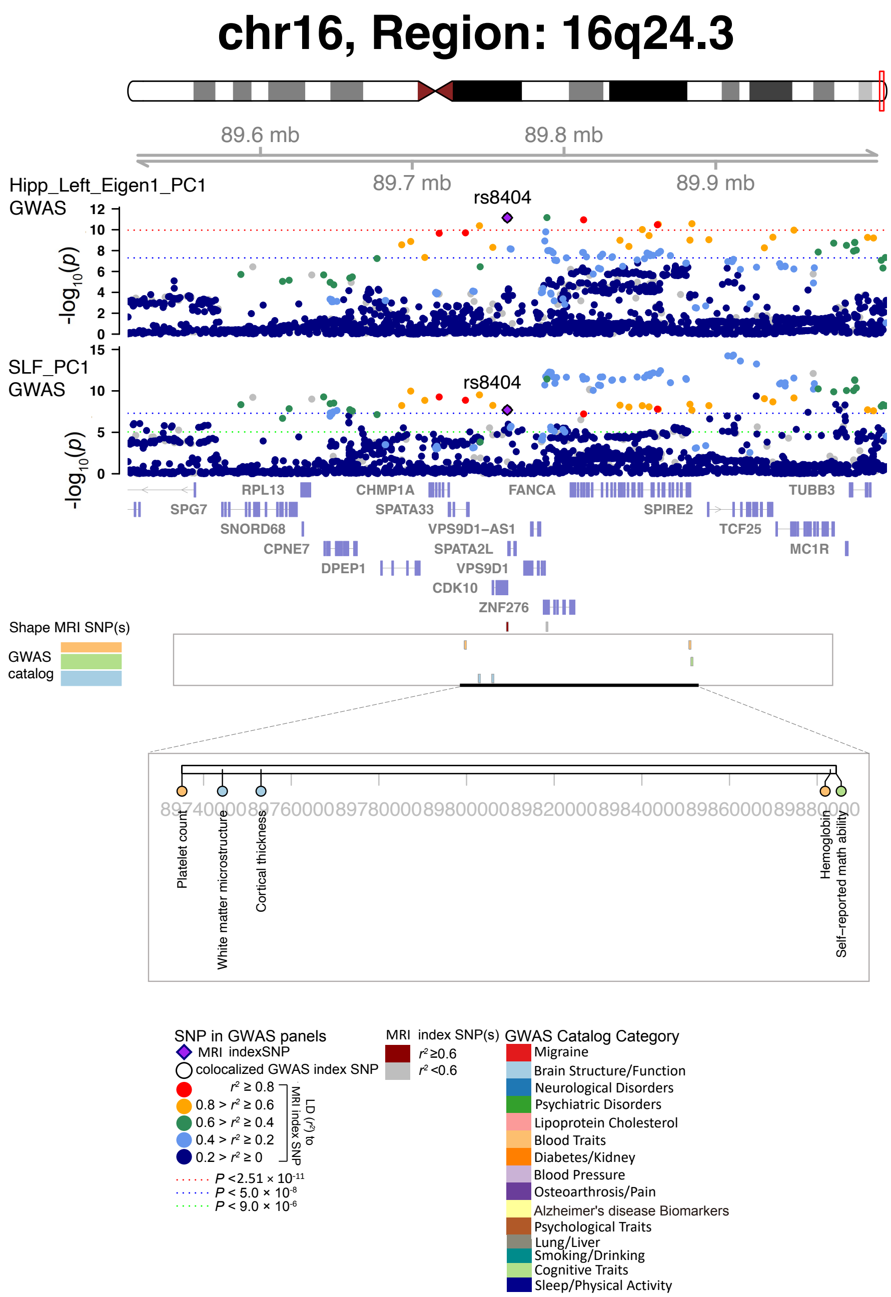


**Fig. S30 Selected genetic locus that were associated with both shape features and other complex traits.**

In 16q24.3, we observed shared genetic influences (LD $r^2 \geq 0.6$) between shape features (e.g., Hipp_Left_Eigen1_PC1, index variant rs8404) and DTI parameters (e.g., SLF_PC1, index variant rs8404). Hipp_Left_Eigen1_PC1, the first PC of the largest eigenvalue in the left hippocampus; SLF_PC1, the first fractional anisotropy PC in superior longitudinal fasciculus.

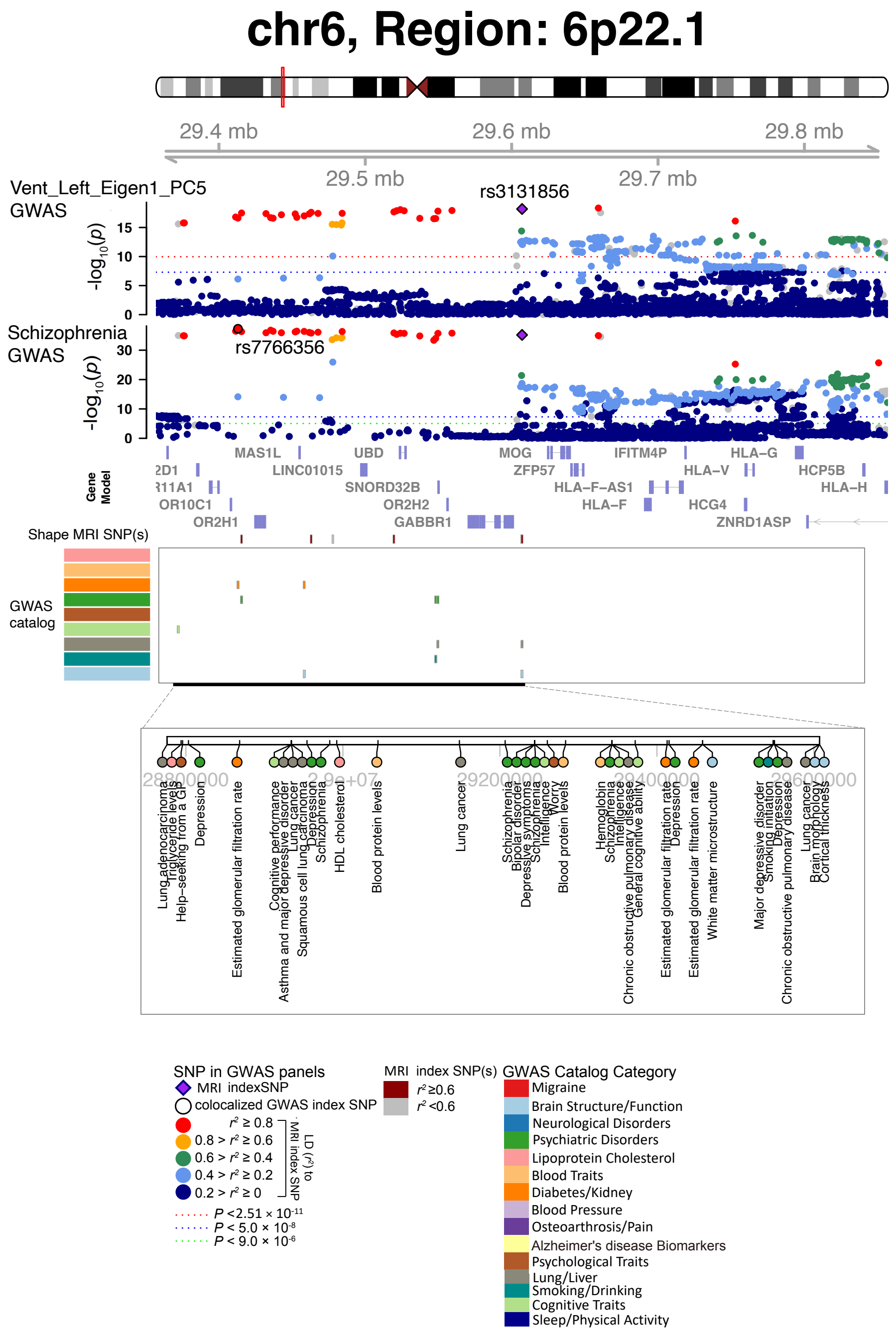


**Fig. S31 Selected genetic locus that were associated with both shape features and other complex traits.**

In 6p22.1, we observed shared genetic influences (LD $r^2 \geq 0.6$) between shape features (e.g., Vent_Left_Eigen1_PC5, index variant rs3131856) and schizophrenia (index variant rs7766356). Vent_Left_Eigen1_PC5, the fifth PC of the largest eigenvalue in the left ventricles.

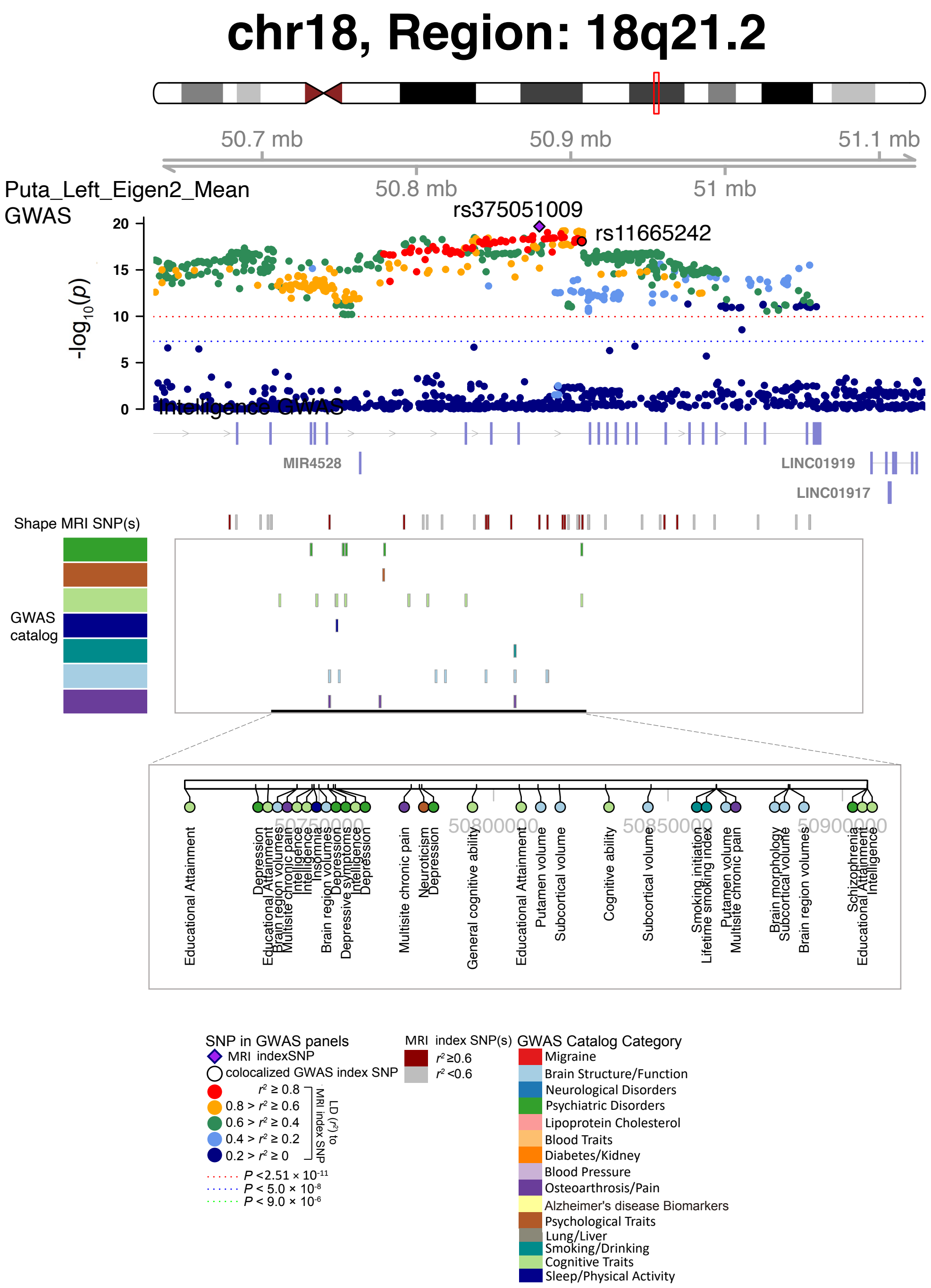


**Fig. S32 Selected genetic locus that were associated with both shape features and other complex traits.**

In 18q21.2, we observed shared genetic influences (LD $r^2 \geq 0.6$) between shape features (e.g., Puta_Left_Eigen2_Mean, index variant rs375051009) and schizophrenia (index variant rs11665242). Puta_Left_Eigen2_Mean, mean of the second largest eigenvalue of the left putamen.

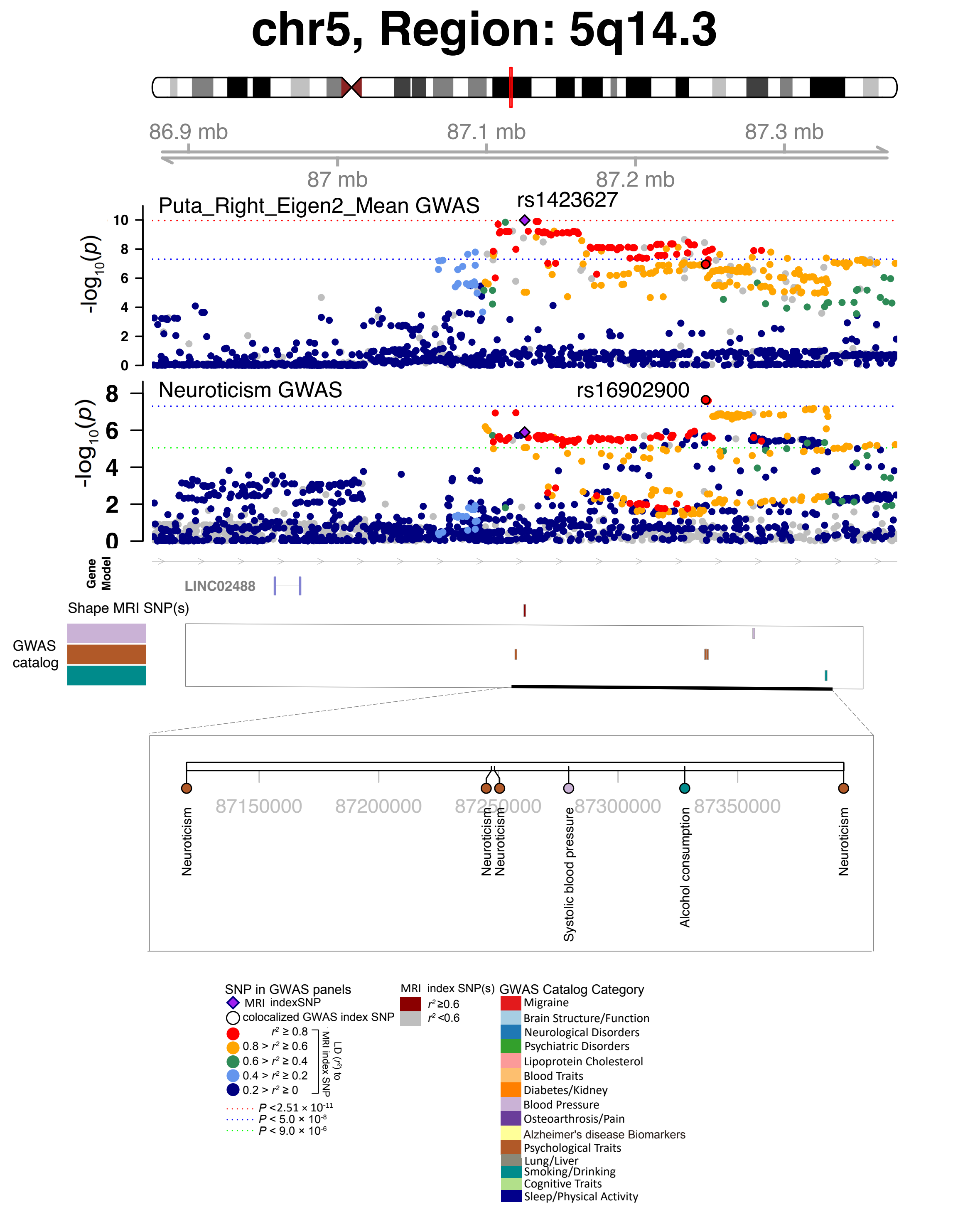


**Fig. S33 Selected genetic locus that were associated with both shape features and other complex traits.**

In 5q14.3, we observed shared genetic influences (LD $r^2 \geq 0.6$) between shape features (e.g., Puta_Right_Eigen2_Mean, index variant rs1423627) and neuroticism (index variant rs16902900). Puta_Right_Eigen2_Mean, mean of the second largest eigenvalue of the right putamen.

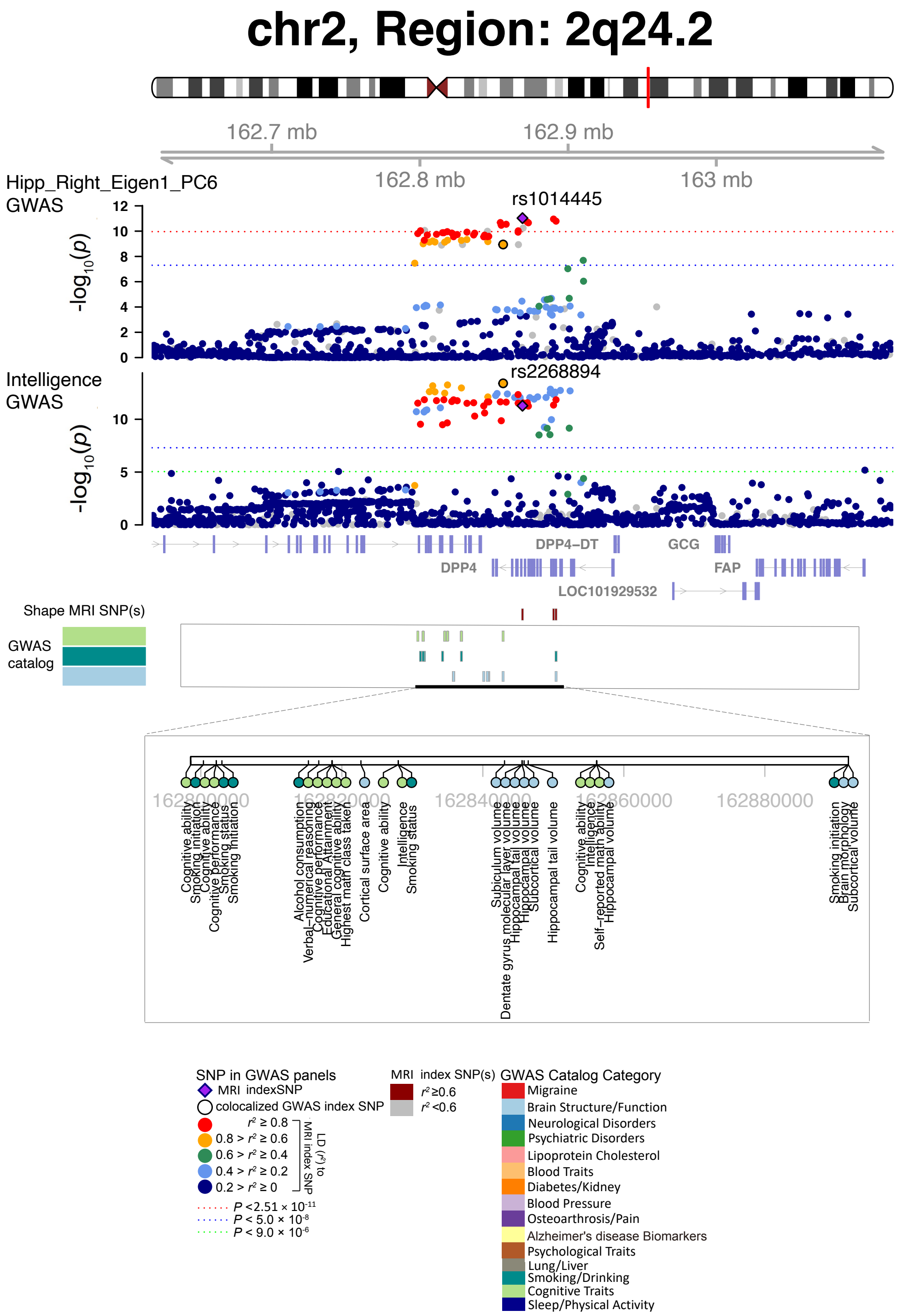


**Fig. S34 Selected genetic locus that were associated with both shape features and other complex traits.**

In 2q24.2, we observed shared genetic influences (LD $r^2 \geq 0.6$) between shape features (e.g., Hipp_Right_Eigen1_PC6, index variant rs1014445) and intelligence (index variant rs2268894). Hipp_Right_Eigen1_PC6, the sixth PC of the largest eigenvalue in the right hippocampus.

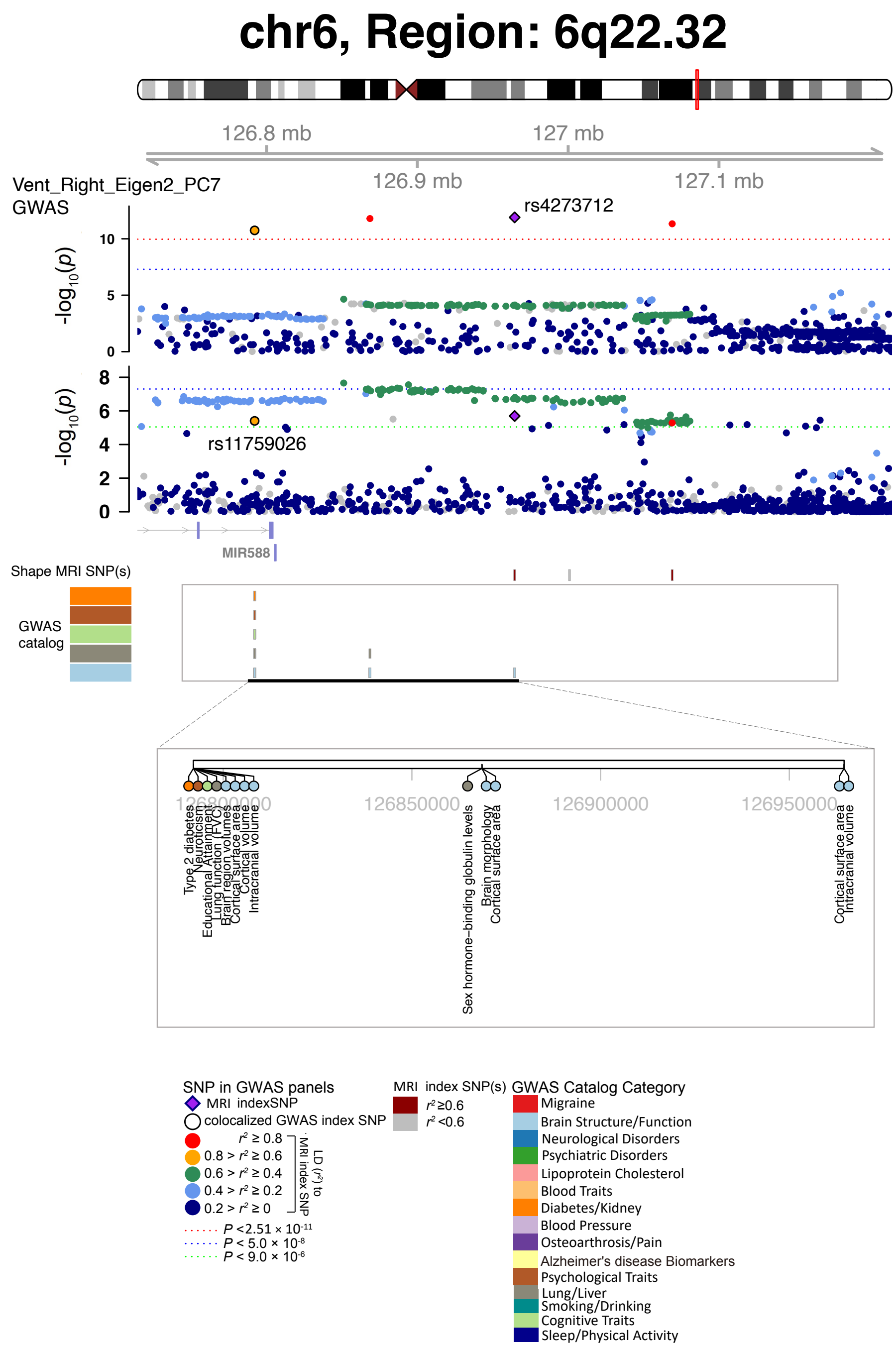


**Fig. S35 Selected genetic locus that were associated with both shape features and other complex traits.**

In 6q22.32, we observed shared genetic influences (LD $r^2 \geq 0.6$) between shape features (e.g., Vent_Right_Eigen2_PC7, index variant rs4273712) and educational attainment (index variant rs11759026). Vent_Right_Eigen2_PC7, the seventh PC of the second largest eigenvalue in the right ventricles.

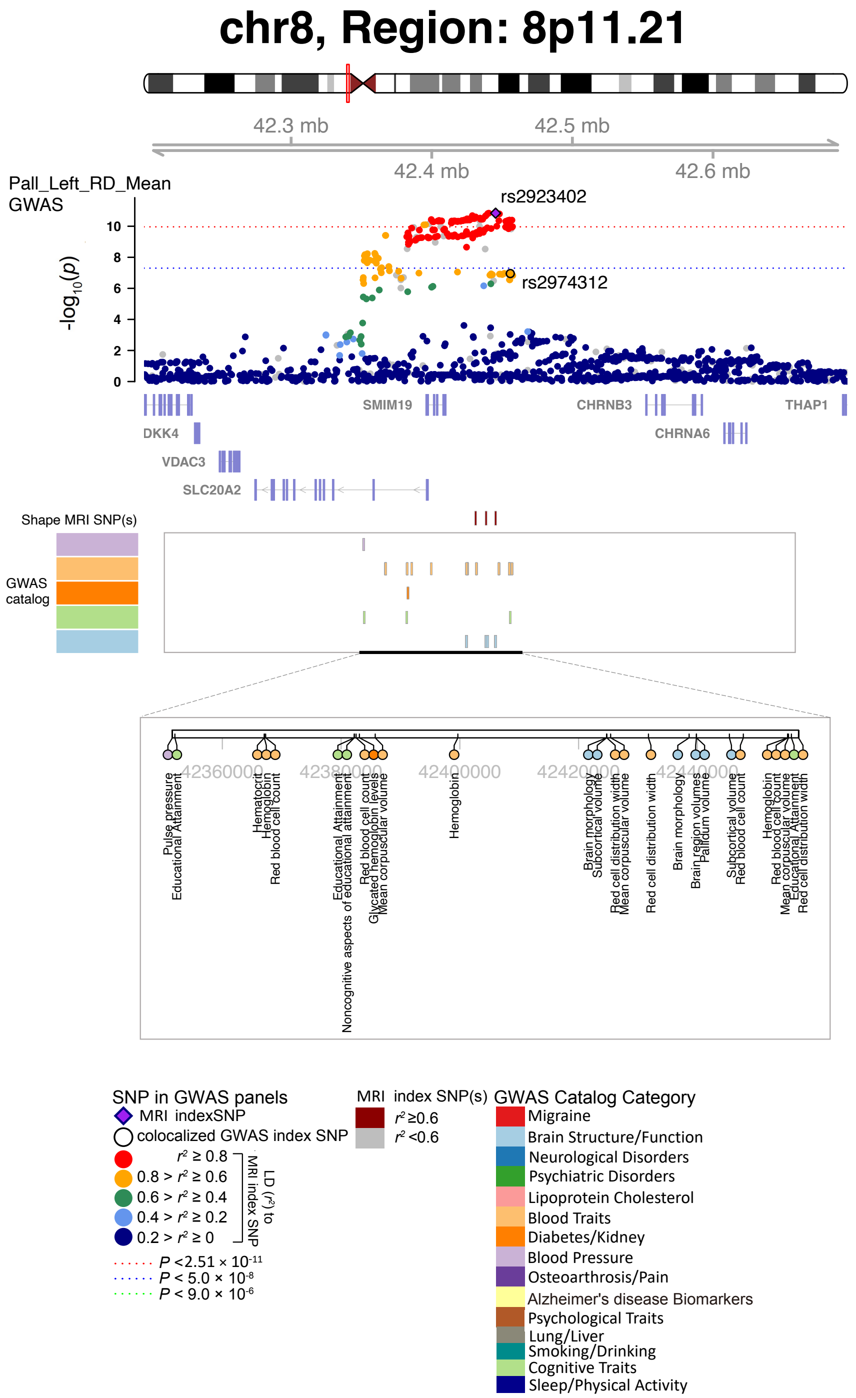


**Fig. S36 Selected genetic locus that were associated with both shape features and other complex traits.**

In 8p11.21, we observed shared genetic influences (LD $r^2 \geq 0.6$) between shape features (e.g., Pall_Left_RD_Mean, index variant rs2923402) and educational attainment (index variant rs2974312). Pall_Left_RD_Mean, mean of radial distance in the left pallidum.

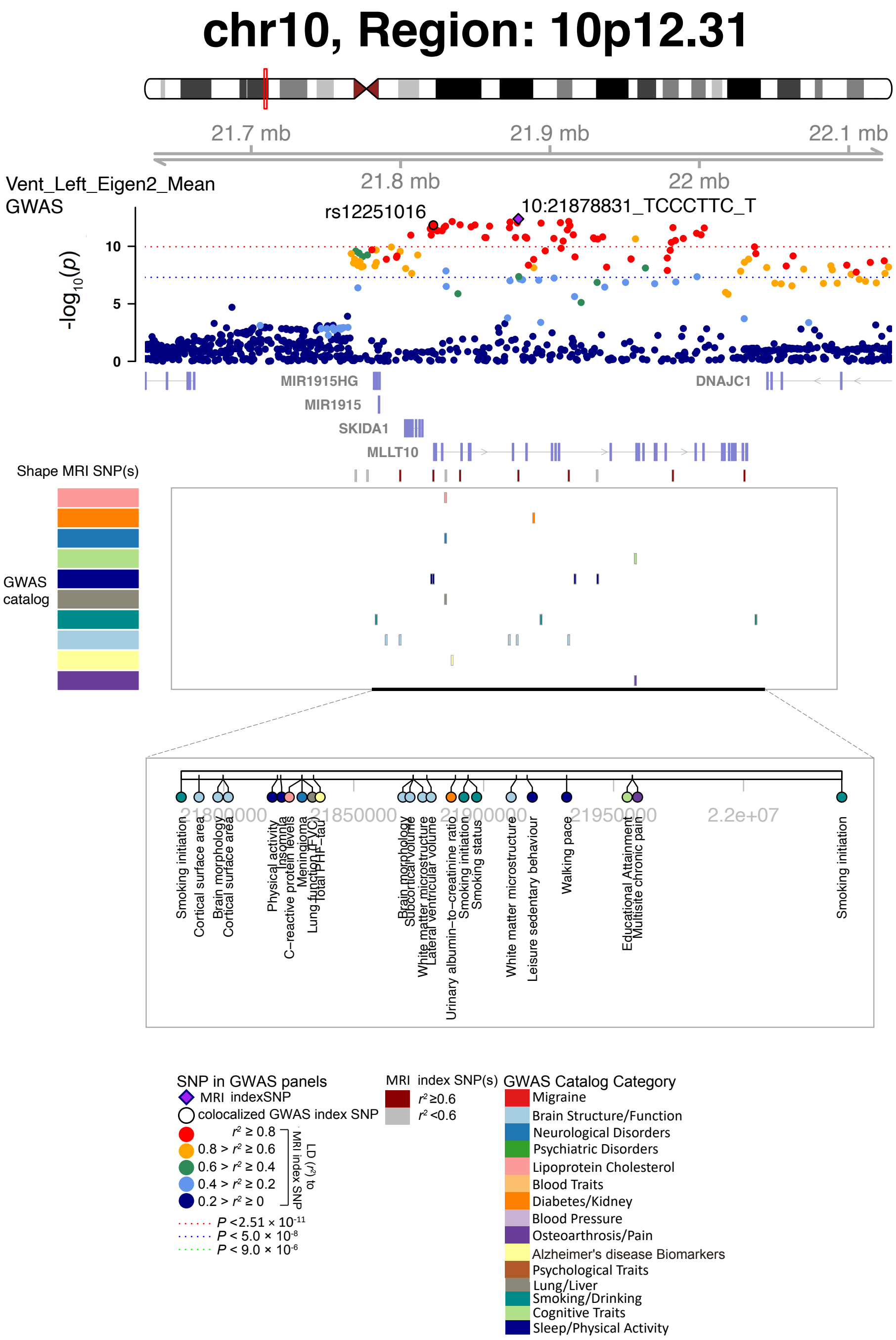


**Fig. S37 Selected genetic locus that were associated with both shape features and other complex traits.**

In 10p12.31, we observed shared genetic influences (LD $r^2 \geq 0.6$) between shape features (e.g., Vent_Left_Eigen2_Mean, index variant 10:21878831_TCCCTTC_T) and insomnia (index variant rs12251016). Vent_Left_Eigen2_Mean, mean of the second largest eigenvalue in the left ventricles.

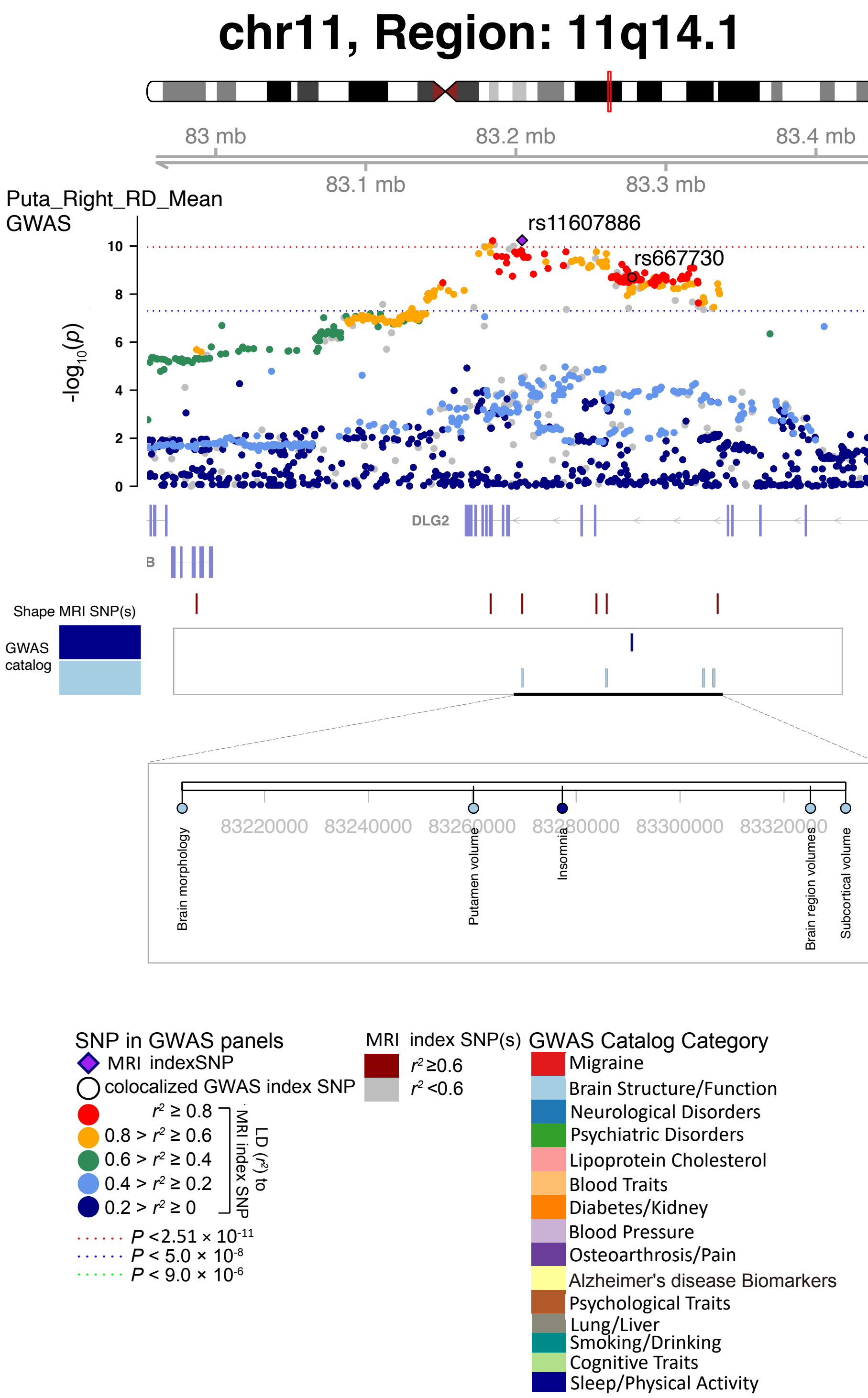


**Fig. S38 Selected genetic locus that were associated with both shape features and other complex traits.**

In 11q14.1, we observed shared genetic influences (LD $r^2 \geq 0.6$) between shape features (e.g., Puta_Right_RD_Mean, index variant rs11607886) and insomnia (index variant rs667730). Puta_Right_RD_Mean, mean of radial distance in the right putamen.

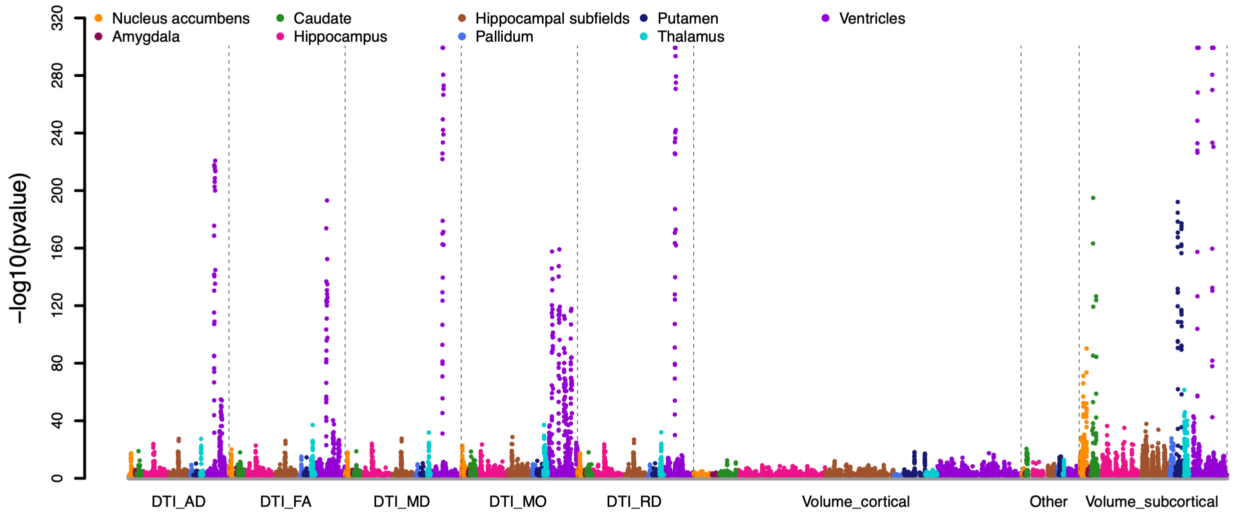


**Fig. S39 Genetic correlation estimates between shape features and other brain structural measures.**

We tested genetic correlations between 457 shape features (from ventricular and subcortical structures) and 211 brain structural traits, including 110 DTI parameters (AD, FA, MD, MO, and RD in different white matter tracts) and 101 regional brain volumes (cortical, subcortical, and other brain structures). AD, axial diffusivity; FA, fractional anisotropy; MD, mean diffusivity; MO, mode of anisotropy; and RD, radial diffusivity.

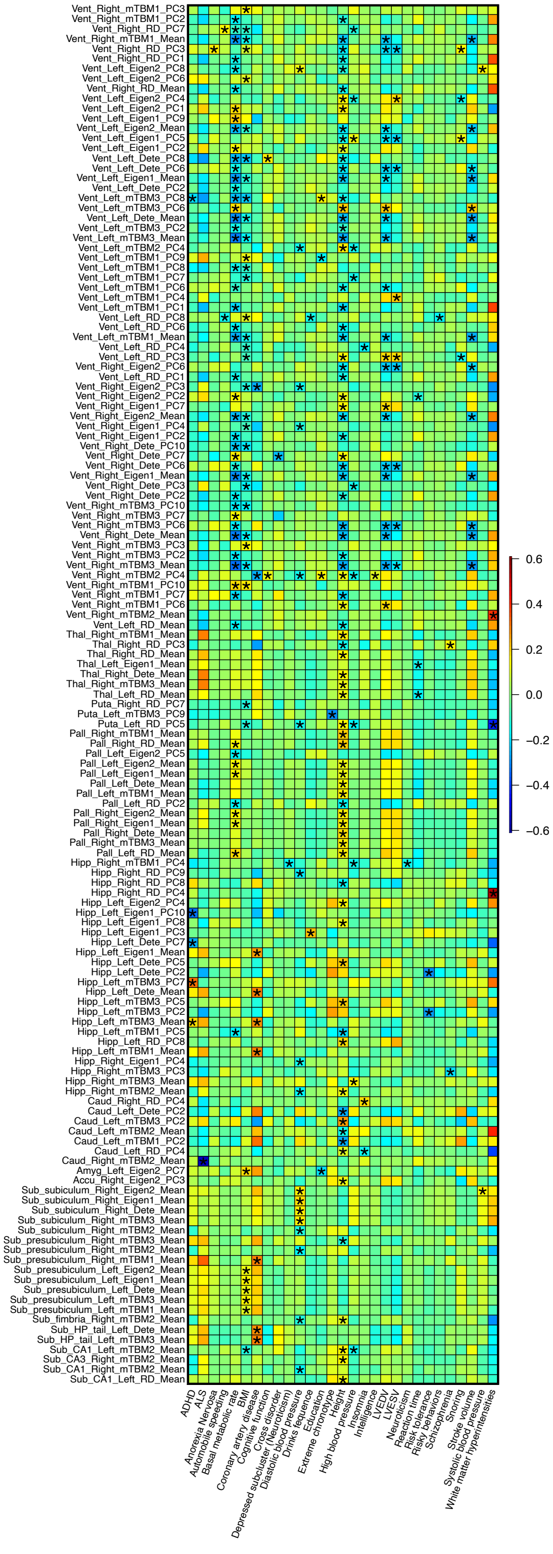


**Fig. S40 Significant genetic correlations between shape features and complex traits and diseases.**

We show the map of significant genetic correlations at the FDR 5% level (457 × 48 tests).

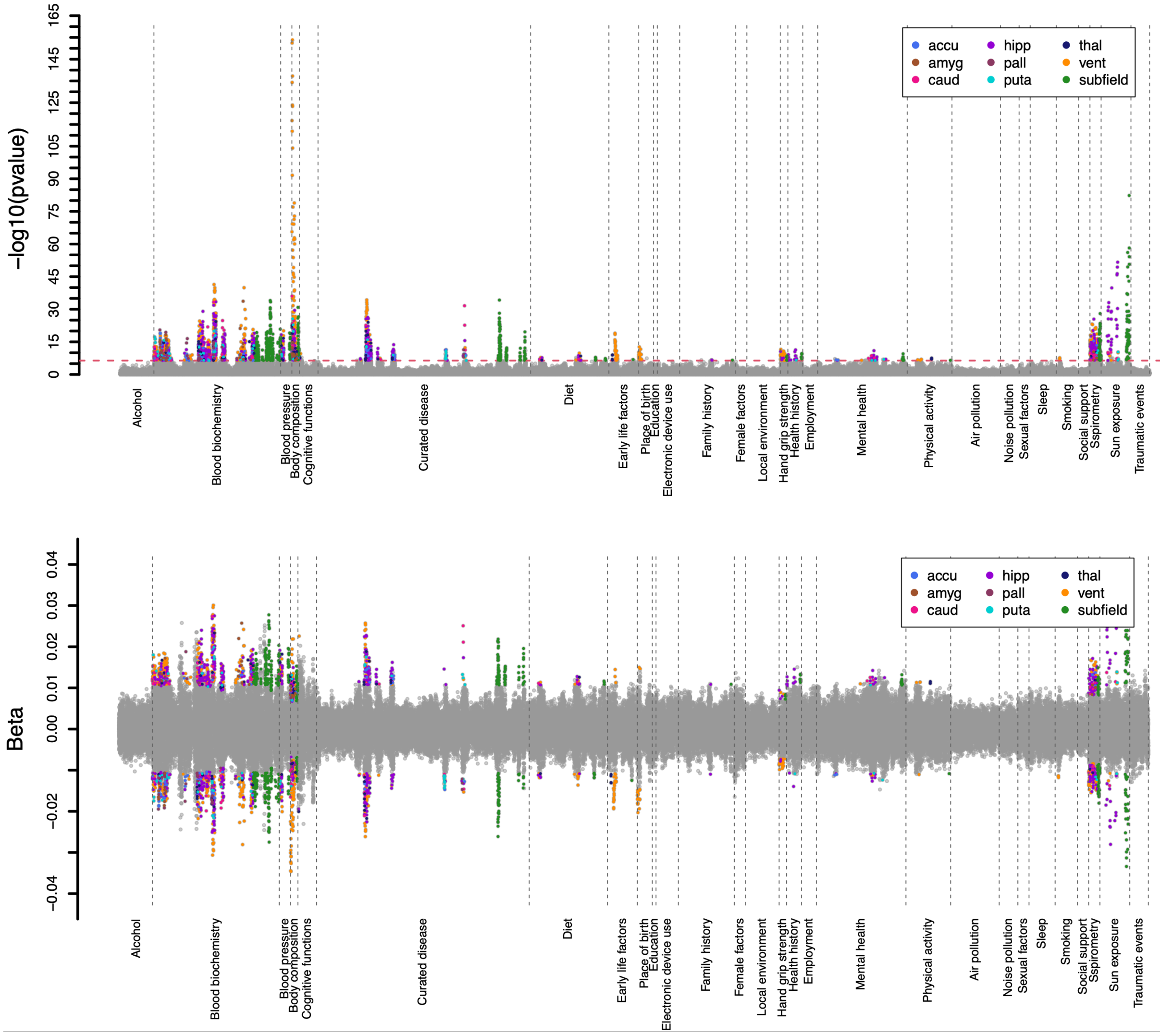


**Fig. S41 Correlations between complex traits and polygenic risk scores (PRS) of shape features.**

We highlighted the associations whose *P*-value pass the Bonferroni multiple testing ($P < 1.33 \times 10^{-5}$) in the discovery sample and were validated in an independent hold-out validation sample. See Table S11 for more information of these complex traits and diseases.

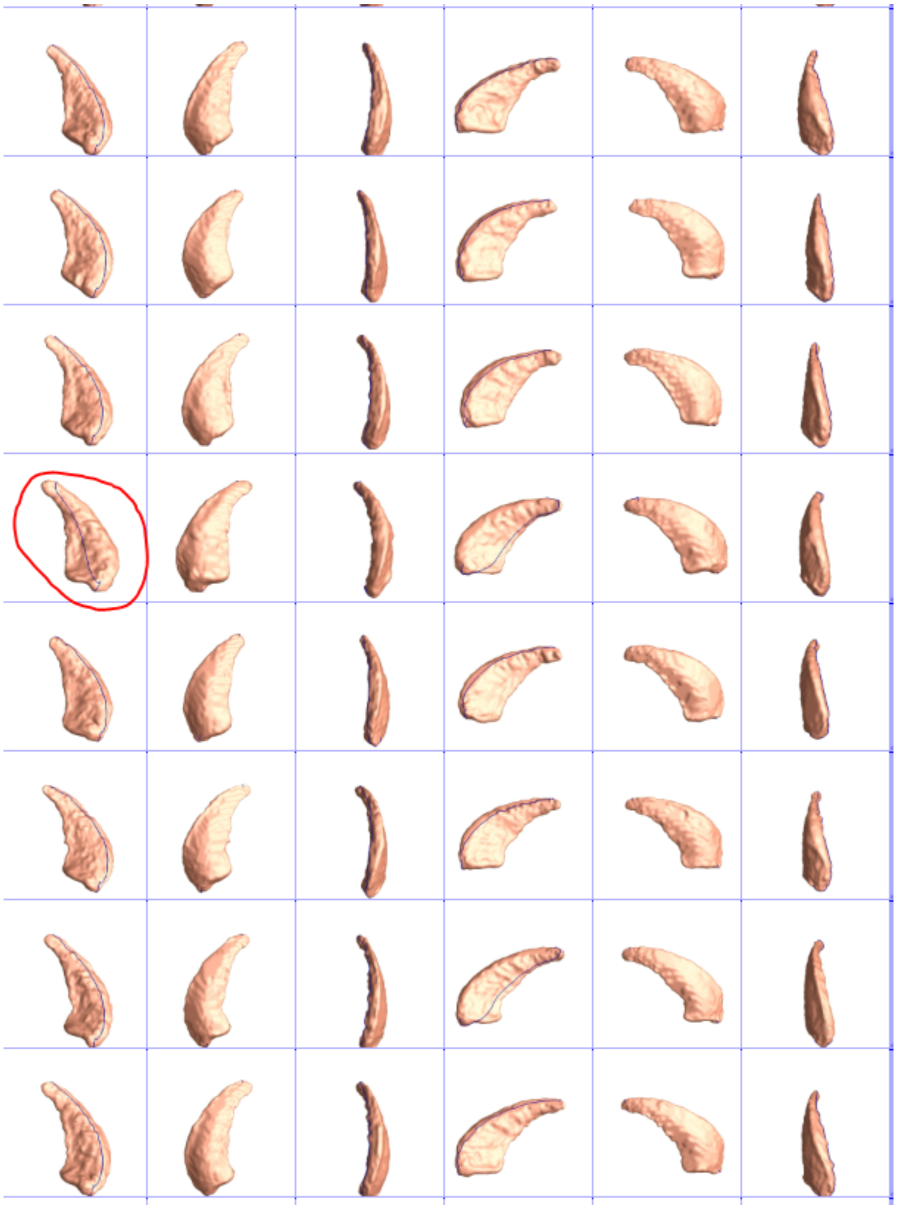

**Fig. S42 Example of surface registration quality check.**

We generated snapshots for each surface of outlier subjects. We draw landmark in blue line in the individual space to show the same location in the standard space. We manually checked if the landmarks indicated the same location for each subject to evaluate the surface registration. For example, the 4th subject in red circle indicates a bad surface registration because its blue line landmark location is different from other subjects. This subject will be removed from our analysis.
