## Supplementary material for "Genetic influences on the shape of brain ventricular and subcortical structures": supp_info

**This PDF file includes:**
Supplementary Note

**Other Supplementary Materials for this manuscript include the following:**
Supplementary Figures S1 to S42 (available in a PDF file)
Supplementary Tables S1 to S14 (.xlsx) (available in a zip file)

### Supplementary Note

#### Shape analysis pipeline

Using the FMRIB Software Library (FSL)[1] (https://fsl.fmrib.ox.ac.uk/) and Statistical Parametric Mapping (SPM) (http://www.fil.ion.ucl.ac.uk/spm/) packages, structural MRIs were linearly registered into a standard brain space (MNI152). Seven subcortical (the amygdala, hippocampus, nucleus accumbens, caudate nucleus, putamen, pallidum, and thalamus) and lateral ventricular structures were segmented[2-4]. Surface meshes were constructed based on the subcortical and ventricular segmentations with a topology-preserving level set method[5] and the marching cube algorithm[6], parameterized with refined triangular meshes using the topological optimization algorithm[7] and holomorphic flow segmentation method[8]. Then the images were registered to a common rectangular grid template using the surface fluid registration algorithm[9]. Multiple morphometry features were extracted, including the multivariate tensor-based morphometry (mTBM), radial distance (RD)[8,10], determinant of the Jacobian matrix, and minimum and maximum eigenvalues of the Jacobian matrix[8]. The overall average and top principal components (PCs) of 7 features for 8 subcortical and ventricular regions and 7 hippocampal subfields were extracted. This pipeline is publicly available at https://www.nitrc.org/frs/?group_id=1461. The detailed steps in our applications were summarized as follows:

1. Individual MRI scans were linearly registered into the MNI152 standard space to remove the effects of individual brain size. Automated subcortical segmentations (the amygdala, hippocampus, nucleus accumbens, caudate nucleus, putamen, pallidum, and thalamus) from the registered MR images were conducted using the FIRST with default parameters in FSL[3]. Since the lateral ventricles had complex geometric structure (i.e., a "multiple-arm" shapes), a different segmentation strategy was applied. Specifically, the cerebrospinal fluid (CSF) was first segmented from the registered individual MR images using the SPM8 packages (http://www.fil.ion.ucl.ac.uk/spm/). A group-wise CSF template was then created by applying the geodesic shooting algorithm[11], which learned the minimal deformation from all the individual's CSF segmentations. The binary ventricular template was extracted by mapping a probability ventricular mask, i.e., the automatic lateral ventricle delineation (ALVIN) binary mask[11], onto this template. The ventricular template boundaries were visually inspected. The deformation matrices from the estimation of CSF template were then used to wrap the ventricular template back to the individual space to generate the individual ventricular segmentations[4].
2. The bilateral subcortical and ventricular surfaces were modeled with a topology-preserving level set method[5]. Based on the voxel-wise binary segmentation results, the marching cubes algorithm[6] was applied to generate the triangular surface meshes. Progressive mesh[12] and loop subdivision[13] methods were applied to refine the generated meshes and down-sample each surface mesh to a consistent number of vertices for subcortical and ventricular structures. Before geometric

analysis of these ventricular surfaces, a two-step mesh smoothing method was applied to remove noise and topologically irregular structures[14].

3. A common geometrical surface structure was defined so that all surfaces can be consistently parameterized[15]. To generate a planar surface conformal parameterization for a closed subcortical surface, the topological optimization algorithm [7] was applied to convert each kind of subcortical surfaces to a tube-like surface with two open consistent boundaries across subjects. The holomorphic 1-form method[16] induced conformal grids which demonstrated the angle preserving property on the tube-like subcortical surfaces. Due to the naturally occurring 3-horn shape, the whole ventricular surface was cut into three sub-structures by the holomorphic 1-forms segmentation method[8].
4. Each individual parameterized subcortical or sub-ventricular surface was registered to a common template surface. With conformal representations[9], a 3D subcortical or sub-ventricular surface can be realized as a 2D image so that general image registration algorithms can be applied. A surface fluid registration algorithm was carried out and an inverse-consistent surface registration framework was added to increase robustness[9,17,18].
5. After surface parameterization and registration, we computed vertex-wise features with multivariate morphometry statistics, including vertex-wise morphometry features along the surface tangent direction (such as the mTBM) and normal direction (i.e., RD)[8,19,20]. We also calculated three more vertex-wise statistics in the standard space, derived from the Jacobian matrix measuring local area differences: the min/max eigenvalues of the Jacobian matrix and the determinant of Jacobian matrix[8].
6. We calculated the average of each shape feature map for each of the subcortical and ventricular regions. We picked out the outlier subjects for each feature map if the average shape feature was greater than five times the median absolute deviation (MAD) from the population. In addition, we generated snapshots for each surface of those outlier subjects, with landmarks drawn in blue line in the individual space to show the same location in the standard space. We manually checked if the landmarks indicated the same location for each subject to evaluate the surface registration. We removed those images with bad surface registration quality (**Fig. S42**).
7. We extracted the top 5 PCs of each feature map for each of the subcortical and ventricular regions. Specifically, for each feature map, we reshaped the vertex-wise UKB phase 3 data (average $n \approx 16{,}000$ subjects) into an $n \times p$ data matrix A, where $p$ is the number of vertices. Top PC scores and PC basis functions for UKB phase 3 data were extracted through singular value decomposition of A. For more details on how to extract PCs from images, please refer to Zhao, et al. [21].
8. The UKB phases 1 and 2 datasets (average $n \approx 20{,}000$ subjects) were used to evaluate the robustness of PC scores in Step 7. For each feature map, the original PC scores and PC basis functions for UKB phases 1 and 2 datasets were extracted in the same procedure as Step 7. Meanwhile, the projected PC scores will be extracted by projecting the UKB phases 1 and 2 data onto the PC basis functions

obtained from the UKB phase 3 data in Step 7. The ICC between the original PC scores and the projected PC scores were calculated to evaluate the reproducibility of PC scores based on the two different PC bases.

9. We segmented the hippocampus surface-based rectangular grid template into 12 subfields (https://surfer.nmr.mgh.harvard.edu/fswiki/HippocampalSubfields) by applying the subfield segmentation algorithm in FreeSurfer v6.0[20] to the MNI152 brain atlas. The average across each of the cornu ammonis 1 (CA1), CA3, fimbria, hippocampus-amygdala-transition-area (HATA), hippocampal tail, presubiculum, and subiculum subfields for each feature map was extracted. Other 5 subfields were excluded because their surfaces were out of view.
